# LEAP-InovAND a multiscale resource to explore genetics, brain imaging and clinical data in autism

**DOI:** 10.1101/2025.11.24.25340858

**Authors:** M. Fleury, Z. M. Mougin, C. S. Leblond, F. Cliquet, A. Mathieu, B. Oakley, A. Maruani, J.-M. de Saint Agathe, S. Korkmaz, R. Holt, A. Vitrac, F. Amsellem, A. Batut, G. Botton-Amiot, T. Rolland, E. Barthome, R. Bonicel, A. Beggiato, N. Lemière, A.-C. Tabet, J. Levy, C. Lehoucq, A. Ayrolles, C. A. Moreau, L. Colomar Molla, R. Toro, N. Traut, A. Lefebvre, D. L. Floris, G. Dumas, G. Huguet, S. Bonnot-Briey, F. Campana, J. Van Gils, V. Wang, J. Buratti, L. Jornea, S. Loiodice, S. Forlani, S. Fournier, D. Bacq-Daïan, R. Olaso, V. Meyer, B. Fin, AIMS2-2-Trials AReps group, A. Ghosh, M. Doherty, S. Li-Williams, InovAND group, EU-AIMS/AIMS-2-Trials LEAP group, J.-F. Deleuze, A. M. Persico, M. Leboyer, C. Gillberg, S. Bölte, C. Beckmann, C. Ecker, E. Jh Jones, V. Warrier, S. Baron-Cohen, T. Charman, E. Loth, J. K. Buitelaar, J. Tillmann, R. Delorme, D. Murphy, T. Bourgeron

## Abstract

Most current autism research focuses on categorical comparisons (e.g., autistic vs. neurotypical people) and usually examines only one biological domain (e.g., cognition, genetics, or brain imaging). Here, we present a resource integrating quantitative phenotypic data, whole genome sequencing, brain magnetic resonance imaging, and electroencephalography. A total of 2,061 autistic people, 2,551 relatives and 875 neurotypical people were recruited in Europe through LEAP-InovAND, including 2,531 participants with both clinical and genetic data, and 875 with additionally neuroimaging data (EEG and/or MRI). By stratifying participants according to autistic traits and cognitive abilities, we identified three clusters with different load of both rare and common variants, particularly in genes associated with synaptic and chromatin remodeling functions. Correspondingly, each cluster exhibits different anatomical and connectivity patterns in brain regions previously linked to sensory and social information processing. This resource is available to support research into the complex connections between genetics, brain structure/function, and autism.

## Main

Autism is currently defined by persistent challenges in social communication and interaction, sensory processing, as well as restricted and stereotyped patterns of behavior, interests, or activities. Our understanding of autism etiology derives from various scientific fields and from research studies comparing a group of autistic people to neurotypical (NT) people ^1^. Genetics plays a crucial role in autism, with an estimated heritability to be around 80% ^2–4^. This influence arises from various genetic factors, including single *de novo* or ultra-rare variants and common variants found in the general population^5,6^. The genetic variants associated with autism can influence early brain development through multiple biological pathways, including the modification of chromatin remodeling complexes that regulate gene transcription by opening or closing genomic regions transcription ^7^. As neuronal circuits mature, genetic variants can also impact activity-dependent signaling and strength of synaptic connections ^8^. Neuroimaging studies have shown differences in brain anatomy/connectivity, particularly in regions involved in social perception, language, or sensory processing ^9–12^. Autism can be seen as an "emergent" condition shaped by the individual genetic profiles and the environment over time ^13^.

Most of this knowledge comes from studies showing results obtained at a group level, but the fact remains that what is relevant for one autistic individual in terms of etiology and support needs might be completely irrelevant for another ^1^. The conceptualization of the heterogeneity of autism has evolved over time. Wing and Gould were instrumental in defining autism as a spectrum ^14^, while Coleman, Fernell and Gillberg later proposed the term "autisms+" to reflect the notion of distinct etiological subgroups and the high prevalence of co-occurring medical conditions, such as intellectual developmental disorder (IDD), attention deficit hyperactivity disorder (ADHD) or epilepsy ^15^. Recently, the Lancet Commission proposed the term "profound autism" to describe the subset of people who are none or minimally verbal with significant intellectual disability, and requiring 24/7 care ^16^.

Understanding autism diversity should help to elucidate the underlying biological mechanisms involved in its physiology and allow the development of more efficient personalized interventions. Previous data-driven stratification using genetic^3^, neuroimaging ^12^, or clinical data ^6^ have identified on average 2-4 more homogeneous subgroups of autistic people, but this subgrouping depends mainly on the methodology and the population included in these studies (see **Supplementary Table 11** for a review). Studies typically focus on either genetics, neuroimaging or clinics, limiting our understanding of how the subgroups differ in genetic architecture, brain structure/function, and behavior.

To address the need for integrative approaches across modalities, we combined data from the EU-AIMS/AIMS-2-Trials Longitudinal European Autism Project (LEAP) ^17,18^ and the Innovation for Autism and Neurodevelopmental Disorders (InovAND) cohort. By clustering clinical data, we identified three subgroups within the autism spectrum showing differences in the nature of the *de novo*/rare and common variants mutations as well as in their pattern of brain anatomy/function. This integrated approach not only enhances our understanding of autism’s heterogeneity but also paves the way for more targeted and effective interventions, ultimately improving the quality of life for people on the autism spectrum.

## Results

### Overview of the LEAP-InovAND dataset

LEAP and InovAND are two distinct studies that recruited participants in different centers across Europe (**Fig. 1a**, **Supplementary Fig. 1**). We collected demographic and clinical data similarly to the Simons Foundation Powering Autism Research for Knowledge (SPARK) ^19, 20^ and Autism Brain Imaging Data Exchange (ABIDE), but our dataset includes a subset of people with clinical, genetic, neuroimaging data and bio- samples (**Fig. 1b**). The combined dataset encompasses 5,549 people (*i.e.,* 2,374 families) including 2,061 diagnosed with autism (DSM criteria) using gold standard (ADOS/ADI-R) instruments (**Fig. 1b**; **Supplementary Fig. 2**). The LEAP dataset includes 493 autistic people, 697 relatives (mostly parents) and 466 neurotypical (NT) people (**Supplementary Table 1**, **Supplementary Fig. 3**). The InovAND dataset includes 1,593 autistic people, 1,854 relatives (nuclear and extended family members) and 388 NT people (**Supplementary Table 2**, **Supplementary Fig. 3**). The clinical data collected by LEAP and InovAND show several similarities (**Supplementary Fig. 3**), such as comparable measured IQ scores among NT people and similar Vineland Adaptive Behavior Scales (VABS) scores among autistic people (**Supplementary Table 3** and **Supplementary Fig. 3**). However, regarding sex ratio, LEAP recruited a higher proportion of female autistic probands compared to InovAND, whereas the opposite pattern was observed among NT people (**Fig. 1c**, **Supplementary Tables 1 and 2**). Furthermore, autistic people from InovAND exhibited higher SRS-2 total t- scores (Mann-Whitney U test (MWU), P_MWU_ = 1.62 x 10^-5^) and lower full scale IQ scores (P_MWU_ = 4.36 x 10^-35^) than those from LEAP. We also collected data on parents and undiagnosed siblings, enriching the dataset for familial comparisons. Finally, 62 people with IDD who do not meet diagnostic criteria for autism were enrolled (36 for LEAP and 26 InovAND). All together the LEAP-InovAND dataset includes a fair representation of the whole continuum of autism in terms of autistic traits, cognition, gender, and age range (**Fig. 1e**).

**Figure 1.**
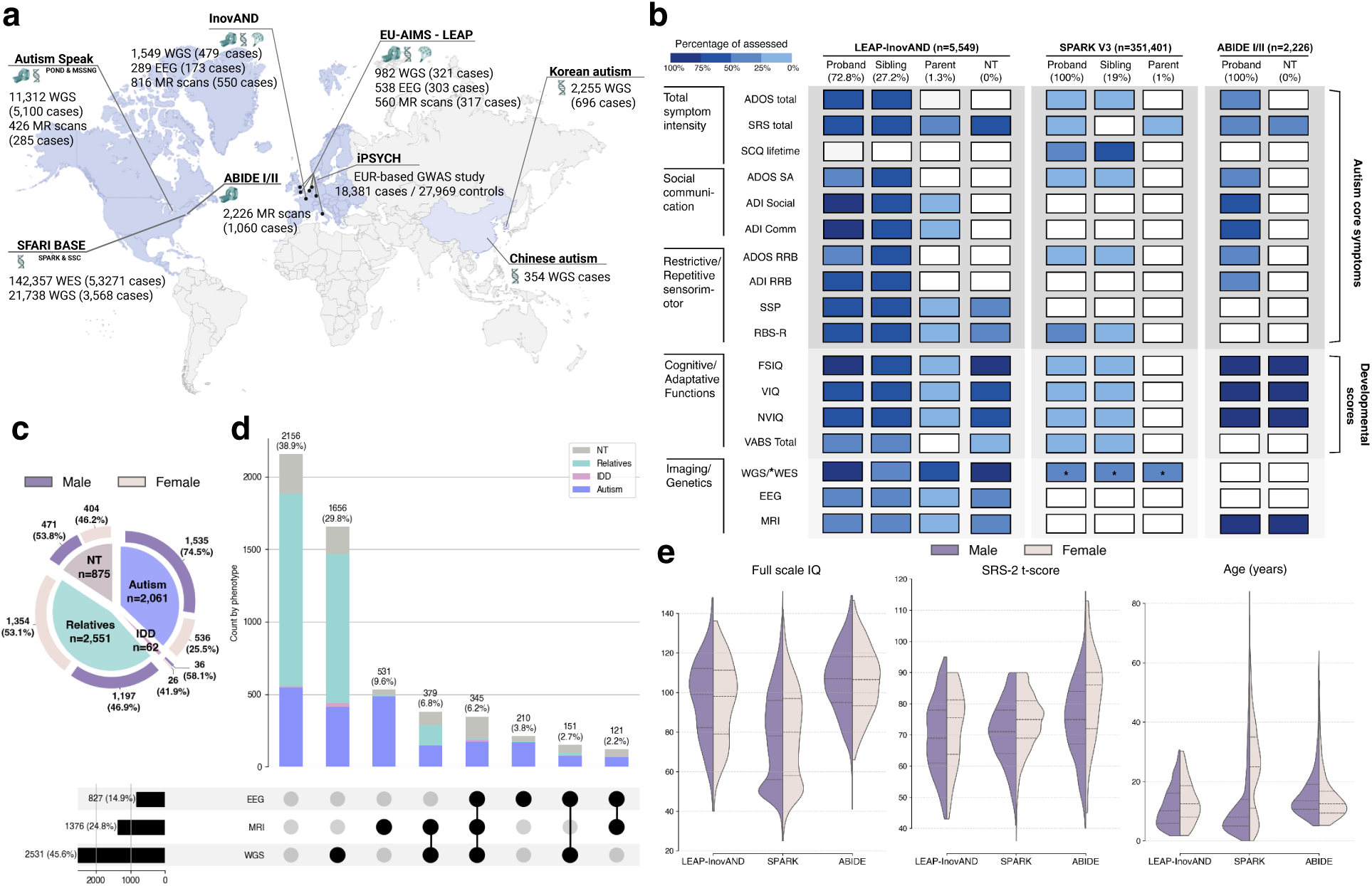
Overview of LEAP-InovAND cohort. **a.** Worldwide distribution of the major autism cohorts with >300 people with WGS or MRI data. For each dataset, sample sizes are indicated for WGS, and where available, MRI and EEG data, along with the number of autistic people. **b.** Percentage of assessed clinical variables across the LEAP-InovAND cohort, stratified by participant group: proband (autistic people; percentage shown below label), siblings, parents, and neurotypical people (NT). Clinical domains include core autism symptoms, developmental scores, and availability of imaging and genetics. The proband is the person in a family who is first included in the cohort. **c.** Composition of the full study samples (N = 5,549), represented as a nested pie chart showing the proportion of autistic and IDD people, NT, and undiagnosed relatives. Outer rings denote sex distribution (male *vs.* female). **d**. UpSet plot illustrating the overlap in available multimodal data (WGS, MRI, EEG), stratified by group (autism, IDD, NT, relatives). **e.** Distribution of full-scale IQ, Social Responsiveness Scale 2 (SRS-2) total t-scores, and age across our cohort, SPARK, and ABIDE datasets. Fig. inspired by Kim *et al*. ^66^.

### Genetic profile of the LEAP-InovAND participants

Following quality control and data harmonization procedures (see **Online Methods**), we analyzed whole genome sequencing (WGS) data through single nucleotide variants (SNV), small insertions/deletions (indel) and structural variants (SV) such as copy-number variants (CNV). The dataset contains WGS data for 2,531 people, including 800 with autism and their 1,210 relatives, 41 people with IDD only, and 480 NT people.

#### De novo variants

In 377 families with parental WGS data available, we could identify on average 77 *de novo* SNV (range 37-126) per participant. Consistent with previous studies ^21^, we observed a positive correlation between paternal age and the number of *de novo* SNV, with no significant excess in autistic people compared to their siblings (**Supplementary Fig. 9**). However, when focusing on predicted deleterious *de novo* variants in genes intolerant to protein-truncating variants (hereafter called “constrained genes” see **Online Methods**), autistic people were significantly more likely to carry such deleterious variants compared to their siblings (**Fig. 2a**; autism = 14.9%, undiagnosed siblings/NT = 3.9%, OR = 4.25, Fisher’s exact test false discovery rate (FDR) corrected (Fisher FDR) P_Fisher FDR_ = 8.1 x 10^-5^). The enrichment was even higher for variants in high-confidence neurodevelopmental genes (HCNDD; autism = 11.50%, undiagnosed siblings/NT = 1.14%, OR = 11.30; P_Fisher FDR_ = 7.0 x 10^-6^), for autism-linked list of genes (*e.g.*, SFARI1/SPARK lists; autism = 8.89%, undiagnosed siblings/NT=0.57%, OR = 16.94, P_Fisher FDR_ = 3.4 x 10^-5^), and for genes with definitive and strong EAGLE^22^ scores, a gene-level metric for autism gene evidence (autism = 4.85%, undiagnosed siblings/NT = 0%, OR = inf). Notably, genes involved in synaptic function (SynGO^23^) and/or chromatin remodeling/transcription (ChromEpiTF), two biological pathways previously implicated in autism ^24^, were enriched in *de novo* variants (SynGO: autism = 8.89%, undiagnosed siblings/NT = 0.57%, OR = 16.94, P_Fisher FDR_ = 3.4 x 10^-5^; ChromEpiTF: autism = 8.82%, undiagnosed siblings/NT = 1.70%, OR = 4.85, P_Fisher FDR_ = 0.0032). We confirmed all *de novo* variants in HCNDD genes using PCR and Sanger sequencing (**Supplementary Fig. 9**). Using the SPARK as a replication cohort, trends are similar for all the gene sets studied above (**Supplementary Fig. 15a**).

**Figure 2.**
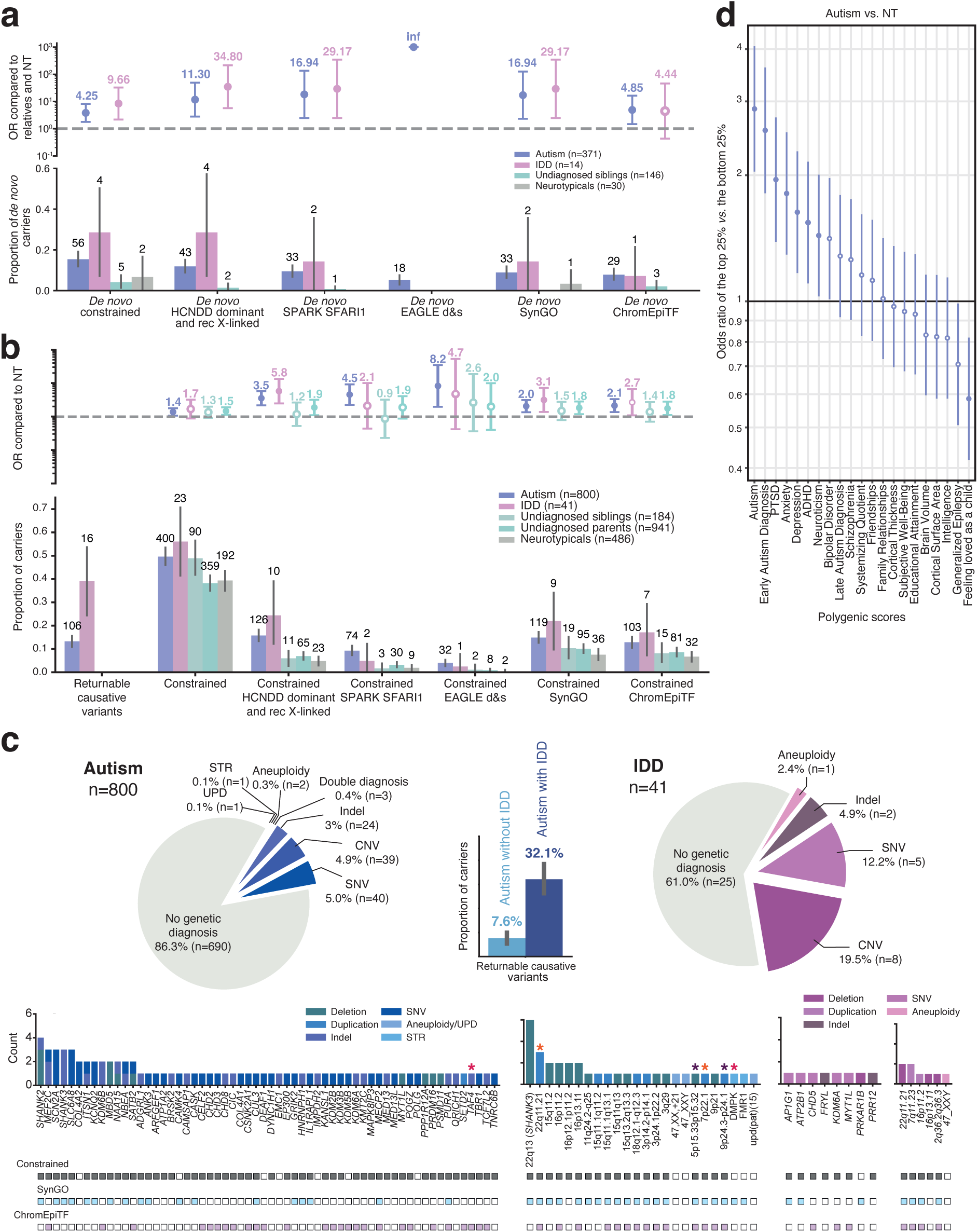
Genetic profile of the participant of the LEAP-InovAND cohort. **a.** Genetic study of *de novo* variants (deletions, loss of function and deleterious missenses) found in participants with both parents with WGS data. The bottom panel represents the proportion of *de novo* variant carriers according to the different gene lists used. The figure represents the number of carriers per population and per gene list. The top panel represents the odd ratio (OR) based on the proportions of the bottom panel. For *de novo* variations, OR (with CI95%) are calculated by comparing the frequency of *de novo* variants in autistic people/IDD to the pooled frequencies in undiagnosed siblings and NT. **b.** Analysis of returnable genetic diagnosis and rare deleterious variants, specifically deletions and predicted loss-of-function variants (hiURV), in predefined lists of constrained genes of interest. For this analysis, ORs (with CI95%) are computed and corrected (FDR, one-sided independent t-tests adjusted for multiple comparisons using Benjamini–Hochberg correction, difference is significant when the circle is full) compared to that of NT**. c.** Summary of genetic diagnostic findings in the autistic and IDD populations. The upper panel shows two pie charts illustrating the proportion of the different types of variants classified as diagnostic. Among autistic people with IDD, 4.7% carry variants affecting *SHANK3* or the 22q13 *locus*. These cases include 9 people with 8 distinct variants (8 distinct families). The lower panel displays the distribution of genes and loci with their respective counts, highlighting their overlap with constrained gene lists, SynGO, and ChromEpiTF datasets. Three cases of dual genetic diagnoses were identified; colored stars (in three distinct colors) indicate variants carried by the same person. **d.** PGS for different GWAS compared to NT, representing the OR for the top 25% versus the bottom 25% of the PGS distribution. The difference is significant when the circle is full. ADHD: attention-deficit/hyperactivity disorder, HCNDD: high confidence NDD, IDD: intellectual developmental disorder, NT: neurotypical, PGS: polygenic score, PTSD: post-traumatic stress disorder, STR: short tandem repeats, UPD: uniparental disomy.

#### High impact ultra-rare variants

the definition of the high impact ultra-rare variants (hiURV) is summarized in **Fig. 2b and Supplementary Fig. 6**. Briefly, these variants can be *de novo* or inherited, but they must have minor allele frequencies (MAF) of less than 0.1% in the general population and be predicted to have a deleterious impact on the function of the protein (see **Online Methods)**. Here, we focused our analysis on loss-of-function (LoF) variations and deletions (DEL) within “constrained genes”. Compared to NT people, autistic people were more likely to carry at least one hiURV in HCNDD genes (**Fig. 2b**, autism = 15.57%, NT = 4.73%, OR = 3.53; P_Fisher FDR_ = 0.035), autism-linked genes (SPARK/SFARI1; autism = 9.15%, NT = 1.85%, OR = 4.54, P_Fisher FDR_ = 1.2 x 10^-5^) and EAGLE genes (autism = 3.96%, NT = 0.41%, OR = 8.23, P_Fisher FDR_ = 0.0018). Genes involved in synaptic function and/or chromatin remodeling/transcription were also enriched for hiURV (SynGO: autism = 14,71%, NT = 7.41%, OR = 2.05, P_Fisher FDR_ = 0.0023; ChromEpiTF: autism = 12.73%, NT = 6.58%, OR = 2.12, P_Fisher FDR_ = 0.0023). Similar results are obtained if deleterious missense variants are added and if genes less constrained are included (**Supplementary Fig. 11f**). Using SPARK as a replication cohort, similar trends are observed (**Supplementary Fig. 15b**).

#### Genetic diagnostic yield

Returnable clinically relevant variants are filtered through a very strict pipeline (see **Supplementary Fig. 5**) that retains only functional variants in genes/*loci*/aneuploidies previously known as causes of autism/neurodevelopmental disorders (NDD). We found these variants in 13.7% of the autistic people (n=110/800) and in 39% of the people with IDD only (n=16/41) (**Fig. 2c**). In autistic people, co-occurring IDD raises the diagnostic yield to 32.1%, while its absence lowers it to 7.6% (**Supplementary Fig. 12b**). *SHANK3* is the most frequently implicated gene in the cohort, encompassing eight distinct families (in one family, two sons carried a *SHANK3* LoF variant transmitted from a mother mosaic for this variant). These nine people carrying *SHANK3* variants accounted for 4.7% of autistic people with IDD (n = 190). *SHANK3* was followed in frequency by recurrent CNV involving the 15q and 16p loci (n = 5 each) and by *SHANK2* (n = 4). (**Fig. 2c**).

#### Polygenic Scores (PGS)

Common variants such as single nucleotide polymorphism (SNP) also contribute to autism and to a range of other traits and neuropsychiatric conditions, including post-traumatic stress disorder (PTSD), ADHD, epilepsy, cognitive performance (as assessed by IQ scores), and various dimensions of subjective well-being (**Supplementary Figs. 16 and 17**). Although each SNP exerts only a small effect on a person’s phenotype, their accumulation in the person can be calculated through PGS. PGS were calculated for each participant in the LEAP- InovAND cohort across multiple traits (see **Online Method**). As expected, autism PGS showed the strongest difference between autistic people and NT (autism *vs.* NT; OR = 2.88, P_MWU_ = 1.5 x 10^-9^), followed by PTSD (OR = 1.95, P_MWU_ = 0.0002), anxiety (OR = 1.81, P_MWU_ = 0.0007), depression (OR = 1.63, P_MWU_ = 0.0052), ADHD (OR = 1.54, P_MWU_ = 0.01), and neuroticism (OR = 1.44, P_MWU_ = 0.04) (**Fig. 2d**). Analyses restricted to people of European ancestry yielded comparable results (**Supplementary Fig. 15a**). Replication analyses conducted using the independent SPARK cohort (**Supplementary Fig. 16a**) corroborated these findings. In the LEAP-InovAND and the SPARK cohorts, the “felt loved as a child” PGS displayed the lowest odds ratio among autistic people (LEAP-InovAND: autism *vs.* NT OR = 0.54, P_MWU_ = 0.0012; SPARK: autism *vs.* undiagnosed siblings OR = 0.86, P_MWU_ = 0.0089). The concordance of the PGS odds ratio across both cohorts (**Supplementary Fig. 16b**) underscores the similar contribution of common variants in autistic people from independent samples.

### Brain imaging profile of the LEAP-InovAND participants

Magnetic resonance imaging (MRI) and electroencephalography (EEG) data were collected across 6 centers and 8 MRI scanners (6 from the LEAP study and 2 from InovAND). Following quality control, standard preprocessing and data harmonization procedures (see **Online Methods**), we evaluated diagnostic differences in cortical morphology, functional connectivity, and electrophysiological features.

#### Structural MRI (sMRI) analysis

The *cortex* was parcellated into 62 regions of interest (ROI) using the Desikan–Killiany (DK) atlas ^25^ (see **Online Methods**). Compared to NT (n = 349) autistic people (n = 776) exhibited a selective pattern of increased cortical thickness, primarily in temporo-parietal and occipital regions (**Fig. 3a**). The largest effect was observed in the left superior temporal gyrus (STG; t = 5.37, Bonferroni-adjusted p-value (BON) P_BON_ = 7.6 x 10^-6^), with additional increases were observed in the right inferior parietal lobe (t = 3.54, P_BON_ = 0.029), right lateral occipital cortex (t = 3.42, P_BON_ = 0.046), left rostral middle frontal (t = 3.67, P_BON_ = 0.017) and right superior temporal (t = 3.6, P_BON_ = 0.02). No surface area differences survived correction for multiple comparisons (**Fig. 3b**). Among subcortical volumes (**Fig. 3c**), only the right lateral ventricles were enlarged in the autism group (t = 3.26, P_BON_ = 0.032).

**Figure 3.**
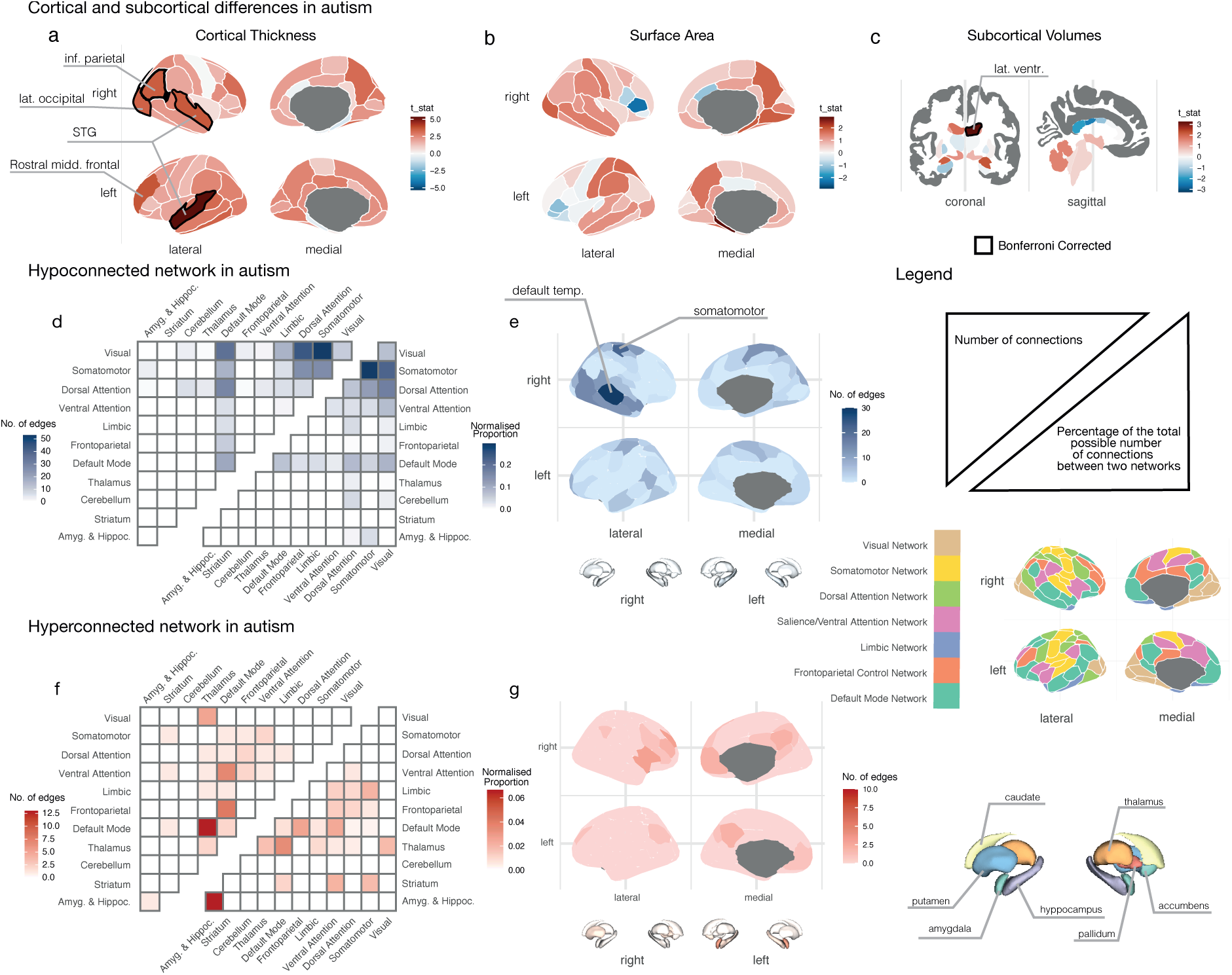
Anatomical and functional cortical and subcortical differences in autistic people compared to neurotypical people (NT). Cortical and subcortical anatomical differences assessed via vertex-wise t-tests on cortical thickness (**a**.), surface area (**b.**) and subcortical volume (**c.**) between autistic (n = 776) and NT people (n = 349). Brain maps show t-statistics, thresholded using Bonferroni correction (q < 0.05), projected onto the Desikan-Killiany (cortical) and aseg (subcortical) atlases. Functional connectivity differences across cortical and subcortical regions using the Schaefer-100 parcellation and CIT168 subcortical atlas between autistic (n = 396) and NT (n = 296) people. **d-g.** Connectivity matrix displaying the number of significantly altered connections (edges) between and within functional networks (FDR-corrected). The lower triangle of the matrix shows the proportion of increased connections falling within and between each of the networks, normalized by the total number of possible connections within or between the corresponding networks; the upper triangle shows the raw count of edges for hypoconnectivity (**d.**) and hyperconnectivity (**f.**). Cortical and subcortical maps showing the degree of each region significant in hypoconnectivity (**e.**) and hyperconnectivity (**g.**), colored by the number of altered edges per region. Note: STG: Superior Temporal Gyrus; inf.: Inferior; Mid.: Middle; Temp.: Temporal; Lat. Ventr.: Lateral Ventricle; DMN: Default Mode Network.

#### Resting state functional MRI (rsfMRI) analysis

Functional connectivity was computed between 115 brain regions for 396 autistic people 296 NT people. We then averaged the voxel-wise time series within each parcel and computed Pearson’s correlation between the time course of all pairs of parcels for each participant as described in ^26^(see **Online Methods**). Group differences were assessed at the edge level, and only connections surviving FDR correction (P_FDR_ < 0.05) were retained for subsequent analyses. To summarize effects at the network level, we quantified edges showing hypoconnectivity or hyperconnectivity and report both the raw number of significant edges (N_raw_) and the normalized proportion of significant edges relative to all possible connections within/between each network pair (N_norm_). Group contrasts revealed extensive underconnectivity in autism. The highest fraction of hypoconnected edges occurred between somatomotor and visual networks (N_norm_ = 0.22; *e.g.*, 22% of possible edges) (**Fig. 3d-e**), followed by visual–dorsal attention (N_norm_ = 0.17), and somatomotor-dorsal attention (N_norm_ = 0.12). Default mode connections to sensory– attentional systems were also reduced, notably default mode-visual (N_raw_ = 36, N_norm_ = 0.088) and default mode-dorsal attention (N_raw_ = 29, N_norm_ = 0.081). Within network hypoconnectivity peaked in somatomotor (N_raw_ = 27, N_norm_ = 0.29), with smaller within network reductions in visual (N_raw_ = 10, N_norm_ = 0.074) and default mode (N_raw_ = 19, N_norm_ = 0.069). In contrast, hyperconnectivity (**Fig. 3f-g**) was sparse and centered on the default mode network’s (DMN) coupling with subcortical and control/attention systems (<4% of possible edges), including default mode-thalamus, default mode-frontoparietal, and default mode-ventral attention. Few within network hyperconnectivity effects were observed.

#### Resting state EEG (rsEEG) analysis

EEG spectral features were compared between 406 autistic people and 217 NT. *Alpha* peak frequency (6-13 Hz) measured in occipital regions was comparable between autistic (n = 406) and NT (n = 217) participants (**Supplementary Fig. 20**), and spectral power across the five canonical frequency bands (*delta*, *theta*, *alpha*, *beta*, and *gamma*) did not differ significantly. This absence of robust EEG differences, at the group level, aligns with recent population-scale reports^27^.

### Clustering analysis on clinical profiles of people

As an example of the analytical potential of the LEAP-InovAND dataset, we conducted a subgrouping analysis of 1,023 unrelated people (609 autistic people, 31 people with IDD, and 383 NT) using the Social Responsiveness Scale total (SRS-2 total t-score) and the measured IQ scores (see **Supplementary Section 6**). The SRS-2 t-score is a widely used, standardized questionnaire that quantifies autism-related traits dimensionally, allowing for more nuanced analyses beyond binary diagnostic categories ^28^. Feature selection for subgrouping balanced multiple considerations: principal component analysis on all available clinical measures showed that IQ and SRS-2 total t-scores were among the top contributors to the first two principal components (capturing 78.6% of the total variance, **Supplementary Fig. 21**), while also maximizing sample size. This 2-variable combination retained 1,023 participants compared to 511 participants when including VABS composite scores (See **Supplementary Section 6**, **Supplementary Table 12**). Hierarchical clustering based on measured IQ and SRS-2 total t-score, revealed an optimal three-cluster solution as determined by comprehensive fit indices analysis (*e.g.*, silhouette, variance ratio score, gap statistic, pseudo-F) (**Fig. 4a**; **Supplementary Table 13**; **Supplementary Fig. 21 and 22**). Cluster 1 (C1) includes 451 people; cluster 2 (C2) comprises 379 people; and cluster 3 (C3) consists of 193 people (**Fig. 4b**). These clusters demonstrate distinct profiles across clinical presentation, genetic architecture, and neuroimaging features, suggesting meaningful stratification within the autism spectrum.

**Figure 4.**
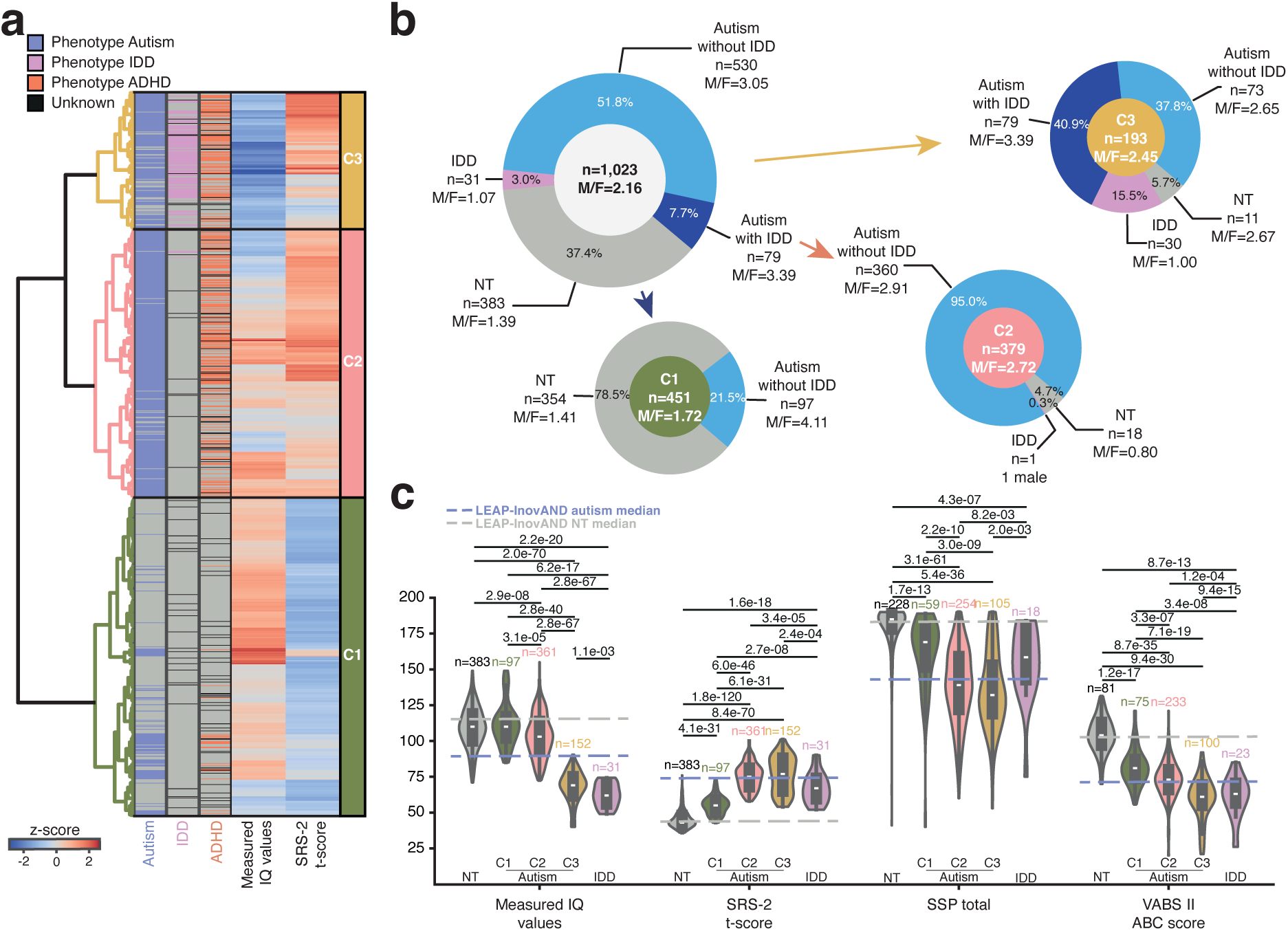
Hierarchical clustering of 1,023 people based on measured IQ values and SRS-2 t-score. **a.** Dendrogram generated using hierarchical clustering with Ward’s method on standardized behavioral measures, revealing three distinct clusters: C1 (n = 451), C2 (n = 379), and C3 (n = 193). These clusters represent the autism- derived subgroups only and do not include NT or IDD only people in downstream visualizations (*e.g.*, panel **c.**). **b.** Circle plots showing the count and proportion of people within each cluster presenting with specific categorical characteristics, including autism without IDD, autism with IDD, IDD, NT and sex ratio. **c.** Cluster-wise distributions of clinical scores including full-scale IQ, SRS 2 total t-score, short sensory profile (SSP) total score, and Vineland Adaptive Behavior Composite II (VABS-II) ABC score. Blue and grey dashed lines indicate the median scores of autistic people (with or without IDD) and neurotypical (NT) people in all the cohort, respectively. The presented statistics are Mann-Whitney-Wilcoxon test results. ADHD: attention-deficit/hyperactivity disorder, IDD: intellectual developmental disorder, NT: neurotypical.

#### Clinical profiles

Cluster C1 gathers people with medium-to-high IQ scores and low levels of autistic traits (*i.e.*, SRS-2 total t-scores below the cut-off for autism). This cluster includes primarily NT people (n = 354; 92.4% of all NT; 78.5% of people in C1), along with a subset of autistic people without IDD (n = 97; 15.9% of all autistic people; 21.5% of C1). Cluster C2 includes people with medium-to-high IQ and elevated SRS-2 total t-scores and is composed almost entirely of autistic people without IDD (n = 360 58.9% of all autistic people; 95.0% of C2). Cluster C3 includes people with low IQ and high SRS-2 scores, with most of the people in this cluster are autistic (n = 152; 25.0% of all autistic people; 78.7% of C3), with approximately half also meeting criteria for IDD (n = 79). Notably, almost all people with a clinical diagnosis of IDD only are also assigned to C3 (n = 30; 96.8% of all people with IDD; 15.5% of C3). Beyond IQ and autistic traits, we observed clinical differences in the autistic people across clusters, particularly with an increased prevalence of clinical diagnosis of ADHD in C2 and C3 compared to C1 (C2: P_Fisher_ = 1.7 x 10^-17^; C3: P_Fisher_ = 1.3 x 10^-11^). Similarly, autistic people within C2 and C3 (**Fig. 4c**) presented with higher sensory atypicality (measured by the short sensory profile, SSP) compared to those within C1 (C2 *vs.* C1: P_MWU_ = 2.2 x 10^-10^; C3 *vs.* C1: P_MWU_ = 3.0 x 10^-9^). Additionally, autistic people within C2 and C3 displayed lower adaptive functioning (assessed via the VABS) compared to those within C1 (C2 *vs.* C1: P_MWU_ = 3.3 x 10^-7^, C3 *vs.* C1: P_MWU_ = 7.1 x 10^-19^). Developmental milestones analysis revealed that autistic people in all clusters showed delayed acquisition of early motor and language milestones (e.g., age at onset of walking, first words, first phrases) compared to NT people, but with a gradient of severity, with the most pronounced delays observed for autistic people from C3 (see **Supplementary Fig. 23** for detailed statistics). Interestingly, no significant sex differences were observed between the autistic people in C1, C2 and C3 (χ² = 1.5, P_Chi squared_ = 0.47).

#### Genetic profiles

We identified significant differences in the genetic architecture of the participants across the three clusters. In total, 22.7% of autistic people in C3 received a genetic diagnosis with a returnable variant linked to autism/NDD, versus 6.1% of autistic people in C2 and 7.5% of autistic people in C1 (C1 *vs.* C2: P_Fisher_ = 0.012; C1 *vs.* C3: P_Fisher_ = 1.3 x 10^-5^) (**Fig. 5a**). Regarding rare variants, for the constrained HCNDD gene list, all clusters of autistic people exhibit a significantly higher variant burden relative to the NT (C1: OR = 4.4, P_Fisher FDR_ = 0.0056; C2: OR = 3.9, P_Fisher FDR_ = 2.7 x 10^-4^; C3: OR = 7.7, P_Fisher FDR_ = 1.7 x 10^-7^; IDD: OR = 10, P_Fisher FDR_ = 0.0013). The same is true for the constrained SPARK/SFARI1 gene list (C1: OR = 4.6, P_Fisher FDR_ = 0.0027; C2: OR = 3.4, P_Fisher-FDR_ = 0.023; C3: OR = 6.8, P_Fisher FDR_ = 0.0003). Interestingly, only autistic people in cluster C3 display a significant association for the constrained EAGLE definitive and strong gene set, as well as for the SynGO and ChromEpiTF gene lists (EAGLE d&s: OR = 6.7, P_Fisher FDR_ = 0.037; SynGO: OR = 2.7, P_Fisher FDR_ = 0.0064; ChromEpiTF: OR = 3.2, P_Fisher FDR_ = 0.048). Focusing on the cell types of the prefrontal cortex and expression patterns during neurodevelopment, only autistic people from cluster C3 showed significant OR compared with NT, mostly for genes expressed early during brain development such as the medial ganglionic eminences (MGE) inhibitory interneurons and the principal excitatory neurons (**Supplementary Fig. 13**).

**Figure 5.**
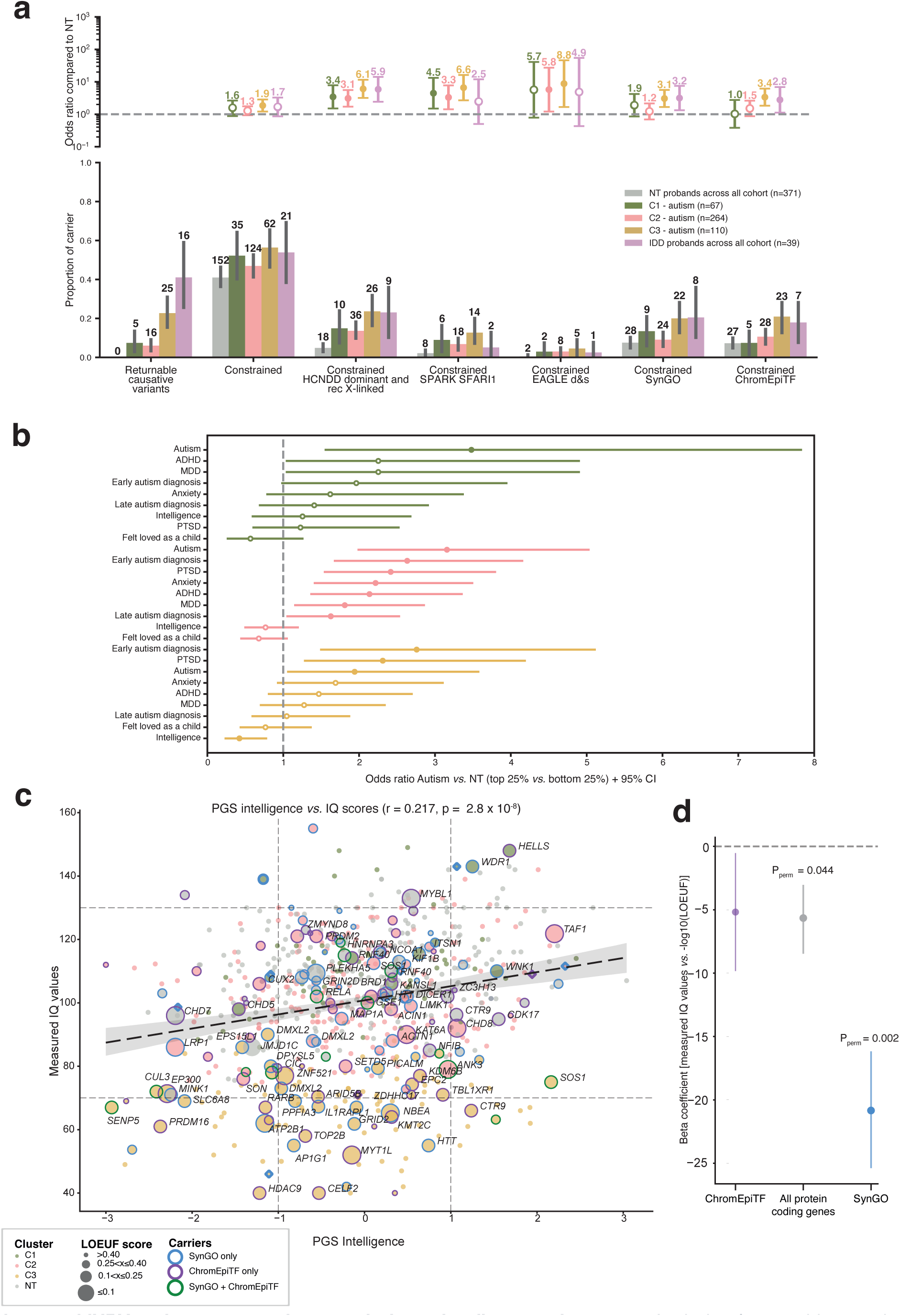
hiURV and common variants analysis of the different clusters. **a.** Analysis of returnable genetic diagnosis and rare deleterious variants (hiURV), specifically deletions and predicted loss-of-function variants, in predefined lists of constrained genes of interest. The figure shows the number of deleterious variant carriers, stratified by cluster and gene list. Results are displayed as counts with corresponding 95% CIs. ORs are presented with their 95% CIs; filled circles denote results that remain significant after FDR correction using the Benjamini– Hochberg method. **b.** PGS for different GWAS compared to NT, representing the OR for the top 25% versus the bottom 25% of the PGS distribution. The difference is significant when the circle is full. **c.** A correlation plot illustrating the relationship between measured IQ, the PGS for intelligence and the hiURV found across people in different clusters. The size of each dot represents the lowest LOEUF score, and people within SynGO, ChromEpiTF and both gene lists. The different gene lists are highlighted with outlines. **d.** Beta coefficients and standard errors from linear regression of measured IQ on −log10(LOEUF) for protein-coding genes, ChromEpiTF, and SynGO. For each gene set, permutation p-values are shown for Pearson correlation between constrained gene scores [- log10(LOEUF)] and measured IQ. Statistical significance was assessed using 1,000 permutation iterations with gene-set–specific null hypotheses: random reassignment of gene scores for protein-coding genes. HCNDD: high confidence NDD genes (only dominant inheritances and recessive X-linked in male), SFARI1/SPARK: union of the SFARI1 and SPARK returnable genes, SynGO and ChromEpiTF are described in **Online Methods**. ADHD: attention-deficit/hyperactivity disorder, EAGLE d&s: EAGLE definitive and strong, HCNDD: high confidence NDD, IDD: intellectual developmental disorder, NT: neurotypical, PGS: polygenic score, PTSD: post-traumatic stress disorder.

Compared to NT, PGS analysis revealed increased OR for autism, ADHD, and major depressive disorder (MDD/depression) in autistic people from clusters C1 (autism OR = 3.2, P_MWU_ = 0.0063; ADHD OR = 2.5, P_MWU_ = 0.035; MDD OR = 2.8, P_MWU_ = 0.02) and C2 (autism OR = 2.6, P_MWU_ = 0.0002; ADHD OR = 2.4, P_MWU_ = 0.0007; MDD OR = 2.0, P_MWU_ = 0.0052), but not in C3. Notably, as mentioned, the autism PGS in C3 did not differ significantly between autistic people and NT (autism OR = 1.8, P_MWU_ = 0.1). Compared to NT people, elevated PGS for PTSD was observed in both C2 (PTSD OR = 3.4, P_MWU_ = 1.0 x 10^-6^) and C3 (PTSD OR = 2.7, P_MWU_ = 0.0021), with anxiety PGS specifically increased in C2 (anxiety OR = 2.3, P_MWU_ = 0.001). Finally, the PGS related to intelligence was significantly reduced in autistic people from C3 compared to NT (intelligence OR = 0.4, P_MWU_ = 0.0041), an effect not seen in C1 or C2 (**Fig. 5b**). In the overall cluster population, there was a positive correlation between the PGS for intelligence and the measured IQ score (r = 0.217, P_Pearson_ = 1.0 x 10^-6^; **Fig. 5c**). Notably, several autistic people in cluster C3 carried highly deleterious rare variants in SynGO and ChromEpiTF genes along with low intelligence PGS scores (**Supplementary Fig. 24**), suggesting cumulative effects of rare and common variants in this subgroup. Finally, we examined the associations between IQ scores and rare variants in genes based on their level of intolerance to LoF variants (LOEUF scores, **Fig. 5d**). A negative correlation was observed for all protein-coding genes (r = -0.08, Pperm = 0.044), but the strongest association was found between higher constraints within SynGO genes and lower IQ (r = -0.24, Pperm = 0.0002).

#### Brain structural/functional profiles of people based on clustering

*sMRI:* Autistic people from cluster C2 showed greater cortical thickness than NT in the left STG (t = 3.89, P_FDR_ = 0.008) (**Fig. 6a**). In contrast, autistic people in C3 exhibited thinner cortex than NT in the right precentral gyrus (t = -4.06, P_FDR_ = 0.004), right temporal pole (t = -3.68, P_FDR_ = 0.008), and left precentral gyrus (t = -3.19, P_FDR_ = 0.026). Cluster C3 also showed a thicker cortex than NT in the right pericalcarine visual cortex (t = 3.63, P_FDR_ = 0.008) and left pericalcarine visual cortex (t = 3.13, P_FDR_ = 0.026). Autistic people in C3 showed reduced surface area compared to NT in the left pars orbitalis (t = -3.39, P_unc_ = 0.001, P_FDR_ = 0.06). For subcortical volumes, Autistic people in C1 showed larger right lateral ventricle volume compared to NT (t = 3.64, P_FDR_ = 0.009).

**Figure 6.**
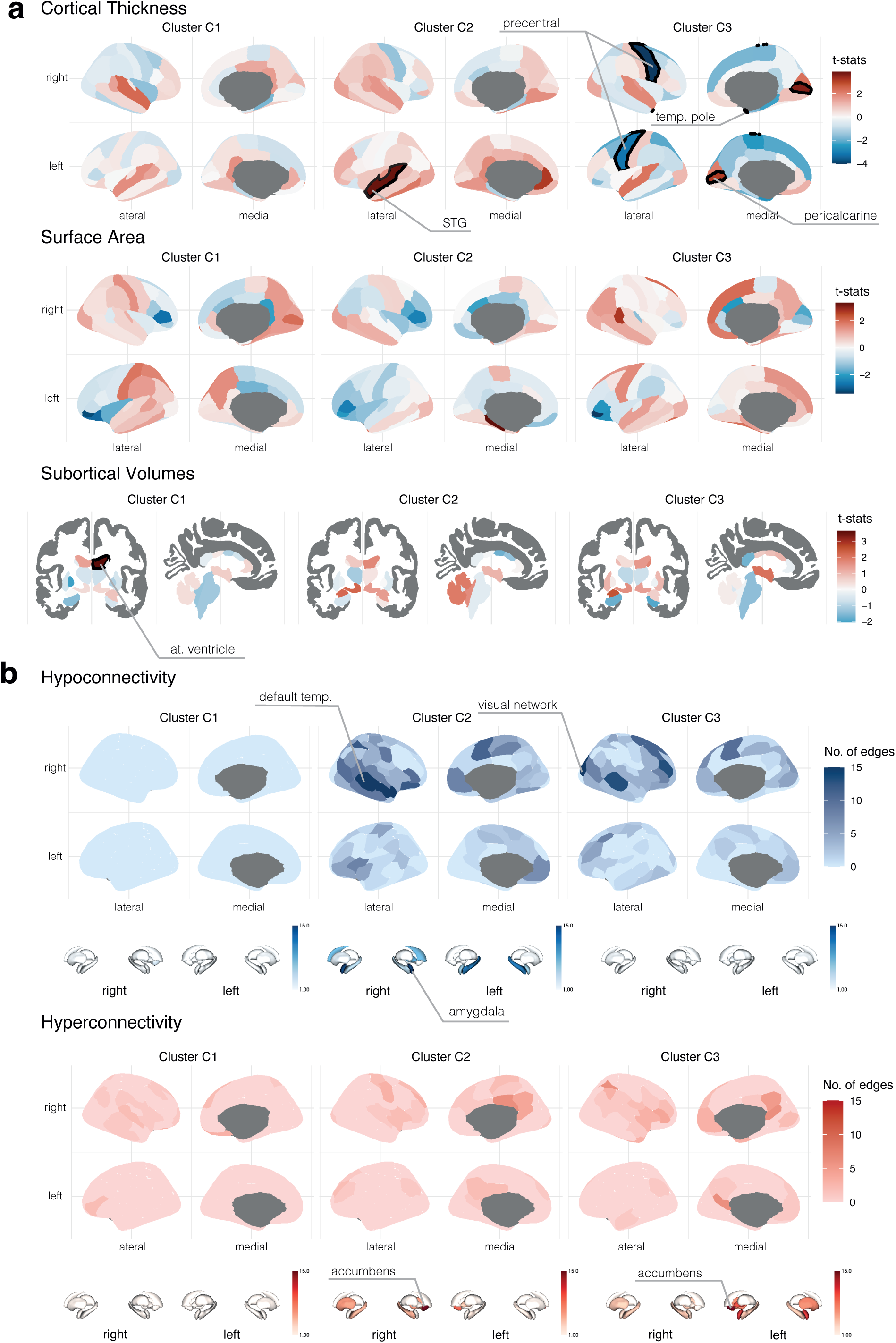
Structural and functional MRI differences between autism in the clusters and neurotypical. **a.** Cortical and subcortical anatomical differences between clusters and neurotypicals (C1: n = 303; C2: n = 208; C3: n = 82) were assessed using vertex-wise t-tests on cortical thickness (left) and volumetric comparisons of subcortical structures (right). t-statistic maps are displayed on the Desikan–Killiany (cortical) and aseg (subcortical) atlases, thresholded using false discovery rate (FDR) correction (q < 0.05). **b.** Functional connectivity differences between clusters and neurotypicals were analyzed using the Schaefer-100 parcellation and CIT168 subcortical atlas. Brain maps show significant alterations in connectivity strength across regions. Regions exhibiting significant hypoconnectivity (top) and hyperconnectivity (bottom) are colored according to the number of significantly decreased or increased connections per region (unc. p < 0.05). Note: Lat.: Lateral.; Temp.: Temporal; STG: Superior Temporal Gyrus.

*rsfMRI:* Autistic people from cluster C2 showed more inter-network hypoconnectivity than NT in the default mode and visual networks (N_raw_ = 48, N_norm_ = 0.12), followed by dorsal attention with default mode (N_raw_ = 30, N_norm_ = 0.08) and ventral attention with somatomotor (N_raw_ = 32, N_norm_ = 0.19) (**Fig. 6b**). Hyperconnectivity in autistic people in C2 *vs.* NT was widespread, notably involving thalamo-cortical and limbic–cortical coupling: thalamus with default mode (N_raw_ = 27, N_norm_ = 0.07), default mode with ventral attention (N_raw_ = 13, N_norm_ = 0.05), limbic with visual (N_raw_ =15, N_norm_ = 0.13), and frontoparietal with somatomotor (N_raw_ = 11, N_norm_ = 0.06). For autistic people in C3 *vs.* NT, hypoconnectivity concentrated between default mode and visual (N_raw_ = 31, N_norm_ = 0.08) and between dorsal attention and somatomotor (N_raw_ =13, N_norm_ = 0.06), while hyperconnectivity featured thalamus–default mode (N_raw_ = 24, N_norm_ = 0.06), default mode–frontoparietal (N_raw_ = 19, N_norm_ = 0.06). In contrast, autistic people in C1 *vs.* NT exhibited sparse effects overall; hypoconnectivity was limited (*e.g.,* thalamus–visual, N_raw_ = 5, N_norm_ = 0.02), and hyperconnectivity was modest, with small increases in default mode–dorsal attention (N_raw_ = 4, N_norm_ = 0.01) and default mode-somatomotor (N_raw_ = 5, N_norm_ = 0.015).

*rsEEG:* The examination of the autism clusters compared to NT people, revealed distinct EEG patterns. Autistic people in C3 (n = 51), showed significantly lower *alpha* peak frequency in occipital region compared to NT (t = -3.47, P_FDR_ = 0.0009) (**Supplementary Fig. 26**). In contrast, autistic people in C1 and C2 showed no significant differences from NT.

### Correlation between genes, brain structure/function and IQ

For all carriers of LoF variants, we examined if there was a link between higher constraint on the mutated gene (indicated by higher -log(LOEUF) score ^29^) and higher cortical thickness (**Fig. 7a-c**). We found two positive correlations: the left STG showed significantly increased thickness associated with higher constraint scores in ChromEpiTF factor genes (r = 0.215, P_perm_ = 0.02) and the left lateral occipital (r = 0.239, P_perm_ = 0.002). To showcase the diversity of clinical, genetic and neuroimaging data, we displayed the IQ score and the SRS-2 total t-score, highlighting one autistic person in each cluster, carrying predicted deleterious variants in either SynGO or ChromEpiTF genes (**Fig. 7d**).

**Figure 7.**
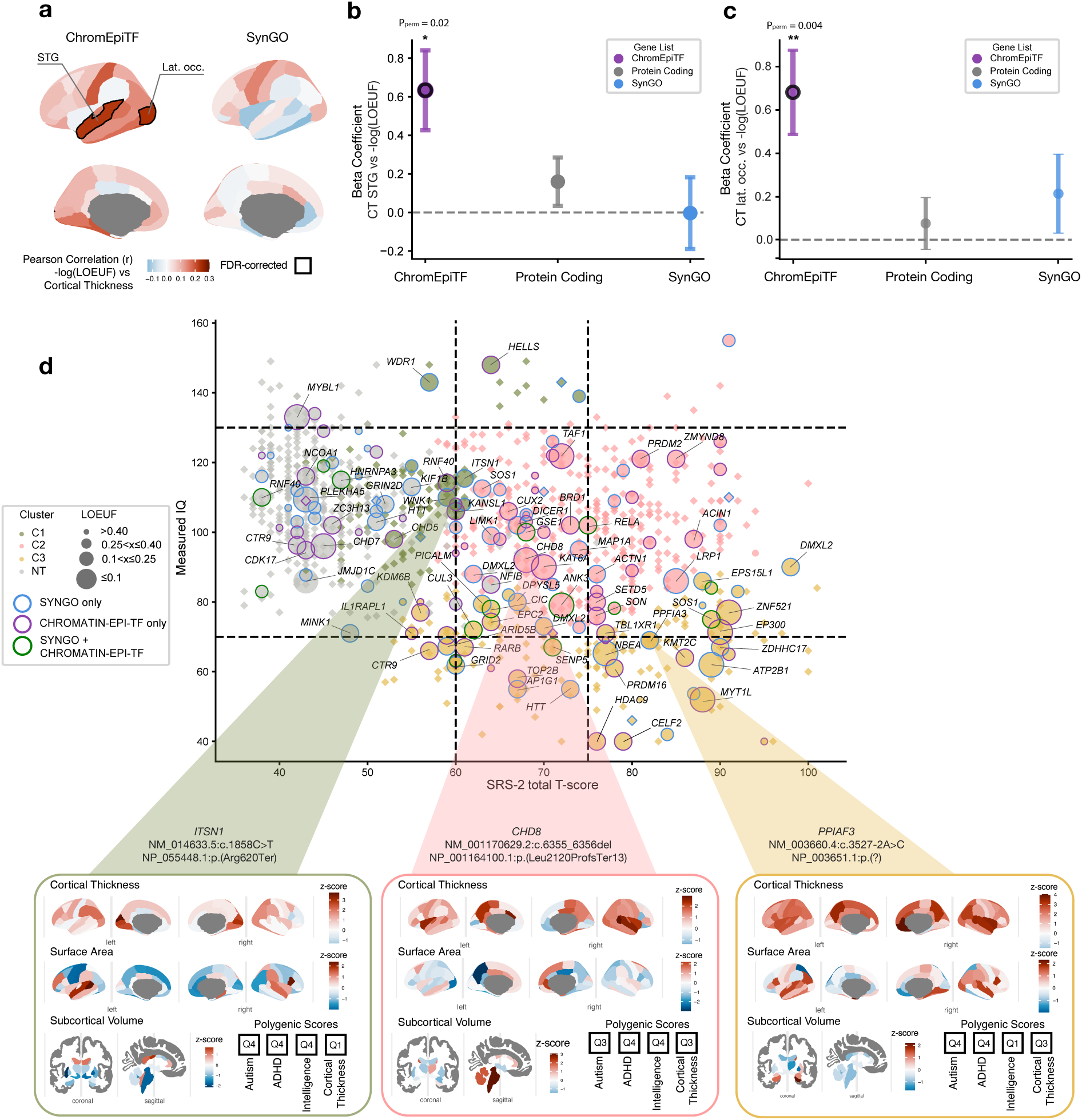
Relationship between genetic variants, brain anatomy and clinical features. **a.** Pearson correlation were computed between constrained gene scores (-log10LOEUF) from SynGO and ChromEpiTF and cortical thickness (CT) in the 68 Desikan–Killiany atlas. LOEUF is a population-based measure of the intolerance of each gene to LoF variants; higher −log₁₀(LOEUF) values indicate stronger constraint. **b**-**c.** Beta coefficients and standard errors from linear regression of CT on -log10(LOEUF) for protein-coding genes, ChromEpiTF (n = 204 carriers), and SynGO (n = 226 carriers). For each gene sets, permutation p-values are displayed. Pearson correlation between constrained gene scores [-log10(LOEUF)] and CT are illustrated for two representative regions: left superior temporal gyrus (STG) and left lateral occipital cortex (lat. occ.). Statistical significance was evaluated through permutation tests (1,000 iterations) with gene list-specific null hypotheses: random assignment for protein-coding genes. **d.** Scatter plot of measured IQ and SRS-2 total t-score colored by the clustering of the people. People with hiURV in SynGO, ChromEpiTF or both are highlighted with outlines. The size of each dot is proportional to the intolerance to LoF variants (a LOEUF value < 0.6 indicates high intolerance to LoF variants as described in GnomADv4.1). Dashed lines represent the standard clinical thresholds. A focus has been made on three different people (one in each cluster), all three carrying a hiURV in specific genes with their sMRI z-score values and 4 different PGS.

## Discussion

This study represents a major collaborative effort that brought together a diverse group of researchers and experts by experience from across Europe, including psychologists, psychiatrists, geneticists, neuroscientists, and, importantly, autistic people, relatives and caregivers (see **Supplementary Section 1 and Section 8** on participatory research and the frequently asked questions). This integrative framework has allowed us to investigate how genetic variation may shape brain structure, and in turn, how these brain differences relate to autistic traits and developmental outcomes.

In the first part of this study, we demonstrated that this new LEAP-InovAND dataset closely resembles previously established datasets in terms of clinical features, brain imaging, and genetics. This similarity confirms shared biological pathways while also highlighting differences in brain anatomy/function between autistic and NT people. In the second part, we found that group-level distinctions can be more precisely attributed to specific clinical subgroups.

For instance, previous work, including our own, has shown that the genetic architecture of autism involves a complex interplay of *de novo* variants, rare and common inherited variants ^2–6^. In the LEAP-InovAND dataset, we found that *de novo*/rare variants in constrained genes were significantly more frequent in autistic people compared to NT people. This enrichment was even more pronounced when variants disrupted coding sequences of genes previously implicated in autism/NDD.

These variants were enriched in autistic people with co-occurring cognitive challenges (**Supplementary Fig. 12**), although they were not exclusive to this subgroup. The returnable genetic variants in autistic people were enriched with genes previously associated with mild-to-moderate IDD (*e.g.*, *ADGRL1*, *ITSN1*, *KDM5B*, *QRICH1*, *CHD5*, *CHD8*), highlighting the overlap of autism with mild IDD. Interestingly, several of these genes have been challenging to implicate in NDD due to their wide clinical spectrum and incomplete penetrance (*ADGRL1*, *ITSN1*, *KDM5B*, *PSMD11*)^30–33^. One example is the gene coding for the presynaptic protein BSN, recently implicated in a wide spectrum of NDD ^34^, for which the only participant with a truncating variant in the LEAP-InovAND cohort was a 12-year-old NT female with some autistic traits according to her SRS-2 t-score at 57. Our cohort that includes phenotypic explorations of NT people provides a valuable resource to better understand some wide-spectrum genetic conditions on the edge of autism.

We also confirmed previous findings implicating genes involved in chromatin remodeling, transcriptional regulation, and synaptic function in autism ^8^. Genes expressed during prenatal development (*e.g.,* chromatin-related genes) and those expressed later in development (*e.g.,* synaptic genes) were similarly represented across subgroups of autistic people. These results suggest that the presence of certain genetic variants alone is insufficient to fully explain the phenotypic variability in autism, pointing to the influence of additional genetic, environmental, or developmental modifiers ^5^.

We then examined the contribution of common genetic variants by calculating PGS for a range of traits related to cognition, psychiatric outcomes, and well-being. Autistic people without IDD (*i.e.*, cluster C1 and C2) tended to have on average slightly higher autism PGS scores compared to autism with IDD (*i.e.*, cluster C3) ^3^.

We found that cluster C3 of autistic people with low IQ exhibited significantly lower intelligence PGS compared to other clusters. This same cluster was also enriched for *de novo*/rare variants affecting constrained genes and genes previously implicated in NDD. Importantly, both carriers and non-carriers of these rare variants within the C3 cluster showed similarly reduced intelligence PGS (**Supplementary Fig. 24**). These findings support a model in which cognitive function in autism can be shaped by the presence of a single variant, but also by the cumulative impact of rare and common genetic variants ^35^.

Finally, we explored PGS associated with traits such as anxiety, MDD, PTSD, and subjective well-being. PGS for anxiety and MDD were significantly higher in autistic people compared to NT. Of particular interest was the PGS derived from a genome wide association study (GWAS) on the question "*Did you feel loved as a child?*", a proxy for social-affective well-being. Autistic people had on average lower PGS for this trait compared to NT people. While the construct captured by this question is broad and somewhat imprecise, this result raises intriguing questions about the relationship between social-affective genetic profiles and autism. It remains to be determined which specific genetic variants underlie this measure and why they appear less frequently in autistic populations. Notably, this PGS is also significantly lower in the autistic people from the SPARK cohort compared to their non-autistic siblings (**Supplementary Fig. 16**). Further research is needed to dissect the biological underpinnings of this and related social-emotional traits, and how they intersect with neurodevelopmental trajectories in autism.

Our neuro-imaging analyses replicate some of the findings reported in previous brain imaging dataset ^20,36^. We observed an increased cortical thickness in the STG and the right pericalcarine visual *cortex,* especially in autistic people from cluster C2 and C3 respectively. Our findings are aligned with the literature in the field and provide potential insight into the neural substrates of social and cognitive skill and sensory process in autism ^36^. The STG has been repeatedly identified as a key region involved in social perception ^37^, including gaze direction, facial expressions, speech, prosody, and multisensory integration ^37–39^. Hemispheric specialization within the STG is well established: the left STG is primarily involved in semantic and linguistic processing, while the right STG is more engaged in prosody and the pragmatic aspects of language ^40^. In line with this functional asymmetry, our results reveal a significant increase in cortical thickness of the left STG in autistic people compared to NT.

Our findings also point to structural variation in the visual *cortex* as a potentially underappreciated feature of autism. Specifically, we observed increased cortical thickness in the visual pericalcarine region among autistic people from cluster C3 with lower IQ, along with a negative correlation between this *cortex* thickness and IQ across autistic people (**Supplementary Fig. 18**). This association has been largely overlooked in prior research, likely due to IQ thresholds in commonly used cohorts such as ABIDE, which predominantly include people with full-scale IQ above 70. The relevance of visual *cortex* involvement is further supported by recent transcriptomic data from postmortem tissue, showing that the primary visual *cortex* exhibits the largest differential gene expression between autistic and neurotypical people ^41^. This molecular finding aligns with our neuroimaging data and supports the notion that early sensory *cortices* may be more central to autism biology than previously assumed ^42^.

Our finding of reduced rsfMRI connectivity between the visual and somatomotor networks aligns with recent studies ^43,44^ reporting lower connectivity among visual association, somatosensory, and motor networks in autism ^43,45^. Decreased somatomotor connectivity has been linked to altered primary sensory processing ^44^, potentially impairing the ability to filter relevant sensory inputs and generate appropriate motor responses, thereby weakening integration between the somatomotor, salience, and dorsal attention networks ^46^. Within the default mode network, we observed hypoconnectivity of the right temporal region, accompanied by widespread hyperconnectivity both within the DMN and between the DMN and other large-scale systems, notably the frontoparietal and visual networks. These findings are consistent with previous reports using independent datasets, including the ABIDE dataset^43,44,47^.

Consistent with prior work, our group-level analyses revealed no significant differences in resting-state EEG *alpha* peak frequency or spectral power between autistic and neurotypical people, suggesting no global electrophysiological signature of autism ^48^. In contrast, autistic people in cluster C3 showed a significantly lower *alpha* peak frequency compared to NT people. Because *alpha* peak frequency indexes cortical excitability and processing speed, this reduction may reflect slower neural dynamics or altered thalamocortical coupling in C3 which align with evidence linking reduced *alpha* activity and/or *alpha* peak frequency to cognitive impairment and lower non-verbal IQ in autism ^49,50^.

The study offers important strengths, though it is also subject to several limitations. A defining feature of this dataset is its comprehensive collection of multiscale data, ranging from WGS to brain imaging and clinical assessments. The inclusion of NT people, undiagnosed relatives, and numerous families with both biological parents enable robust investigation of *de novo* variants and the interplay between rare and common genetic variants within familial contexts. An additional strength is the inclusion of brain imaging data from autistic people with IDD, a population often underrepresented in previous neuroimaging datasets. Another notable advantage is the harmonization of data across multiple sites and across datasets, including LEAP and InovAND. These harmonization protocols are now being adopted by several European and international research initiatives, highlighting the broader utility and reproducibility of our multidimensional approach ^51^. However, the collection of such a large multiscale dataset also presents certain limitations. One is related to our clustering analysis, which is constrained by the number of people in each cluster. For instance, we were unable to incorporate adaptive functioning measures, such as the VABS, despite their potential to significantly enhance subgroup differentiation, particularly within clusters C1 and C3. VABS data were available for only approximately 500 people, which we deemed insufficient for reliable subgroup characterization, especially given that some clusters included fewer than 100 people. A second limitation concerns the interpretation of increased cortical thickness observed in the temporal and visual *cortices*. These findings should be approached with caution due to the age range of our cohort, which spans primarily from 6 to 18 years. From a developmental standpoint, the observed increases in cortical thickness may reflect delayed cortical thinning, a process that typically begins before age 6 ^52^. To fully understand these developmental trajectories, future studies should aim to include younger people or a follow-up cohort. This would allow for characterization of earlier stages of brain maturation and provide a more comprehensive understanding of neurodevelopment in autism. Rather than interpreting results in terms of absolute increases or decreases in cortical thickness, future research should adopt a growth chart-based framework, leveraging age-specific developmental norms to better assess deviations from typical cortical development ^53–55^. Several ongoing initiatives are now focused on collecting longitudinal data beginning in early childhood ^53^. As part of this effort, our consortium will contribute to a new longitudinal cohort PIP (Preschooler Brain Imaging and Behavior Project; https://www.aims-2-trials.eu/pip/) focusing on children aged 3-6 years. This dataset will enable deeper exploration of early developmental trajectories and help clarify the neural mechanisms underlying autism from a much earlier age.

Our results also open new perspectives for understanding the biological and developmental foundations of autism in its multidimensionality. Understanding the complex interplay between genetic variation, brain structure/function, and developmental trajectories in autism remains a major challenge. Comprehensive insights will ultimately require large, multiscale datasets that integrate genomic, neuroimaging, and clinical data across diverse populations. While the current dataset represents a significant step in this direction, enabling analyses across these interconnected levels, further expansion will be essential to capture the full spectrum of variability. Nonetheless, our findings offer preliminary insights into how genetic variations, spanning *de novo*/rare, and common variants, relate to structural/functional brain differences and clinical profiles in autism. By bridging molecular and neural phenotypes with cognitive and behavioral outcomes, this work helps to decipher the heterogeneity of autism and may in future inform the personalized support that each autistic person and their family wants and needs.

## Online Methods

### Dataset and participants

For the LEAP study, participants were recruited between January 2014 and March 2017 across six European centers: Institute of Psychiatry, Psychology and Neuroscience, King’s College London (IoPPN/KCL, UK), Autism Research Centre, University of Cambridge (UCAM, UK), University Medical Centre Utrecht (UMCU, Netherlands), Radboud University Nijmegen Medical Centre (RUNMC, Netherlands), Central Institute of Mental Health (CIMH, Germany) and the University Campus Bio-Medico (UCBM) in Rome, Italy. At each site, an independent ethics committee approved the study. All participants (where appropriate) and their parent/legal guardians provided written informed consents for research and data sharing ^17,18^. C07- InovAND participants were recruited between 2007 and 2016 in four French hospitals: Hopital Robert-Debré (Paris), Hopital Albert Chenevier (Creteil), Hopital Alpes Isère (Grenoble) and Hopital Charles Perrens (Bordeaux). For both studies, all participants and their parent/legal guardians provided written informed consent upon inclusion.

### Genetics

#### DNA isolation and sequencing

DNA was extracted from whole blood/saliva, using manufacturer’s protocols on a Tecan Freedom EVO®-HSM workstation and a Maxwell® RSC 48 Instrument, and sequenced by the CNRGH using short read Illumina technology at a read depth of 30× on average. A total of 2,560 samples were sequenced, and reads were aligned to GRCh37.

#### Variant detection

Individual gVCF were called using DeepVariant and genotyped jointly using Glnexus. Detected SNVs/indel of interest were validated by visualization of the CRAM files and then by Sanger sequencing. For SV, we first ran WisecondorX and ClinSV. A total of 2,189 variants were manually visualized and further used to create a random forest model to discriminate true SV from noise. For short tandem repeats (STR), we ran ExpansionHunter using the loci dictionary from stripy.org and validated using targeted PCR. The list of genes carrying variants analyzed in this study is available in **Supplementary Data 1**.

#### Ancestry

We computed the admixture of the cohort along with the Human Genome Diversity Project. We then predicted the superpopulation (EUR, AFR, EAS, CSA, MID, OCE, AME) with a random forest model using admixture results and known data from the Human Genome Diversity Project.

#### Polygenic scores

We used summary statistics from different GWAS and used SBayesRC to compute the PGS. All individual PGS were corrected for ancestry using the first 10 principal components.

#### Statistical analysis

Differences between categorical variables were assessed using the Chi-square test, with effect sizes quantified by Cramer’s V. When necessary, Fisher’s exact approach was applied to adjust for small sample sizes or sparse data. Comparisons between independent quantitative variables were performed using the Mann-Whitney-Wilcoxon test; effect sizes were estimated using the rank biserial correlation when appropriate. Statistical significance was defined as *p* < 0.05.

For genetic data, associations between the presence of genetic variants and categorical variables were assessed using Fisher’s exact test, with false discovery rate (FDR, one-sided independent t-tests adjusted for multiple comparisons using Benjamini–Hochberg correction) correction. Odds ratios (ORs) with 95% confidence intervals (CI95%) were calculated using the NT group or, when appropriate, the combined NT/sibling group as the reference category.

#### Gene lists

The list of genes used in this study is available on our website GeneTrek (https://genetrek.pasteur.fr/) and in **Supplementary Data 1**. The notion of gene constraint is based on the union of GnomAD v4 for autosomes, GnomAD v2 for X chromosome and sHET ^35^.

#### Replication in the SPARK cohort

We replicated the genetic analyses in the SPARK cohort using whole exome sequence (WES) data from the iWES v.3 dataset.

### Brain Imaging analyses

#### MRI Acquisition

For the LEAP cohort, Magnetic resonance imaging (MRI) scans were acquired across five European research sites (University of Cambridge, King’s College London, Central Institute of Mental Health Mannheim, Radboud University Nijmegen Medical Centre, and University Medical Center Utrecht) using a harmonized protocol based on ADNI- 2/GO recommendations. High-resolution T1-weighted structural images were acquired using a magnetization-prepared rapid gradient-echo (MPRAGE) sequence (1.2 mm isotropic resolution). Resting-state functional MRI (rsfMRI) scans were acquired using single-echo echo-planar imaging (EPI) with full brain coverage (see **Supplementary Tables 4 and 5** for acquisition parameters). In the InovAND cohort, MRI data were collected at a single site (Robert Debré Hospital, Paris) between 2010 and 2024, using three Philips scanners (1.5T and 3T field strengths). T1-weighted images were acquired with Fast Field Echo (FFE) or Turbo Field Echo (TFE) sequences (voxel size 1 mm isotropic, TR = 7–25 ms, TE = 3–5.6 ms, FOV = 240–260 mm). Updated 3D-TFE protocols with compressed sensing and SENSE acceleration achieved voxel resolutions up to 0.9 mm isotropic. Resting-state fMRI was acquired with single-echo EPI sequences (TR = 2000–2500 ms; TE = 30–35 ms; voxel size = 3 × 3 × 3 mm). Detailed sequence parameters are listed in **Supplementary Table 6 and 7**.

#### Structural MRI data preprocessing

T1-weighted structural images were processed using FreeSurfer (v6.0.0) Cliquez ou appuyez ici pour entrer du texte. with the full *recon-all* pipeline. Quality control of cortical and subcortical segmentations was performed manually by trained raters blinded to diagnosis and scans failing manual QC were excluded. Cortical thickness (CT) and surface area (SA) were extracted from 68 cortical regions (Desikan–Killiany atlas ^25^) and volumes from 14 subcortical regions and lateral ventricles. Cortical thickness and surface area were extracted for 68 cortical regions according to the Desikan–Killiany atlas, while volumes were obtained for 14 subcortical regions and the lateral ventricles. Global metrics, including mean cortical thickness, total surface area, intracranial volume, total gray matter, and cerebrospinal fluid volume, were also derived. For each regional measure, residuals were computed after modeling diagnosis, age, sex, age squared, MRI site, and intracranial volume. Site-related variability was mitigated using ComBat ^57^ harmonization. Residuals were z-scored relative to neurotypical controls, and group differences were assessed using two-sided Welch’s t-tests with Bonferroni correction (P_BON_ < 0.05).

#### fMRI preprocessing and analyses

Preprocessing of rsfMRI data was performed using fMRIPrep (v23.0.0; ^58^) and XCP-D (v0.8.0; ^59^). Steps included realignment, slice-timing correction, coregistration to the participant’s T1-weighted image using FLIRT, normalization to MNI152NLin2009cAsym space, and resampling to 2 mm isotropic resolution. Spatial smoothing was applied using a 6 mm FWHM Gaussian kernel. Physiological and motion confound regression was conducted using the 36p+Despike model. This included removal of six motion parameters and their derivatives, global signal, white matter and CSF signals, their squares, and quadratic terms, followed by bandpass filtering (0.01–0.08 Hz). Volumes with excessive motion were scrubbed when mean framewise displacement (FD) exceeded 0.5 mm or when more than 20% of volumes exceeded the FD threshold. Participants not meeting these motion criteria were excluded. Mean FD values per group are reported in **Supplementary Fig. 19a**. Functional connectivity (FC) was estimated at the ROI level using the Schaefer 100- parcel cortical atlas ^60^ combined with 15 subcortical ROIs from CIT168 subcortical atlas ^61^. For each participant, mean time series were extracted per ROI, and pairwise Pearson correlations were computed to generate a 115 × 115 symmetric connectivity matrix. Correlation coefficients were Fisher z-transformed prior to group-level analyses. FC matrices were standardized by subtracting the median and dividing by the interquartile range (IQR) to ensure comparable scaling across participants. For each connectivity feature, residuals were computed after modeling diagnosis, age, sex, age squared, MRI site. To mitigate site-related variability, ComBat harmonization was applied to all FC features ^57^. The efficiency of harmonization was confirmed using one-vs-one SVM classification of acquisition site before and after ComBat (**Supplementary Fig. 19b**). Between-group and between-cluster differences were evaluated for each connection using two-sided Welch’s t-tests, with FDR correction (q < 0.05). Connections were classified as hyper- or hypoconnected based on the direction of the t-statistic, and network-level summaries were derived by counting altered edges within and between Yeo 7-network systems ^43^.

#### Resting state EEG acquisition

Five sites acquired EEG data at baseline. In the LEAP cohort, 4 minutes of resting state EEG were recorded per participant (2 minutes with eyes open and 2 minutes with eyes closed) with 30s blocks alternatively, using 10–20 layout caps with 60–70 electrodes. In the InovAND cohort, all subjects were recorded in resting-state with eyes open and closed for periods of 3 to 5 minutes using a 129-GSN hydrocel Geodesic montage. Sampling was at 1,000 Hz, and the reference electrode was the Cz position.

#### Resting-state EEG data preprocessing

Preprocessing was done using the MNE-Python software ^62^, according to the OHBM COBIDAS MEEG good practice recommendations ^63^. EEG signals were band-pass filtered at 1–100 Hz (FIR) and notched at 50 Hz to suppress line noise. Robust referencing and bad-channel handling were performed per subject with PyPREP, and bad channels were interpolated with spherical splines before setting a common average reference. We fit ICA using FastICA to the epoched data and classified components with ICLabel ^64^; components labeled as non-brain sources (e.g., eye, muscle, heart, line noise, channel noise) were excluded and the cleaned solution was applied to the data. Next, EEG signals were segmented into 4-second epochs. The Autoreject toolbox ^65^ was used to interpolate or reject bad epochs. Then, we extracted the spectral features across all bands (*theta*, *delta*, *alpha*, *beta,* and *gamma*). Alpha peak frequency was identified as the maximum amplitude within the 6–15 Hz range. Further methodological details are provided in **Supplementary Section 5**.

Statistical Correction: For each feature (power band × brain region × laterality), linear regression was performed to remove effects of age, age-squared, and sex. ComBat harmonization was then applied to the regression residuals to remove site and cohort effects, with age, sex, and age-squared included as covariates. ComBat-adjusted residuals were z-scored based on the mean and standard deviation of the TD control group. Subjects with any feature value exceeding 10 standard deviations (z-score > 10) were excluded as outliers. Between-group differences in corrected PSD measures and alpha peak frequencies were assessed using two-sided Welch’s t-tests with FDR correction (q < 0.05) applied independently for each cohort and for the combined dataset.

#### Clustering analyses

We began with a broad set of clinical measures, including IQ subscales (full-scale, verbal, non-verbal), SRS-2 total t-score, Vineland composite and domain scores, RBS-R, and SSP. PCA was conducted after excluding people with missing data and relatives (n = 380) and standardizing all features. The first two components explained most of the variance (>75%), with IQ and SRS contributing most strongly whereas Vineland scores contributed substantially to PC1 but minimally to PC2 (**Supplementary Fig. 21**). Restricting analyses to IQ and SRS also maximized sample size (n = 1,026 vs. ∼500 with VABS; **Supplementary Table 12**). For both statistical and practical reasons, clustering focused on these two dimensions, Full methodological details are provided in **Supplementary Section 6**.

To determine the optimal number of clusters, we evaluated k = 2–10 using a multi-criterion approach. NbClust, which computes 26 internal validation indices, identified three clusters as the best partition (10/26 indices), with a secondary peak at k = 8. Additional model-based and partition-based indices (AIC, BIC, ICL, Silhouette coefficient, Variance Ratio Score, Davies–Bouldin index, Pseudo-F statistic) and the Gap statistic also favored k = 3 (**Supplementary Table 13**).

## Data Availability

The datasets generated and analyzed in this study, including all modalities from the LEAP (EUAIMS / AIMS2 TRIALS) and InovAND cohorts are securely stored on ELIXIR LU servers, part of the European infrastructure for life science information, based at the Luxembourg Centre for Systems Biomedicine (LCSB) and supported by the Luxembourg National Data Service (LNDS). The LEAP dataset (clinical, cognitive, eye-tracking, neuroimaging, and genetic data) is available via the ELIXIR Luxembourg data catalog. The InovAND dataset will be made available through the same repository upon publication. Access to both datasets is granted upon reasonable request following review and approval by the Data Access Committee, including scientific leads, ethics experts, and Autism community representatives. Requests should be submitted via the data catalog website with a detailed project proposal describing the intended use of the data and must be aligned with General Data Protection Regulation (GDPR) requirements as well as the AIMS2 TRIALS consortium or InovAND data sharing policies, respectively.

## Acknowledgments

The authors thank all the participants of the LEAP and InovAND cohorts for their involvement in the research on autism. This work was funded by Institut Pasteur, Université Paris Cité, the Simons Foundation Autism Research Initiative (SFARI award #240059), the Bettencourt-Schueller Foundation, the GenMed Labex, and AIMS-2- TRIALS, which received support from the Innovative Medicines Initiative 2 Joint Undertaking under grant agreement No 777394 for the project AIMS-2-TRIALS. This Joint Undertaking receives support from the European Union’s Horizon 2020 research and innovation program and EFPIA and AUTISM SPEAKS, Autistica, SFARI, and the Inception program (Investissement d’Avenir grant ANR-16-CONV-0005). This project has received funding from the European Union’s Horizon 2020 Research and Innovation Program under grant 847818 (CANDY), and from Horizon Europe under grant 101057385 (R2D2-MH). Views and opinions expressed are, however, those of the authors only and do not necessarily reflect those of the European Union. Neither the European Union nor the granting authority can be held responsible for them. This work benefited from the DNA & cell bank core facility, at the ICM-Paris Brain Institute. This work received support from the French government, managed by the National Research Agency (Agence Nationale de la Recherche), under the France 2030 program, reference ANR-23-IAHU-0010. Part of this work was funded by a grant from the Conseil Régional d’Ile de France (grant number EX024087).

## Data Availability

The datasets generated and analyzed in this study—including all modalities from the LEAP (EU-AIMS / AIMS-2-TRIALS) and InovAND cohorts are securely stored on ELIXIR-LU servers, part of the European infrastructure for life science information, based at the Luxembourg Centre for Systems Biomedicine (LCSB) and supported by the Luxembourg National Data Service (LNDS) (https://elixir-luxembourg.org/). The LEAP dataset (clinical, cognitive, eye-tracking, neuroimaging, and genetic data) is available via the ELIXIR-Luxembourg data catalog at https://datacatalogue.elixir-luxembourg.org/e/study/ELU-4-4D20BB-1. The InovAND dataset will be made available through the same repository upon publication. Access to both datasets is granted upon reasonable request following review and approval by the Data Access Committee, including scientific leads, ethics experts, and Autism community representatives. Requests should be submitted via the data catalog website with a detailed project proposal describing the intended use of the data and must be aligned with General Data Protection Regulation (GDPR) requirements as well as the AIMS-2- TRIALS consortium or InovAND data sharing policies, respectively.

## Code availability

A GitHub repository will be made available upon publication. Analysis scripts can be obtained from the corresponding authors upon reasonable request.

---

**Table 1:**
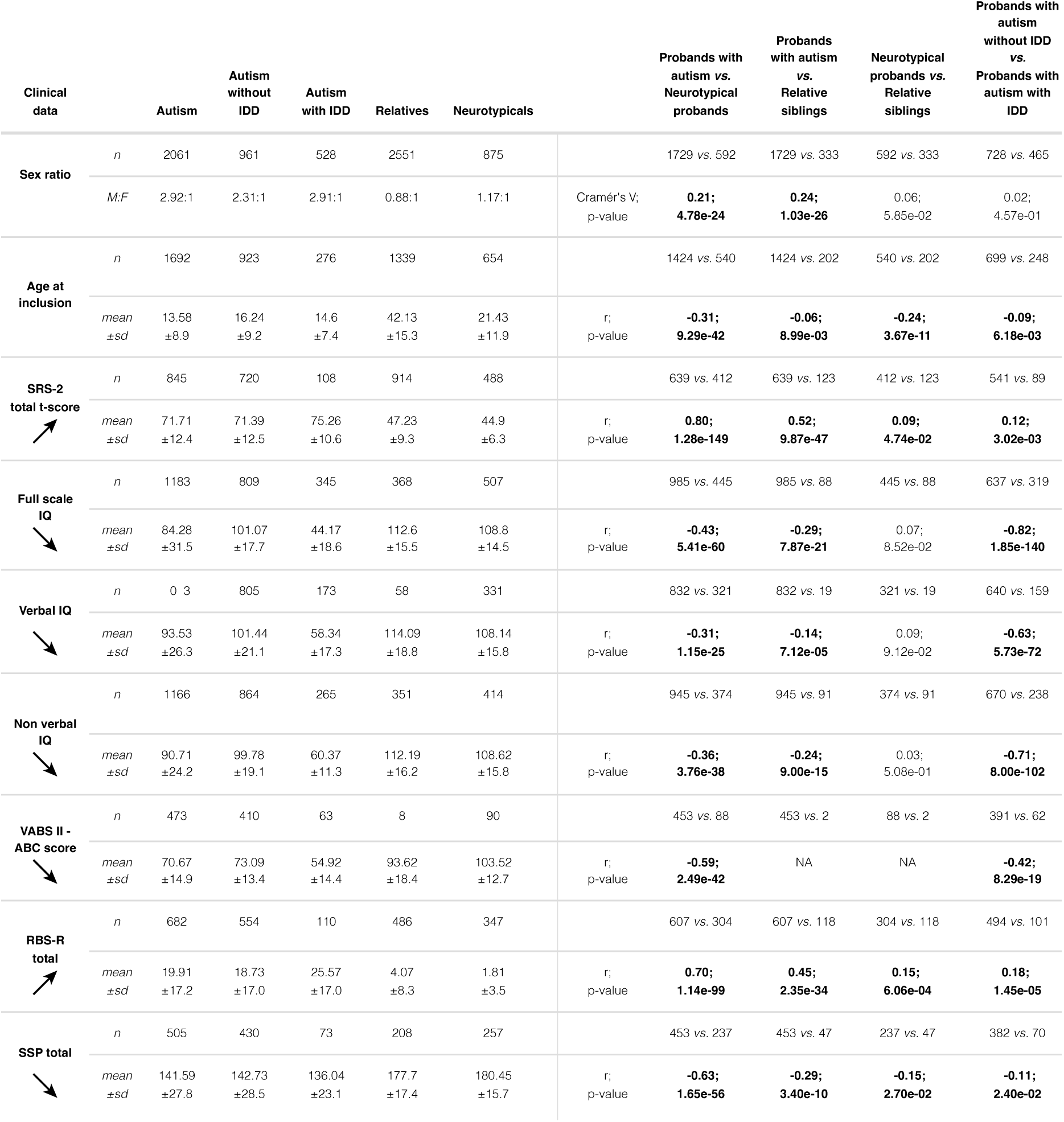
Clinical characteristics of the people enrolled in LEAP-InovAND. Continuous variables were compared using the Mann-Whitney-Wilcoxon test due to non-normal distribution associated with rank-biserial correlation for the effect size, while categorical variables were analyzed using the Chi-square (χ²) test associated with Cramér’s V for the effect size. Statistically significant differences between groups were highlighted where applicable. The interpretation of effect sizes is as follows; absolute value near or below 0.1: small effect, absolute value near 0.3: middle effect, absolute value near or greater than 0.5: large effect. A positive effect size indicates that the first group has a higher score than the second group, whereas a negative effect size indicates that the second group has a higher score than the first. The arrows next to the test names indicate the direction in which symptom severity is highest. IDD: intellectual developmental disorder, IQ: intelligence quotient, NA: not applicable, RBS-R: repetitive behavior scale-revised, SRS-2: social responsiveness scale 2nd edition, SSP: short sensory profile, NT: neurotypical people, VABS II - ABC: vineland adaptative behaviors scale 2nd edition - communication, daily living skills and socialization

