## Supplementary Information for "LEAP-InovAND a multiscale resource to explore genetics, brain imaging and clinical data in autism"

### Section 1. Participatory research

#### Collaboration with the autism's representatives (A-Reps)

The A-Reps are a group of autistic people, parents and caregivers from across Europe who have agreed to bring their perspectives on being autistic or caring for an autistic loved one to the AIMS-2-TRIALS research program. They are coordinated by a team at the University of Cambridge and were recruited in early 2019 from nearly 100 European applications. Recruitment attempted to account for a range of strengths, needs and challenges of autistic people, factors such as gender, age, intellectual ability, nationality, life experience and interests were also collected. Final group selection was made with the support of the A-Reps Steering Committee, a small group of interested people invited by Autistica to help oversee the running of A-Reps group, throughout the project.

A-Reps have shaped the design of research protocols and questionnaires for various studies. A-Reps were invited to attend specially created webinars about biomarkers, biomarker development and genetics, which included opportunities for questions and discussion. This webinar series helped to ensure that the A-Reps have the background knowledge required to engage in discussions about AIMS-2-TRIALS research and sustainability, for example in the biomarker and data sharing working groups. This started with an introductory webinar early in the project to explain more about genetics research and the plans for this project as well as introducing the internal database for the project. Later in this webinar series, one of the sessions focused on this cohort and on the data, we wanted to highlight. Thanks to this webinar, we received feedback that helped make some of the data we present more accessible, but above all, it allowed us to consider the questions and concerns raised by the A-Reps who attended.

For this article, we provided a draft of the manuscript and left one month to suggest and provide feedback. We also organized a specific virtual meeting to address directly the questions. We would like to express our particular thanks to everyone from the A-Reps who took part in this work and significantly contributed to improving it.

#### Section 2. Demographic and clinical data

We combined data from 2 large datasets: the Longitudinal European Autism Project LEAP (https://www.eu-aims.eu/ and https://www.aims-2-trials.eu/) and InovAND (https://robertdebre.aphp.fr/creation-centre-excellence-autisme/), see **Supplementary Fig. 1**. Our final sample for analysis included 2,061 people with autism (524 females; age range: 1.7–70 years, mean age ± sd: 13.58 ± 8.90), 854 neurotypical (NT) people (389 females; age range: 2-65 years, mean age ± sd: 21.43 ± 11.94) and 2,551 relatives (1,353 females; age range: 1–83 years, mean age ± sd: 42.13 ± 15.29). The clinical and demographic characteristics of included participants are listed in **Supplementary Tables 1, 2 and 3, and Supplementary Fig. 3.**

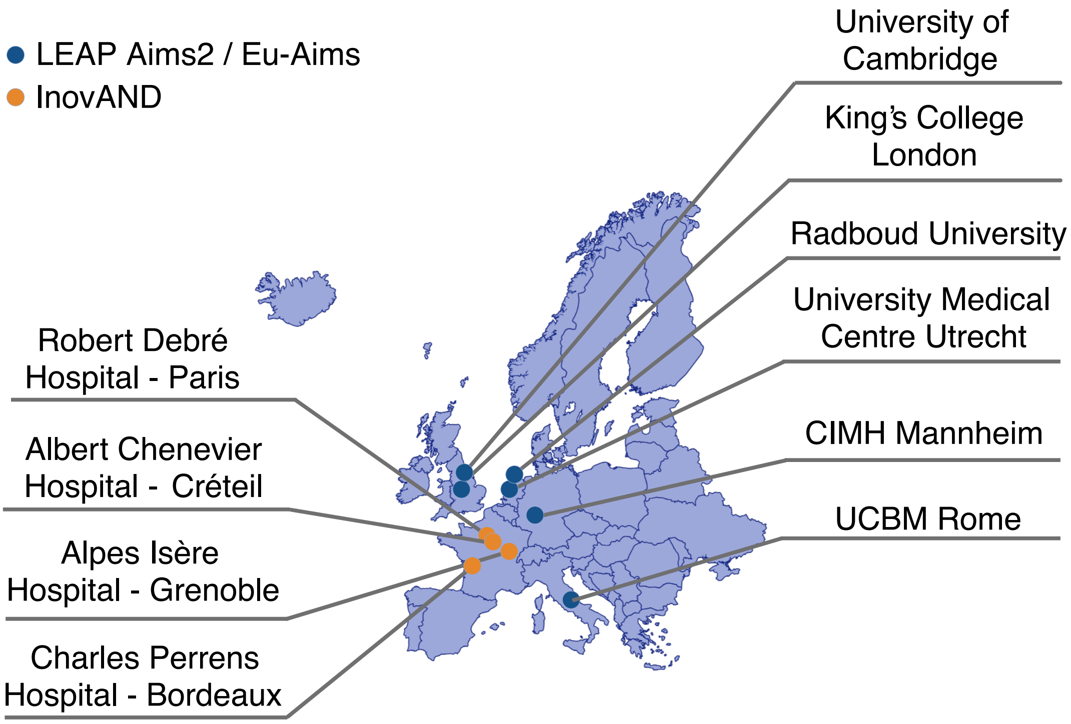

Supplementary Figure 1: Centers which have included the participants from the LEAP and InovAND datasets.

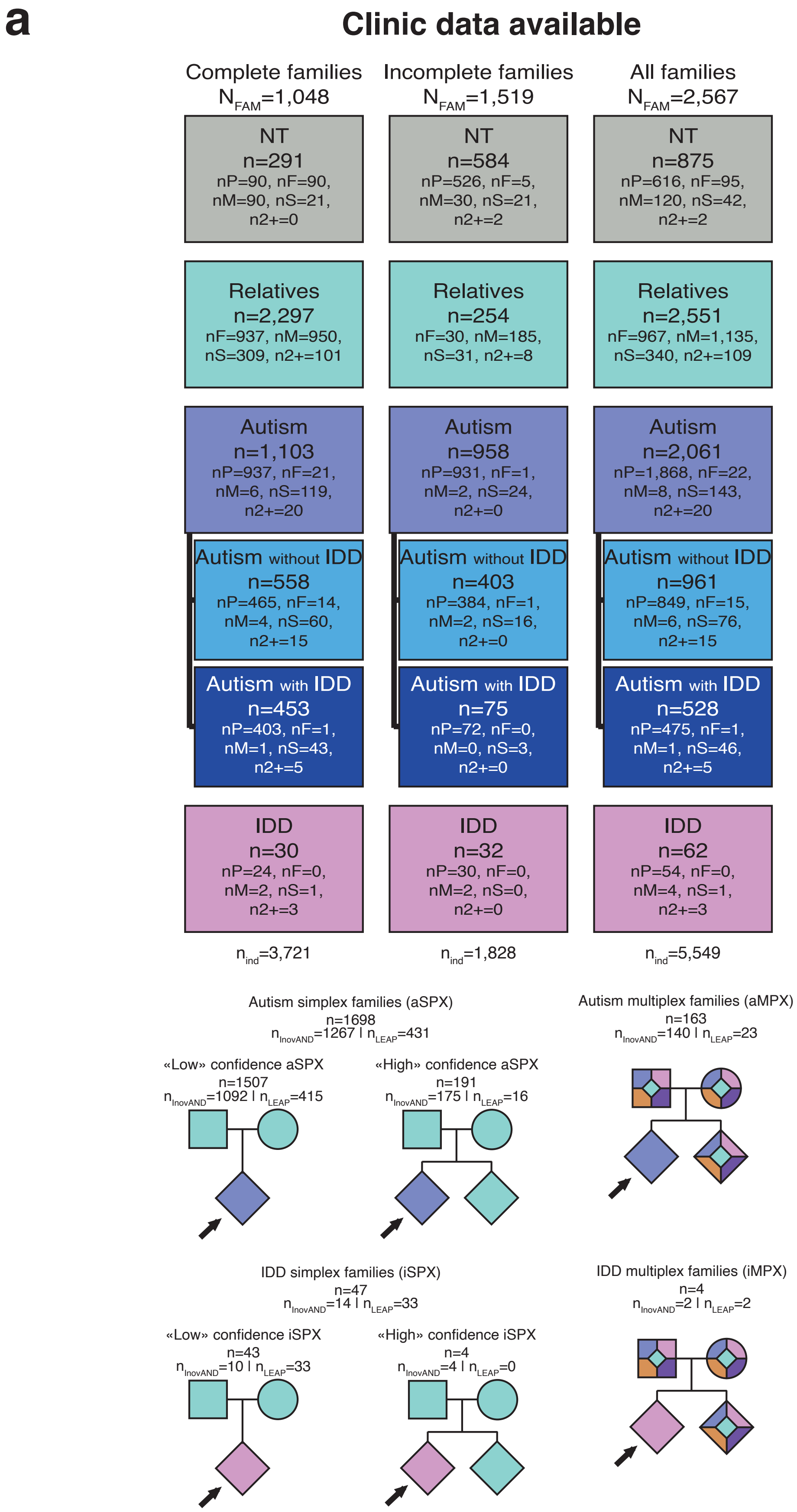
a
Clinic data available
Complete families
$N_{FAM}$=1,048
Incomplete families
$N_{FAM}$=1,519
All families
$N_{FAM}$=2,567
NT
n=291
nP=90, nF=90, nM=90, nS=21, n2+=0
NT
n=584
nP=526, nF=5, nM=30, nS=21, n2+=2
NT
n=875
nP=616, nF=95, nM=120, nS=42, n2+=2
Relatives
n=2,297
nF=937, nM=950, nS=309, n2+=101
Relatives
n=254
nF=30, nM=185, nS=31, n2+=8
Relatives
n=2,551
nF=967, nM=1,135, nS=340, n2+=109
Autism
n=1,103
nP=937, nF=21, nM=6, nS=119, n2+=20
Autism
n=958
nP=931, nF=1, nM=2, nS=24, n2+=0
Autism
n=2,061
nP=1,868, nF=22, nM=8, nS=143, n2+=20
Autism without IDD
n=558
nP=465, nF=14, nM=4, nS=60, n2+=15
Autism without IDD
n=403
nP=384, nF=1, nM=2, nS=16, n2+=0
Autism without IDD
n=961
nP=849, nF=15, nM=6, nS=76, n2+=15
Autism with IDD
n=453
nP=403, nF=1, nM=1, nS=43, n2+=5
Autism with IDD
n=75
nP=72, nF=0, nM=0, nS=3, n2+=0
Autism with IDD
n=528
nP=475, nF=1, nM=1, nS=46, n2+=5
IDD
n=30
nP=24, nF=0, nM=2, nS=1, n2+=3
IDD
n=32
nP=30, nF=0, nM=2, nS=0, n2+=0
IDD
n=62
nP=54, nF=0, nM=4, nS=1, n2+=3
$n_{ind}$=3,721
$n_{ind}$=1,828
$n_{ind}$=5,549
Autism simplex families (aSPX)
n=1698
$n_{InovAND}$=1267 | $n_{LEAP}$=431
«Low» confidence aSPX
n=1507
$n_{InovAND}$=1092 | $n_{LEAP}$=415
«High» confidence aSPX
n=191
$n_{InovAND}$=175 | $n_{LEAP}$=16
Autism multiplex families (aMPX)
n=163
$n_{InovAND}$=140 | $n_{LEAP}$=23
IDD simplex families (iSPX)
n=47
$n_{InovAND}$=14 | $n_{LEAP}$=33
«Low» confidence iSPX
n=43
$n_{InovAND}$=10 | $n_{LEAP}$=33
«High» confidence iSPX
n=4
$n_{InovAND}$=4 | $n_{LEAP}$=0
IDD multiplex families (iMPX)
n=4
$n_{InovAND}$=2 | $n_{LEAP}$=2

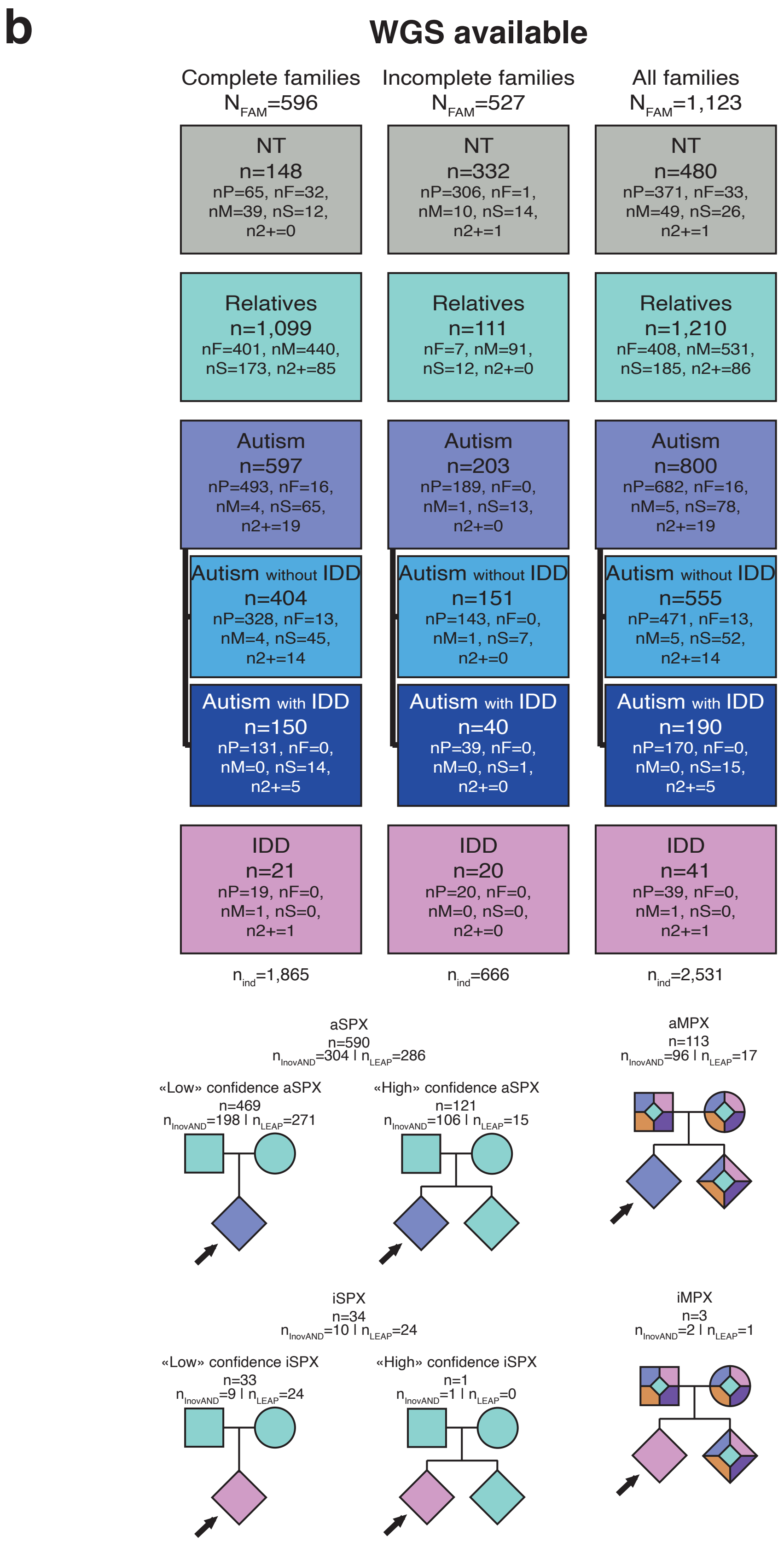
b
WGS available
Complete families
$N_{FAM}$=596
Incomplete families
$N_{FAM}$=527
All families
$N_{FAM}$=1,123
NT
n=148
nP=65, nF=32, nM=39, nS=12, n2+=0
NT
n=332
nP=306, nF=1, nM=10, nS=14, n2+=1
NT
n=480
nP=371, nF=33, nM=49, nS=26, n2+=1
Relatives
n=1,099
nF=401, nM=440, nS=173, n2+=85
Relatives
n=111
nF=7, nM=91, nS=12, n2+=0
Relatives
n=1,210
nF=408, nM=531, nS=185, n2+=86
Autism
n=597
nP=493, nF=16, nM=4, nS=65, n2+=19
Autism
n=203
nP=189, nF=0, nM=1, nS=13, n2+=0
Autism
n=800
nP=682, nF=16, nM=5, nS=78, n2+=19
Autism without IDD
n=404
nP=328, nF=13, nM=4, nS=45, n2+=14
Autism without IDD
n=151
nP=143, nF=0, nM=1, nS=7, n2+=0
Autism without IDD
n=555
nP=471, nF=13, nM=5, nS=52, n2+=14
Autism with IDD
n=150
nP=131, nF=0, nM=0, nS=14, n2+=5
Autism with IDD
n=40
nP=39, nF=0, nM=0, nS=1, n2+=0
Autism with IDD
n=190
nP=170, nF=0, nM=0, nS=15, n2+=5
IDD
n=21
nP=19, nF=0, nM=1, nS=0, n2+=1
IDD
n=20
nP=20, nF=0, nM=0, nS=0, n2+=0
IDD
n=41
nP=39, nF=0, nM=1, nS=0, n2+=1
$n_{ind}$=1,865
$n_{ind}$=666
$n_{ind}$=2,531
aSPX
n=590
$n_{InovAND}$=304 | $n_{LEAP}$=286
«Low» confidence aSPX
n=469
$n_{InovAND}$=198 | $n_{LEAP}$=271
«High» confidence aSPX
n=121
$n_{InovAND}$=106 | $n_{LEAP}$=15
aMPX
n=113
$n_{InovAND}$=96 | $n_{LEAP}$=17
iSPX
n=34
$n_{InovAND}$=10 | $n_{LEAP}$=24
«Low» confidence iSPX
n=33
$n_{InovAND}$=9 | $n_{LEAP}$=24
«High» confidence iSPX
n=1
$n_{InovAND}$=1 | $n_{LEAP}$=0
iMPX
n=3
$n_{InovAND}$=2 | $n_{LEAP}$=1

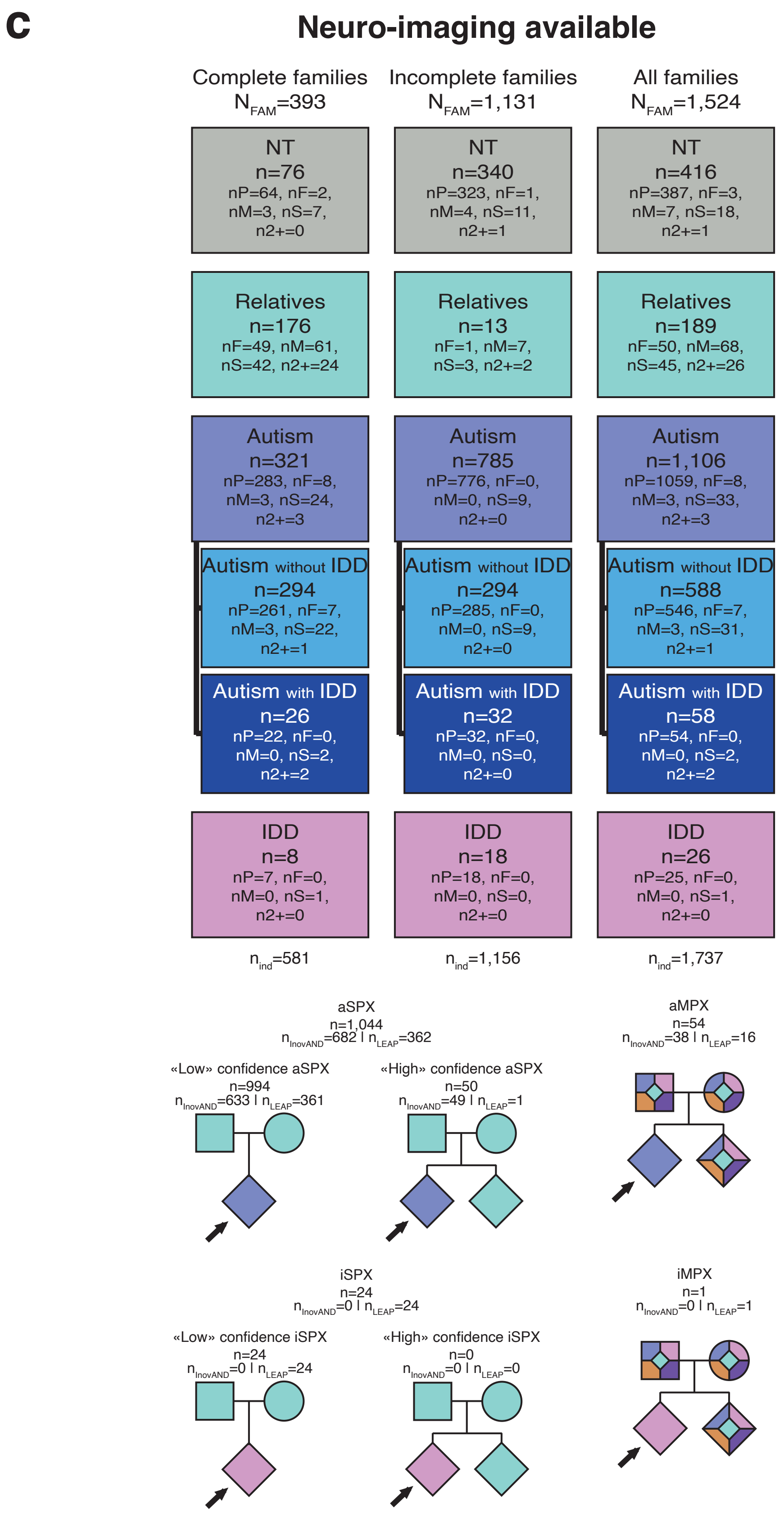

Supplementary Figure 2: Count of all families within LEAP-InovAND and description of the different clinical categories (a), with whole genome sequencing (b) and with either EEG and/or MRI (c). A family is considered multiplex when there is at least one person with autism, IDD, ADHD and/or epilepsy in addition to the autistic (aMPX) or IDD (iMPX) proband. Conversely, simplex families are those in which, with the current clinical knowledge of the families, no one other than the proband is diagnosed with NDD. The notion of "low" confidence means that there are no other siblings in the family, so the SPX criterion is uncertain, whereas the notion of "high" confidence reflects the presence of at least one other sibling without NDD. In the genetic trees, orange and purple represent ADHD and epilepsy, respectively. Complete families correspond to families with both parents of the proband, while incomplete families correspond to families where at least one parent is missing. P: probands, F, fathers, M: mothers, S: siblings, 2+: extended family members.

*Clinical Diagnosis:* Inclusion criteria for the EU-AIMS/AIMS-2-Trials LEAP dataset were an existing clinical diagnosis of autism according to DSM-IV, DSM-5, or ICD-10, with the majority also meeting the cut-offs criteria on the Autism Diagnostic Interview-Revised (ADI-R) [1] and the Autism Diagnostic Observation Schedule 2 (ADOS 2 [2]) (see Charman *et al.* [3,4]). Similarly, inclusion criteria for InovAND were an existing clinical diagnosis of autism based on DSM-IV-TR criteria, associated with ADI-R and ADOS evaluations.

*Autistic traits:* Overall, the intensity of autism-related traits was assessed using a variety of instruments. The Autism Diagnostic Observation Schedule-2 (ADOS-2) [2] was administered to probands and their siblings, utilizing different modules tailored to each participant's age and verbal language ability. For comparison across the different modules, the total calibrated severity scores (CSS) were used for analysis. Additionally, caregivers completed the social responsiveness scale-2 (SRS-2) [5], which measures the overall intensity of autistic symptoms. For the SRS-2, t-scores were used. Parents also completed self-reported questionnaires [*e.g.* SRS-2, Hansen Research Services Matrix Adaptive Test (HRS-MAT)]. Across all instruments, higher scores were indicative of greater intensity in autism-related traits symptoms.

*Social communication skills*: To evaluate social communication skills, social affect CSS scores from the ADOS-2 (ADOS SA) were utilized, alongside the autism diagnostic interview, revised (ADI-R) [1] social interaction (ADI-R A) and communication domains (ADI-R B). The ADI-R, consisting of a semi-structured interview with caregivers, was administered by trained professionals who rated each question item. Based on the diagnostic algorithm, four domain scores were aggregated. Considering each participant's developmental trajectory, the communication domain within the ADI-R was further divided into subdomains, specifically catering to people with and without fluent verbal communication (ADI-R B verbal and non-verbal). Consistent with the scoring approach of the ADOS-2 CSS, higher scores on the ADI-R indicated greater difficulties in social communication.

*Restricted interest and repetitive behavior:* Restricted and repetitive behaviors (RRB) were assessed using the RRB Calibrated Severity Score (RRB CSS) from the ADOS-2 and the RRB subdomain (domain C) from the ADI-R. In addition, the Repetitive Behavior Scale-Revised (RBS-R), a parent-report questionnaire, was used to capture parent-reported repetitive behaviors.

*Cognitive ability*: Intelligence was assessed using the Wechsler Preschool and Primary Scale of Intelligence (WPPSI), Wechsler Intelligence Scale for Children (WISC) [6], and Wechsler Adult Intelligence Scale. For people with limited verbal language abilities, the Leiter international Performance Scale-Revised [7] was performed to measure non-verbal intelligence quotient (IQ). The Raven's Progressive Matrices test was used to assess the IQ of the probands' relatives.

*Adaptive behaviors*: The Vineland adaptive behavior scale (VABS)-II [8] has been utilized to assess adaptive functioning in children and their siblings. The VABS-II is a caregiver-report questionnaire covering various domains, with each item inquiring whether the child can perform the specified task. Four domain scores, socialization, communication, daily living, and motor skills, along with a composite score, were calculated and standardized based on age-matched NT people. Lower scores in each of these domains were indicative of developmental delays.

*Statistical Analysis:* Clinical data were summarized using means and standard deviations. To assess statistically significant differences between participant groups, the Chi-square ($\chi^2$) test was applied to categorical variables. Cramér's V [9] was used to estimate effect sizes for these comparisons. For continuous variables, which were found not to follow a normal distribution, the Mann–Whitney–Wilcoxon test was employed. Rank-biserial correlation was used to quantify the corresponding effect sizes. All statistical analyses were conducted using two-tailed tests, and a p-value below 0.05 was considered statistically significant. No correction for multiple testing (*e.g.*, FDR adjustment) was applied.

Supplementary Table 1: Clinical table of the LEAP cohort. Continuous variables were compared using the Mann–Whitney–Wilcoxon test due to non-normal distribution associated with rank-biserial correlation for the effect size, while categorical variables were analyzed using the Chi-square ($\chi^2$) test associated with Cramér's V for the effect size. Statistically significant differences between groups were highlighted where applicable. The interpretation of effect sizes is as follows; absolute value near or below 0.1: small effect, absolute value near 0.3: middle effect, absolute value near or greater than 0.5: large effect. A positive effect size indicates that the first group has a higher score than the second group, whereas a negative effect size indicates that the second group has a higher score than the first. The arrows next to the test names indicate the direction in which symptom severity is highest. Note: IDD: intellectual developmental disorder, IQ: intelligence quotient, NA: not applicable, RBS-R: repetitive behavior scale-revised, SRS: social responsiveness scale 2nd edition, SSP: short sensory profile, NT: neurotypical people, VABS II - ABC: vineland adaptative behaviors scale 2nd edition - communication, daily living skills and socialization.

| Clinical data LEAP | | Autism | Autism without IDD | Autism with IDD | Relatives | Neurotypicals | | Probands with autism *vs.* Neurotypical probands | Probands with autism *vs.* Relative siblings | Neurotypical probands *vs.* Relative siblings | Probands with autism without IDD *vs.* Probands with autism with IDD |
|---|---|---|---|---|---|---|---|---|---|---|---|
| **Sex ratio** | *n* | 492 | 411 | 51 | 697 | 466 | | 458 *vs.* 291 | 458 *vs.* 12 | 291 *vs.* 12 | 388 *vs.* 49 |
| | *M:F* | 2.57:1 | 2.81:1 | 1.83:1 | 0.69:1 | 1.44:1 | Cramér's V; p-value | **0.08; 3.85e-02** | 0.00; 9.33e-01> | 0.00; 1.00e+00 | 0.04; 4.63e-01 |
| **Age at inclusion** | *n* | 453 | 403 | 50 | 524 | 275 | | 433 *vs.* 268 | 433 *vs.* 1 | 268 *vs.* 1 | 384 *vs.* 49 |
| | *mean ± sd* | 16.77 ±5.9 | 16.5 ±6.0 | 18.95 ±4.8 | 48.39 ±7.3 | 16.93 ±6.0 | r; p-value | -0.01; 7.66e-01 | NA | NA | **0.14; 2.70e-03** |
| **SRS-2 total t-score** ↗ | *n* | 405 | 362 | 43 | 386 | 237 | | 385 *vs.* 228 | 385 *vs.* 1 | 228 *vs.* 1 | 343 *vs.* 42 |
| | *mean ± sd* | 70.48 ±11.9 | 69.85 ±12.1 | 75.74 ±8.5 | 48.12 ±8.7 | 46.0 ±6.7 | r; p-value | **0.77; 6.43e-82** | NA | NA | **0.15; 3.00e-03** |
| **Full scale IQ** ↘ | *n* | 442 | 392 | 50 | 1 | 273 | | 423 *vs.* 267 | 423 *vs.* 1 | 267 *vs.* 1 | 374 *vs.* 49 |
| | *mean ± sd* | 96.83 ±20.0 | 101.21 ±16.6 | 62.45 ±6.8 | 129.0 | 108.69 ±12.9 | r; p-value | **-0.31; 6.62e-16** | NA | NA | **-0.55; 4.84e-30** |
| **Verbal IQ** ↘ | *n* | 438 | 389 | 49 | 1 | 273 | | 419 *vs.* 267 | 419 *vs.* 1 | 267 *vs.* 1 | 371 *vs.* 48 |
| | *mean ± sd* | 95.95 ±20.2 | 99.93 ±17.5 | 64.31 ±10.5 | 110.0 | 108.09 ±14.5 | r; p-value | **-0.30; 1.71e-15** | NA | NA | **-0.51; 2.65e-25** |
| **Non verbal IQ** ↘ | *n* | 444 | 395 | 49 | 1 | 273 | | 425 *vs.* 267 | 425 *vs.* 1 | 267 *vs.* 1 | 377 *vs.* 48 |
| | *mean ± sd* | 97.77 ±21.6 | 102.18 ±18.2 | 62.18 ±10.6 | 147.0 | 108.28 ±4.9 | r; p-value | **-0.25; 3.59e-11** | NA | NA | **-0.51; 3.39e-26** |
| **VABS II - ABC score** ↘ | *n* | 382 | 336 | 46 | 0 | 64 | | 364 *vs.* 62 | 364 *vs.* 0 | 62 *vs.* 0 | 319 *vs.* 45 |
| | *mean ± sd* | 70.7 ±15.5 | 73.15 ±13.9 | 52.8 ±15.3 | NA | 103.47 ±13.9 | r; p-value | **-0.55; 8.73e-30** | NA | NA | **-0.42; 1.21e-15** |
| **RBS-R total** ↗ | *n* | 380 | 337 | 43 | 0 | 150 | | 362 *vs.* 145 | 362 *vs.* 0 | 145 *vs.* 0 | 320 *vs.* 42 |
| | *mean ± sd* | 16.12 ±14.2 | 15.47 ±14.1 | 21.21 ±14.3 | NA | 1.33 ±3.0 | r; p-value | **0.68; 7.09e-53** | NA | NA | **0.15; 4.38e-03** |
| **SSP total** ↘ | *n* | 311 | 277 | 34 | 0 | 127 | | 295 *vs.* 123 | 295 *vs.* 0 | 123 *vs.* 0 | 262 *vs.* 33 |
| | *mean ± sd* | 140.44 ±27.8 | 141.88 ±28.1 | 128.74 ±22.2 | NA | 180.22 ±10.9 | r; p-value | **-0.62; 1.90e-34** | NA | NA | **-0.19; 1.53e-03** |

Supplementary Table 2: Clinical table of the InovAND cohort. Continuous variables were compared using the Mann–Whitney–Wilcoxon test due to non-normal distribution associated with rank-biserial correlation for the effect size, while categorical variables were analyzed using the Chi-square ($\chi^2$) test associated with Cramér's V for the effect size. Statistically significant differences between groups were highlighted where applicable. The interpretation of effect sizes is as follows; absolute value near or below 0.1: small effect, absolute value near 0.3: middle effect, absolute value near or greater than 0.5: large effect. A positive effect size indicates that the first group has a higher score than the second group, whereas a negative effect size indicates that the second group has a higher score than the first. The arrows next to the test names indicate the direction in which symptom severity is highest. Note: IDD: intellectual developmental disorder, IQ: intelligence quotient, NA: not applicable, RBS-R: repetitive behavior scale-revised, SRS: social responsiveness scale 2nd edition, SSP: short sensory profile, NT: neurotypical people, VABS II - ABC: vineland adaptative behaviors scale 2nd edition - communication, daily living skills and socialization

| **Clinical data InovAND** | | **Autism** | **Autism without IDD** | **Autism with IDD** | **Relatives** | **Neurotypicals** | | **Probands with autism *vs.* Neurotypical probands** | **Probands with autism *vs.* Relative siblings** | **Neurotypical probands *vs.* Relative siblings** | **Probands with autism without IDD *vs.* Probands with autism with IDD** |
|---|---|---|---|---|---|---|---|---|---|---|---|
| **Sex ratio** | *n* | 1569 | 550 | 477 | 1854 | 409 | | 1271 *vs.* 301 | 1271 *vs.* 321 | 301 *vs.* 321 | 340 *vs.* 416 |
| | *M:F* | 3.04:1 | 2.02:1 | 3.08:1 | 0.97:1 | 0.92:1 | Cramér's V; p-value | **0.28; 1.02e-27** | **0.29; 2.61e-31** | 0.02; 6.16e-01 | **0.08; 2.43e-02** |
| **Age at inclusion** | *n* | 1239 | 520 | 226 | 872 | 379 | | 991 *vs.* 272 | 991 *vs.* 201 | 272 *vs.* 201 | 315 *vs.* 199 |
| | *mean ± sd* | 12.41 ±9.5 | 16.03 ±11.0 | 13.64 ±7.6 | 38.78 ±17.3 | 24.7 ±13.9 | r; p-value | **-0.39; 1.07e-42** | **-0.18; 1.34e-09** | **-0.33; 1.45e-12** | **-0.12; 7.09e-03** |
| **SRS-2 total t-score ↗** | *n* | 440 | 358 | 65 | 528 | 251 | | 254 *vs.* 184 | 254 *vs.* 122 | 184 *vs.* 122 | 198 *vs.* 47 |
| | *mean ± sd* | 72.85 ±12.8 | 72.96 ±12.8 | 74.94 ±11.8 | 46.58 ±9.7 | 43.87 ±5.8 | r; p-value | **0.84; 1.06e-68** | **0.69; 8.32e-41** | **0.16; 4.09e-03** | 0.04; 5.53e-01 |
| **Full scale IQ ↘** | *n* | 741 | 417 | 295 | 367 | 234 | | 562 *vs.* 178 | 562 *vs.* 87 | 178 *vs.* 87 | 263 *vs.* 270 |
| | *mean ± sd* | 76.8 ±34.6 | 100.93 ±18.7 | 41.07 ±18.2 | 112.55 ±15.5 | 108.93 ±16.2 | r; p-value | **-0.49; 6.16e-41** | **-0.42; 7.18e-27** | 0.08; 1.90e-01 | **-0.86; 1.11e-88** |
| **Verbal IQ ↘** | *n* | 575 | 416 | 124 | 57 | 58 | | 413 *vs.* 54 | 413 *vs.* 18 | 54 *vs.* 18 | 269 *vs.* 111 |
| | *mean ± sd* | 91.69 ±29.9 | 102.86 ±24.0 | 55.98 ±18.8 | 114.16 ±19.0 | 108.34 ±21.1 | r; p-value | **-0.24; 2.16e-07** | **-0.19; 7.38e-05** | 0.13; 2.83e-01 | **-0.71; 2.12e-43** |
| **Non verbal IQ ↘** | *n* | 722 | 469 | 216 | 350 | 141 | | 520 *vs.* 107 | 520 *vs.* 90 | 107 *vs.* 90 | 293 *vs.* 190 |
| | *mean ± sd* | 86.37 ±24.7 | 97.76 ±19.7 | 59.96 ±11.4 | 112.09 ±16.1 | 109.28 ±17.4 | r; p-value | **-0.37; 1.02e-20** | **-0.38; 1.68e-20** | 0.03; 7.05e-01 | **-0.77; 5.01e-65** |
| **VABS II - ABC score ↘** | *n* | 91 | 74 | 17 | 8 | 26 | | 89 *vs.* 26 | 89 *vs.* 2 | 26 *vs.* 2 | 72 *vs.* 17 |
| | *mean ± sd* | 70.54 ±11.9 | 72.81 ±11.3 | 60.65 ±9.5 | 93.62 ±18.4 | 103.65 ±9.1 | r; p-value | **-0.70; 5.42e-14** | NA | NA | **-0.40; 1.38e-04** |
| **RBS-R total ↗** | *n* | 302 | 217 | 67 | 486 | 197 | | 245 *vs.* 159 | 245 *vs.* 118 | 159 *vs.* 118 | 174 *vs.* 59 |
| | *mean ± sd* | 24.68 ±19.4 | 23.8 ±19.6 | 28.42 ±18.5 | 4.07 ±8.3 | 2.18 ±3.8 | r; p-value | **0.75; 2.90e-52** | **0.65; 4.70e-35** | **0.12; 3.10e-02** | 0.10; 1.37e-01 |
| **SSP total ↘** | *n* | 194 | 153 | 39 | 208 | 130 | | 158 *vs.* 114 | 158 *vs.* 47 | 114 *vs.* 47 | 120 *vs.* 37 |
| | *mean ± sd* | 143.43 ±27.9 | 144.27 ±29.2 | 139.95 ±22.5 | 177.77 ±17.4 | 180.67 ±19.4 | r; p-value | **-0.65; 2.36e-23** | **-0.43; 1.82e-09** | **-0.21; 2.21e-02** | -0.01 9.20e-01 |

Supplementary Table 3: Clinical table comparison between the autistic and neurotypical probands between both cohorts. Continuous variables were compared using the Mann–Whitney–Wilcoxon test due to non-normal distribution associated with rank-biserial correlation for the effect size, while categorical variables were analyzed using the Chi-square ($\chi^2$) test associated with Cramér's V for the effect size. Statistically significant differences between groups were highlighted where applicable. The interpretation of effect sizes is as follows; absolute value near or below 0.1: small effect, absolute value near 0.3: middle effect, absolute value near or greater than 0.5: large effect. A positive effect size indicates that the first group has a higher score than the second group, whereas a negative effect size indicates that the second group has a higher score than the first. The arrows next to the test names indicate the direction in which symptom severity is highest. Note: IDD: intellectual developmental disorder, IQ: intelligence quotient, NA: not applicable, RBS-R: repetitive behavior scale-revised, SRS: social responsiveness scale 2nd edition, SSP: short sensory profile, NT: neurotypical people, VABS II - ABC: vineland adaptative behaviors scale 2nd edition - communication, daily living skills and socialization

| | | Autistic probands | | Neurotypical probands | | | LEAP vs. InovAND | |
|---|---|---|---|---|---|---|---|---|
| **Clinical data** | | **LEAP** | **InovAND** | **LEAP** | **InovAND** | | **Autistic probands** | **Neurotypical probands** |
| **Sex ratio** | *n* | 458 | 1271 | 291 | 301 | | 458 *vs.* 1271 | 291 *vs.* 301 |
| | *M:F* | 2.58:1 | 4.73:1 | 1.83:1 | 1.12:1 | Cramér's V; p-value | **0.11; 2.38e-06** | **0.12; 4.71e-03** |
| **Age at inclusion** | *n* | 433 | 911 | 268 | 272 | | 433 *vs.* 911 | 268 *vs.* 272 |
| | *mean ±sd* | 16.87 ±5.9 | 11.89 ±8.7 | 17.01 ±6.0 | 22.34 ±12.4 | r; p-value | **0.39; 2.10e-49** | **-0.18; 2.16e-05** |
| **SRS-2 total t-score** | *n* | 385 | 254 | 228 | 184 | | 385 *vs.* 254 | 228 *vs.* 184 |
| | *mean ±sd* | 70.5 ±12.0 | 74.98 ±11.6 | 45.88 ±6.4 | 44.14 ±5.7 | r; p-value | **-0.17; 1.62e-05** | **0.17; 7.00e-04** |
| **Full scale IQ** | *n* | 423 | 562 | 267 | 178 | | 423 *vs.* 562 | 267 *vs.* 178 |
| | *mean ±sd* | 96.66 ±20.0 | 70.09 ±34.4 | 108.79 ±13.0 | 108.72 ±16.0 | r; p-value | **0.39; 4.36e-35** | -0.01; 8.79e-01 |
| **Verbal IQ** | *n* | 419 | 413 | 267 | 54 | | 419 *vs.* 413 | 267 *vs.* 54 |
| | *mean ±sd* | 95.78 ±20.2 | 86.6 ±29.9 | 108.13 ±14.6 | 108.31 ±20.5 | r; p-value | **0.16; 4.31e-06** | -0.03; 6.27e-01 |
| **Non verbal IQ** | *n* | 520 | 425 | 520 | 107 | | 520 *vs.* 425 | 520 *vs.* 107 |
| | *mean ±sd* | 97.58 ±21.6 | 83.2 ±25.0 | 108.44 ±15.0 | 108.41 ±17.2 | r; p-value | **0.29; 2.16e-19** | -0.00; 9.56e-01 |
| **VABS II - ABC score** | *n* | 364 | 89 | 62 | 26 | | 364 *vs.* 89 | 62 *vs.* 26 |
| | *mean ±sd* | 70.74 ±15.8 | 70.67 ±11.9 | 104.26 ±13.1 | 103.65 ±9.1 | r; p-value | 0.01; 7.85e-01 | 0.02; 8.80e-01 |
| **RBS-R total** | *n* | 362 | 245 | 145 | 159 | | 362 *vs.* 245 | 145 *vs.* 159 |
| | *mean ±sd* | 15.94 ±14.2 | 26.71 ±19.4 | 1.18 ±2.1 | 2.42 ±4.0 | r; p-value | **-0.31; 4.31e-14** | -0.10; 6.10e-02 |
| **SSP total** | *n* | 295 | 158 | 123 | 114 | | 295 *vs.* 158 | 123 *vs.* 114 |
| | *mean ±sd* | 140.77 ±28.0 | 141.28 ±26.8 | 180.63 ±9.7 | 179.68 ±20.7 | r; p-value | -0.01; 9.09e-01 | **-0.14; 3.36e-02** |

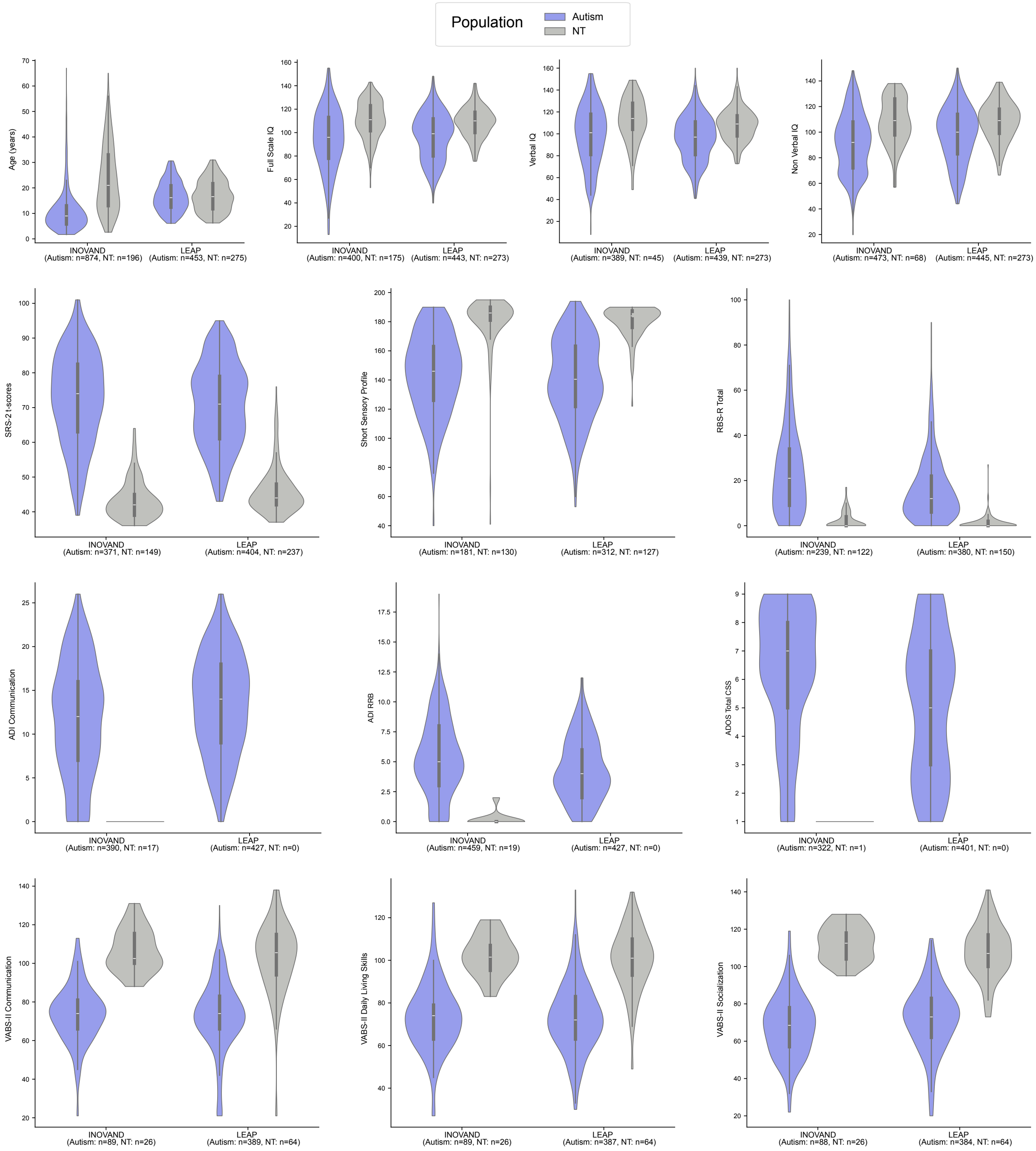

Supplementary Figure 3: Distribution of clinical profiles from both autistic and NT people between the LEAP and InovAND cohorts. VABS-II = Vineland Adaptive Behavior Scales, 2nd Edition; SRS-2 = Social Responsiveness Scale, 2nd Edition; ADI = Autism Diagnostic Interview; ADOS = Autism Diagnostic Observation Schedule; RBS-R = Repetitive Behavior Scale – Revised.

### Section 3. Genetics

*2.1 Whole genome sequencing*

DNA was extracted from whole blood/saliva and was sequenced at a read depth of 30×. A total of 2,560 samples were sequenced by the CNRGH using Illumina platforms, including 1,314 on HiSeq and 1,246 on NovaSeq. Additionally, 99 samples were sequenced using Illumina platforms in other facilities, and 17 samples were sequenced by NGXBio. Reads were aligned to GRCh37[10 11](v.20240522) and tagged, sorted and converted to CRAM format using samblaster[12] (v.0.1.26), sambamba[13] (v.0.8.0) and samtools[14] (v.1.14) (**Supplementary Fig. 4**).

***SNV and Indel***

Individual gVCF were then called using DeepVariant [15] (v.1.8.0) and genotyped jointly using GInexus [16,17] (v.1.4.1). Variants were normalized and left aligned using bcftools [12] (v.1.12). Individual variants were filtered using a depth of coverage (DP) ≥10 and genotype quality (GQ) ≥20. We filtered out variants occurring in low complexity regions (LCR). We also required heterozygous variants to have a variant allele frequency (VAF) in [0.25 0.75]. Detected single nucleotide variants (SNVs)/indel of interest were validated by visualization of the CRAM files and then by Sanger sequencing.

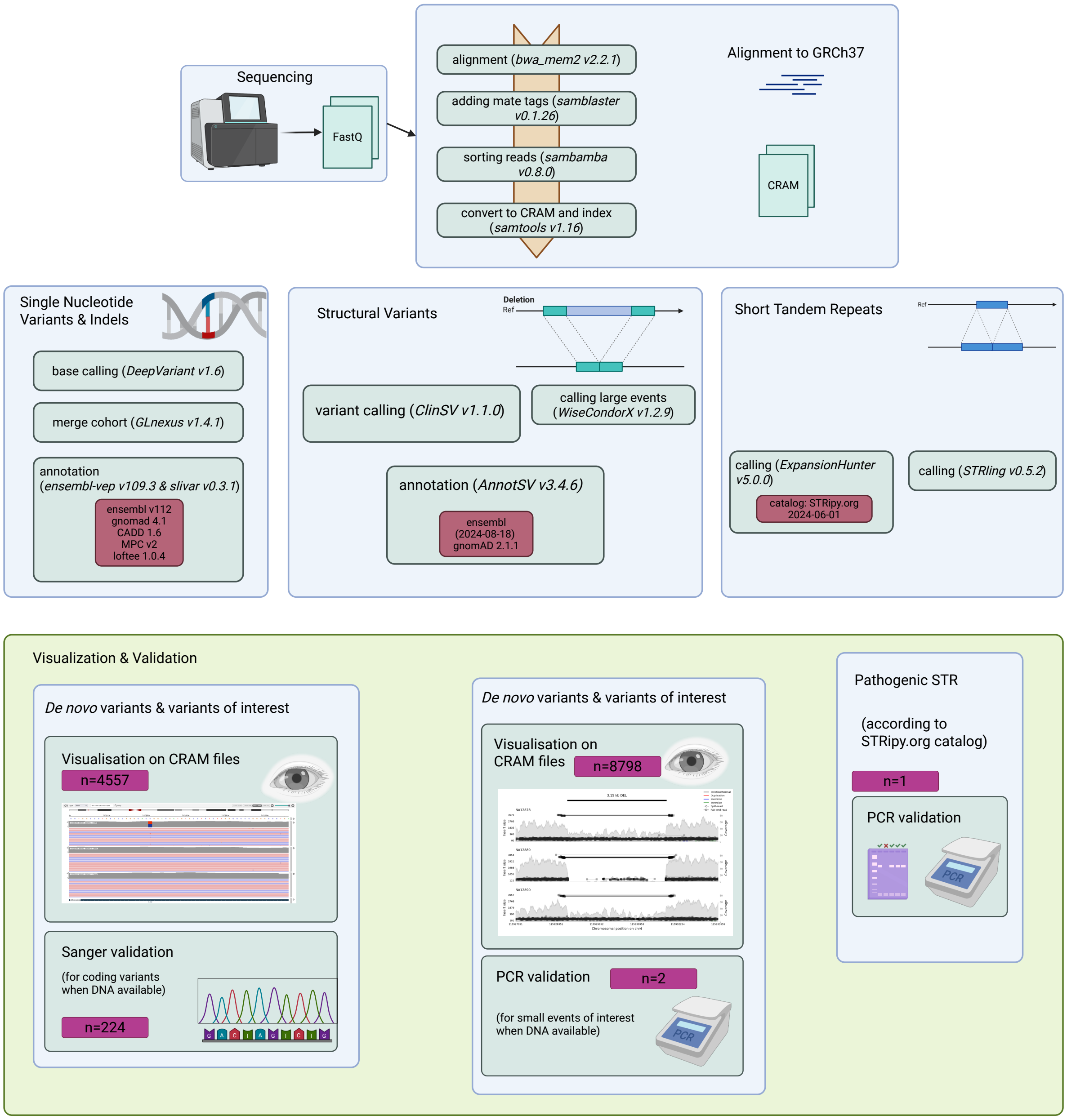

Supplementary Figure 4: Pipeline from whole genome sequencing to detection of the variants.

#### ***Structural variants***

In total, 282 people were excluded from the structural variants (SV) and copy-number variants (CNV) analysis due to highly variable levels of waviness in their WGS coverage. To classify samples as "wave" or "not wave," we used a supervised machine learning approach with a Random Forest classifier (scikit-learn v1.3, Python v.3.8.18). The model was trained on a labeled dataset containing genomic features (*e.g.*, read depth, variant counts, GC content, coverage uniformity) and then used to predict labels for unlabeled samples. All features were standardized, and a fixed random seed ensured reproducibility.

For SV detection, we first ran WisecondorX [18] (v.1.2.9) using default parameters and with a window size of 5kb to improve the detection of large events (aneuploidy & SVs size > 10Mb). A subset of 424 people from NT families have been used as a coverage calibration dataset to predict SV. Only WisecondorX deletion with a ratio ≤ -0.1 and WisecondorX duplication with a ratio ≥ 0.2 were kept. Recurrent SV with an overlap of 90% with SPARK loci were identified using bedtools [19,20] (v.2.30.0) and included in the list of SV associated with NDD. SPARK loci are annotated by “Dosage Sensitivity Expert Panel” from ClinGen (Clinical Genome resource) and only deletions with sufficient evidence for haploinsufficiency and duplications with sufficient evidence for haploinsufficiency were selected. Filtered WisecondorX SV were validated by visualization using WisecondorX plot or on CRAM.

We then used ClinSV [46] (v.1.1.0) with default parameters and filtered variants based on confidence: High (large size and/or high AMQ), Pass (DOC deviation > 20% and sufficient DP+SR), Balanced (DP+SR ≥ 6), and Low (all others), where DP = discordant pairs, SR = split reads, DOC = depth of coverage, and AMQ = average mapping quality. A total of 2,189 variants were manually visualized and further used to create a Random Forest model to discriminate true SV from noise. This model relies on ClinSV and provides score in addition to median depth information within the SV, in its 10kb flanking regions and in its chromosome. SV were annotated using AnnotSV [47] (v.3.4.6). We excluded 282 samples as outliers in number of events (> 1.5 * IQR) most likely due to DNA quality issue. Detected CNVs (> 8,000) were then validated by visualization on CRAM. Detected CNVs of interest (> 8,000) were then validated by visualization of the CRAM files and then validated using PCR and Sanger sequencing.

***Short Tandem Repeats***

For STR, we ran ExpansionHunter [21] (v.5.0.0) using the loci dictionary from stripy.org [22] (v.1.2), which includes 52 loci previously identified as pathogenic. We also reinforced the results by running STRling [23] using its outliers' script (v.0.5.2) and validated using targeted PCR.

##### *2.3 Pipelines of genetic analyses for returnable clinically relevant variants*

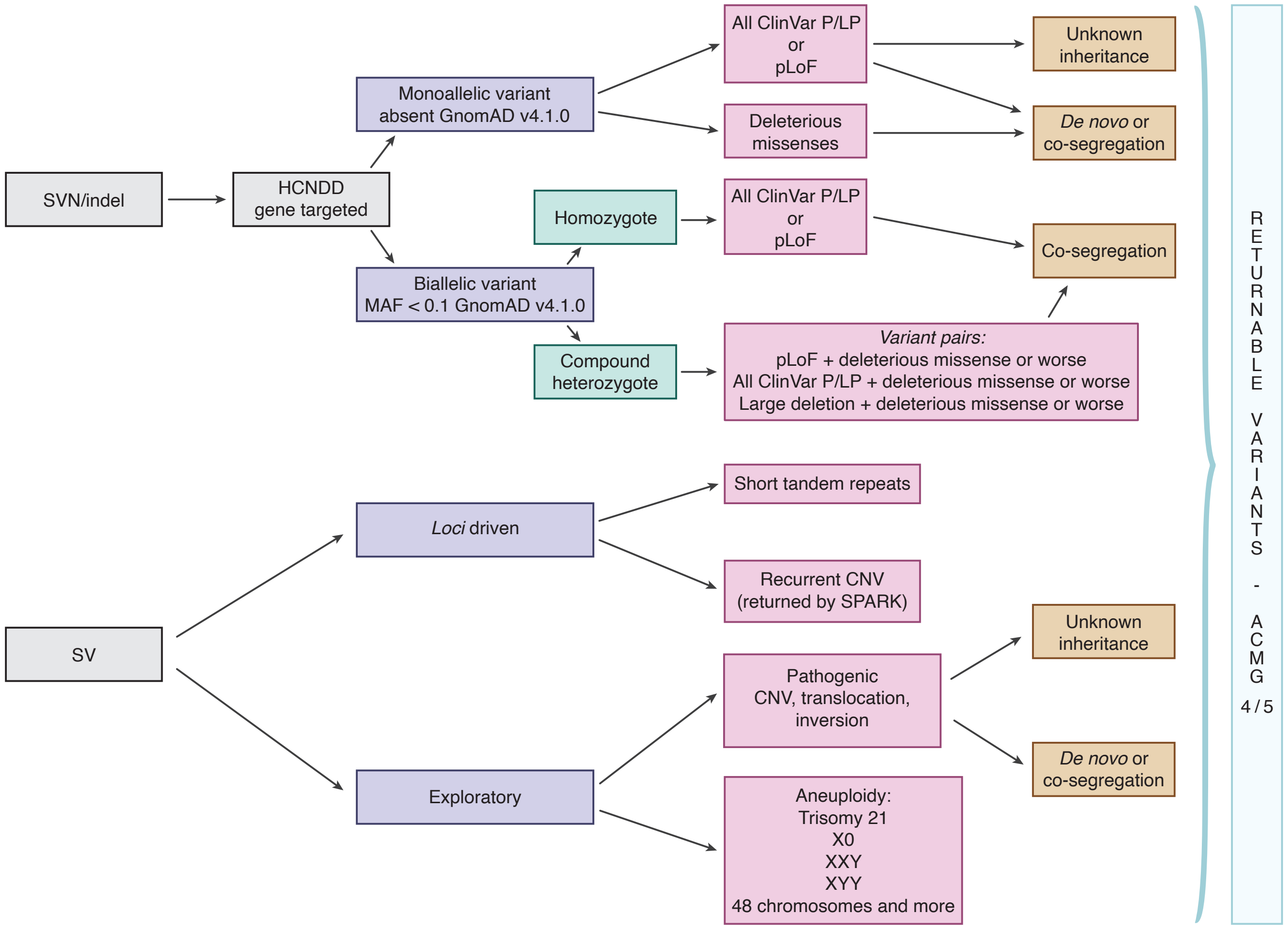

Supplementary Figure 5: Pipeline for detection of clinically relevant variants. ACMG: American college of medical genetics and genomics, ClinVar LP/P (accessed 2025-06): ClinVar likely pathogenic and pathogenic, CNV: copy-number variant, SV: structural variants, GnomAD: genome aggregation database, HCNDD: high confidence NDD, MAF: minor allele frequency, pLoF: protein-truncating loss of function, STR: short tandem repeats.

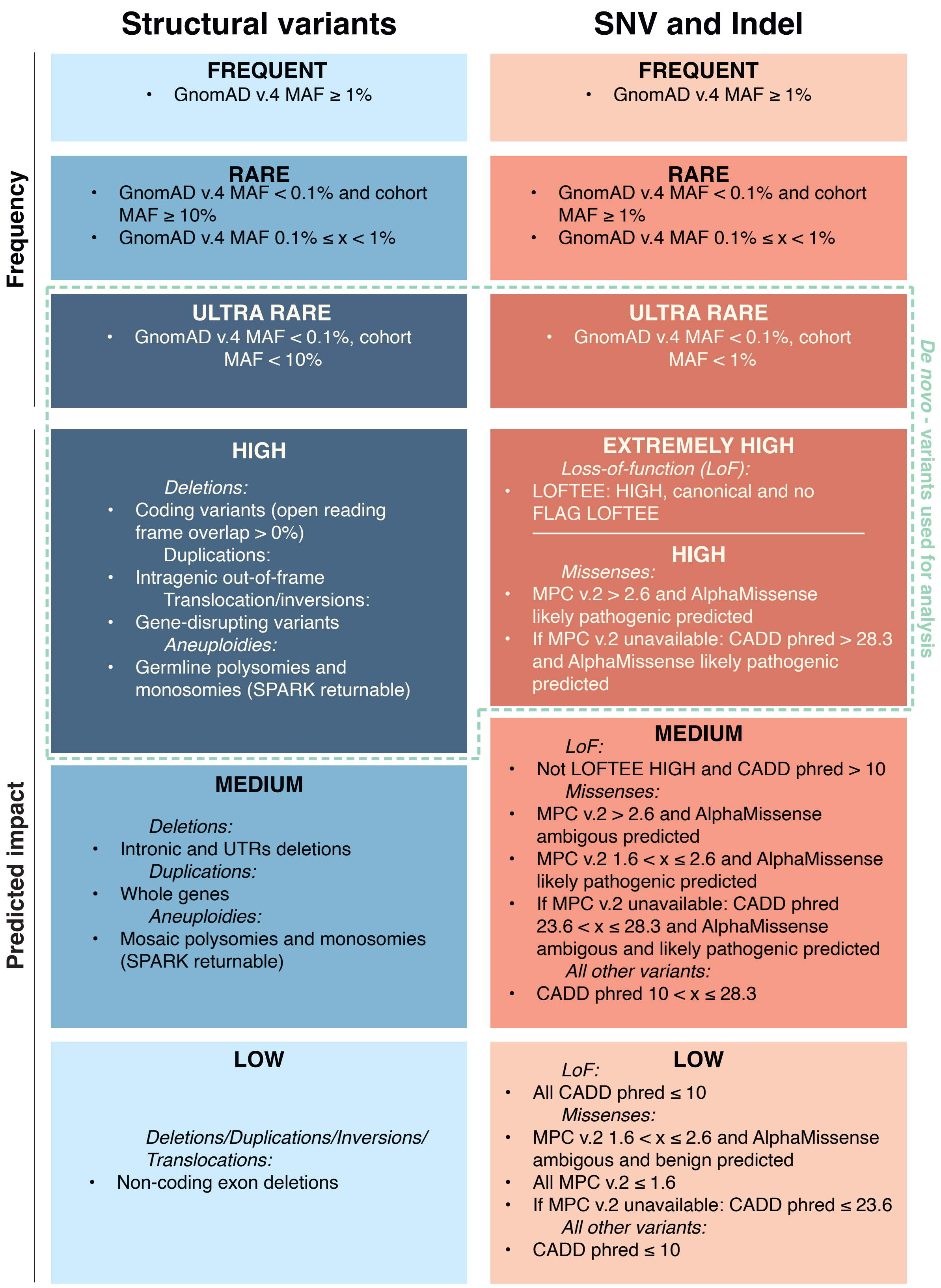

Supplementary Figure 6: Schematic representation of the classification of variants in this research study. The classification is based on the event type (structural variants (SV)/copy number variants (CNV) *vs.* single nucleotide variants (SNV)/indel), the pathogenicity prediction and the frequency in the general population.

*2.2 The ancestry clustering of LEAP-InovAND*

We used Somalier [24] (v.0.2.19) to confirm familial relationships and person's sex. We also used the Somalier extracted SNPs to compute the admixture of the cohort along with the Human Genome Diversity Project [25] (neural-admixture v.1.4.1). We then predicted the superpopulation (EUR, AFR, EAS, CSA, MID, OCE, AME) with a Random Forest model using admixture results and known data from the Human Genome Diversity Project. We left people admix between ancestries (probability of belonging to a single ancestry $<0.5$) as unknown (UNK) (**Supplementary Figs. 7-8**).

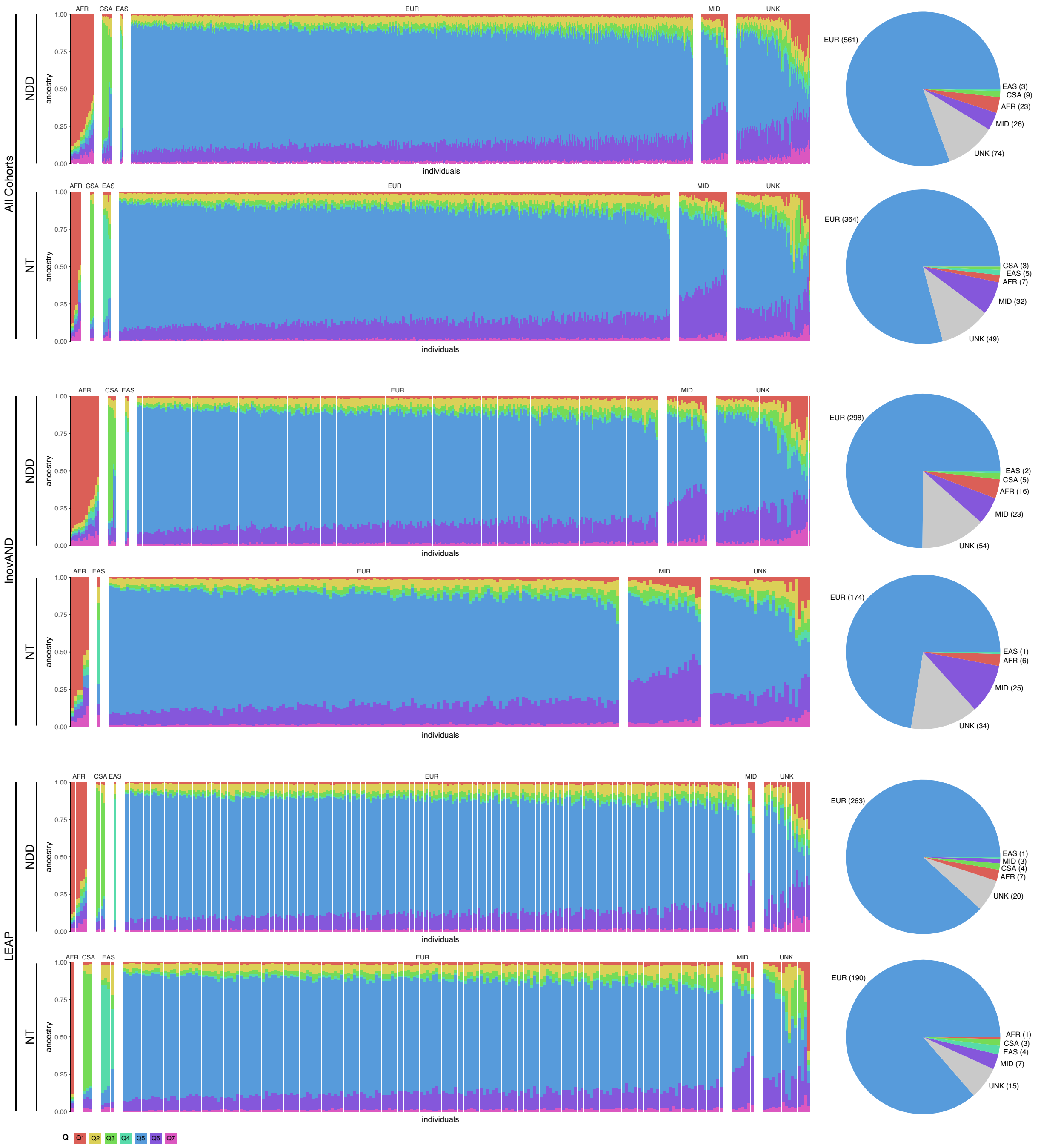

Supplementary Figure 7: Ancestral composition of people in both cohorts based on admixture analysis. Each bar represents one individual, and the colored segments indicate the proportion of genetic ancestry attributed to different ancestral populations as inferred by the admixture model. People are grouped according to their predominant ancestry component.

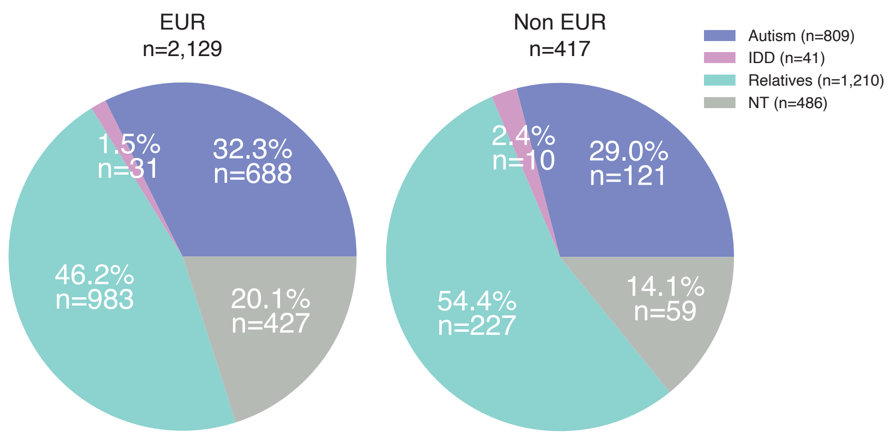

Supplementary Figure 8: Pie charts representing the distribution of population across the European and non-European ancestries

#### *2.4 De novo genetic variations*

For *de novo* variants, we additionally filtered then requiring a cumulated parent allelic depth (AD) < 2 and a cohort allele count (AC) < 3 (tolerating twins or germinal mosaicism). The coding variants and a random selection of non-coding variants were visualized on CRAM and used to feed a Random Forest method to predict their likelihood of being true *de novo* variants. Variants were annotated with VEP [26] (v.109.3) using the Ensembl database (r.112). We also used LOFTEE [27] (v.1.0.4) to assess the loss-of-function variants, gnomAD [28] (v.4.1) for population frequency, MPC [29] (v.2) for missense deleteriousness, CADD [30] (v.1.6) for general deleteriousness of variants. These filters correspond to the extremely high and high impact ultra-rare variants (hiURV) (**Supplementary Fig. 6**).

In 377 families with parental WGS data available, we could perform a *de novo* analysis on 371 people with autism, 146 undiagnosed siblings, 30 NT people, and 14 with IDD. These *de novo* variants, which are not detected in the parents' DNA samples, typically originate in the parental germline, predominantly paternal (75–81%), and early in embryonic development (~16%) [21]. On average, in this study, each participant had 77 *de novo* SNVs (range 37-126). Consistent with previous studies [31], we observed a positive correlation between paternal age and the number of *de novo* SNVs, with no significant excess in people with autism relative to their siblings (**Supplementary Fig. 9**).

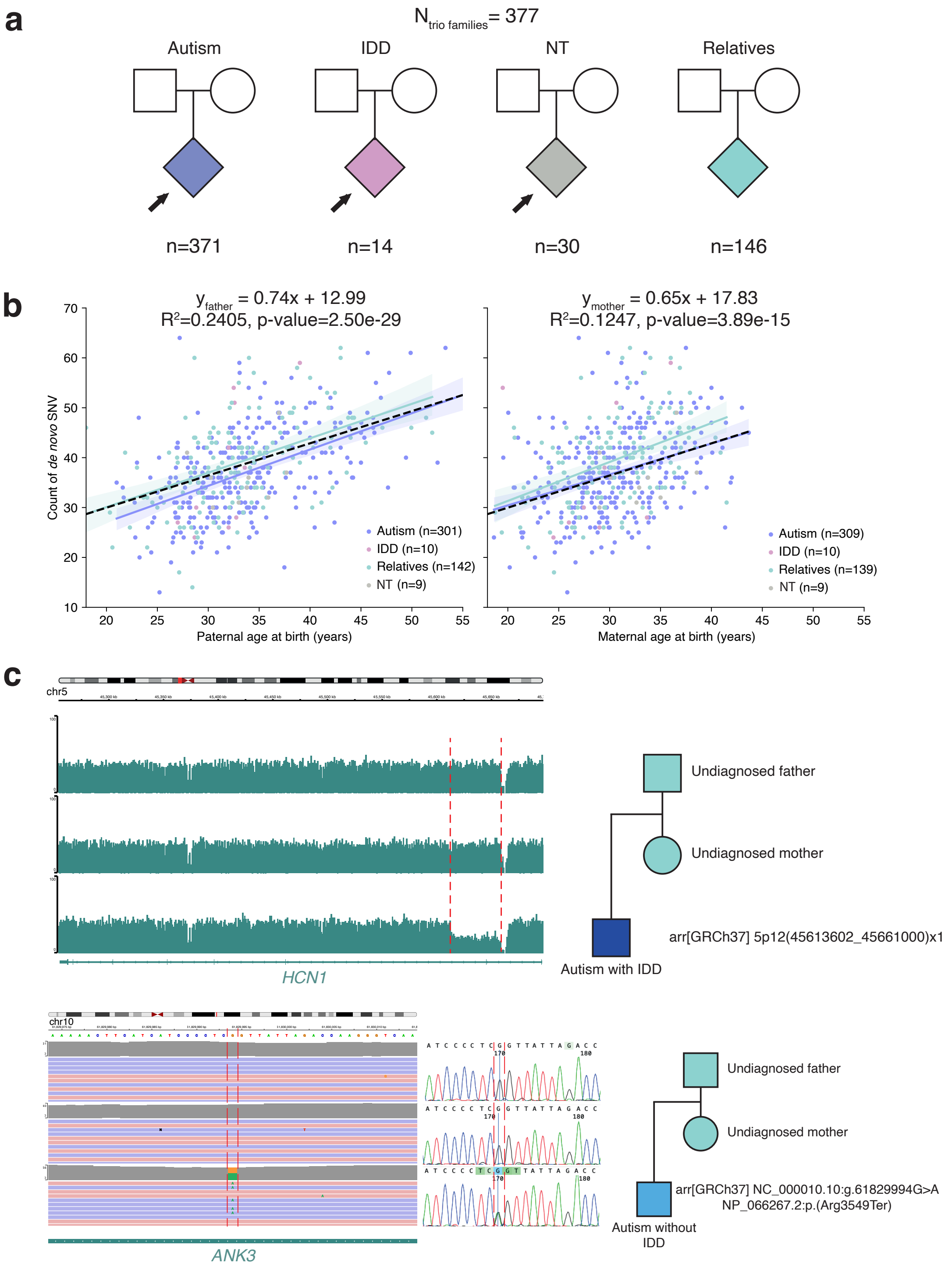

Supplementary Figure 9: Examples of *de novo* genetic variants identified in autistic participants. **a**. Schematic description of the family count in the *de novo* study, in addition to the participant count by phenotype. **b**. Correlation between parental age at birth (father on the left, mother on the right) and the number of *de novo* single nucleotide variants (SNV) detected. Regression curves and their corresponding equations are shown. **c**. Example of a *de novo* copy number variant (CNV): a deletion affecting the *HCN1* gene (removing one out-of-frame coding exon) found in an autistic person with IDD, born from non-autistic parents. **d**. Example of a *de novo* SNV: a nonsense variant in the *ANK3* gene found in an autistic person without IDD, born from non-autistic parents.

*2.5 Ultra rare variants in LEAP-InovAND*

We analyzed high impact ultra rare variants (hiURV) in gene sets related to autism/NDD using different criteria regarding the genetic variants (deletions and loss of functions or deleterious missense variants), for tolerance to loss of function variants and for European ancestry or all ancestry (multi-ancestry). Missense variants are considered deleterious according to MPC score (v2) and AlphaMissense [32]. The notion of gene constraint is based on the union of GnomAD v4 for autosomes, GnomAD v2 for X chromosome and sHET [33]. The HCNDD gene list is based on various NDD databases [34] updated with the PanelApp England and Australia (**Supplementary Data 1**). The SPARK/SFARI1 gene list is the union of genes returned by SPARK and genes strongly associated with autism according to SFARI [35]. The EAGLE definitive and strong gene list is the union of EAGLE genes classified as “Definitive” and “Strong”, which is a curated list of genes associated with autism [36]. The SynGO gene list corresponds to the Synapses Gene Ontology gene list [37]. Finally, the ChromEpiTF gene list is the union of genes described in chromatin remodeling [38], the Epifactors gene list [39], and two transcription factor databases (Rummagene [40] and TRRUST [41]).

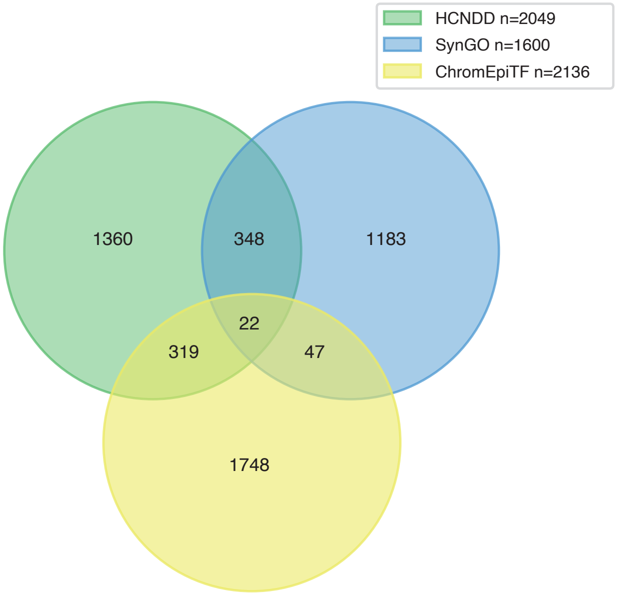

Supplementary Figure 10: Venn diagram showing the counts of HCNDD, SynGO and ChromEpiTF genes.

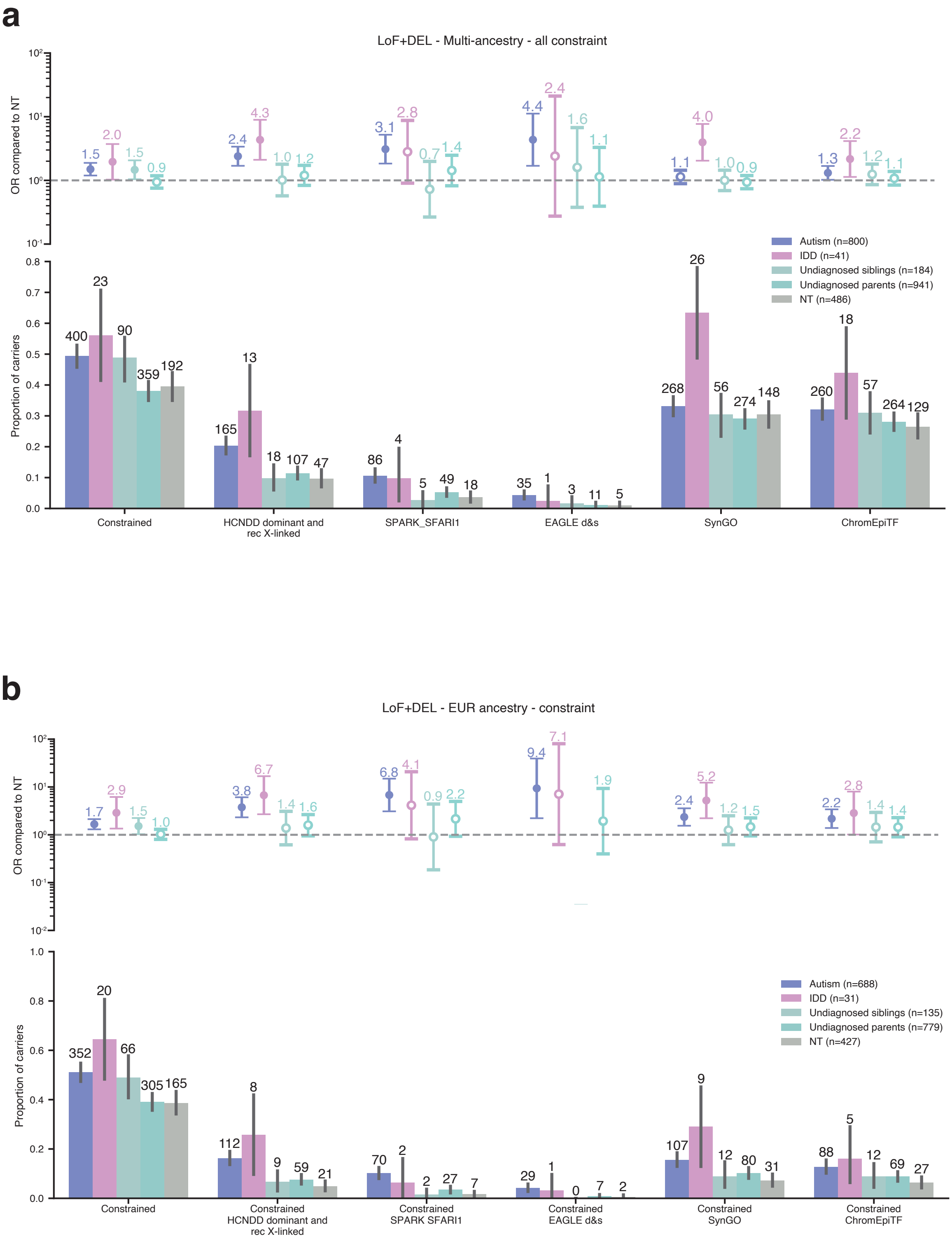
a
LoF+DEL - Multi-ancestry - all constraint
OR compared to NT
Proportion of carriers
Autism (n=800)
IDD (n=41)
Undiagnosed siblings (n=184)
Undiagnosed parents (n=941)
NT (n=486)
Constrained
HCNDD dominant and rec X-linked
SPARK_SFARI1
EAGLE d&s
SynGO
ChromEpiTF
b
LoF+DEL - EUR ancestry - constraint
OR compared to NT
Proportion of carriers
Autism (n=688)
IDD (n=31)
Undiagnosed siblings (n=135)
Undiagnosed parents (n=779)
NT (n=427)
Constrained
Constrained HCNDD dominant and rec X-linked
Constrained SPARK SFARI1
Constrained EAGLE d&s
Constrained SynGO
Constrained ChromEpiTF

c

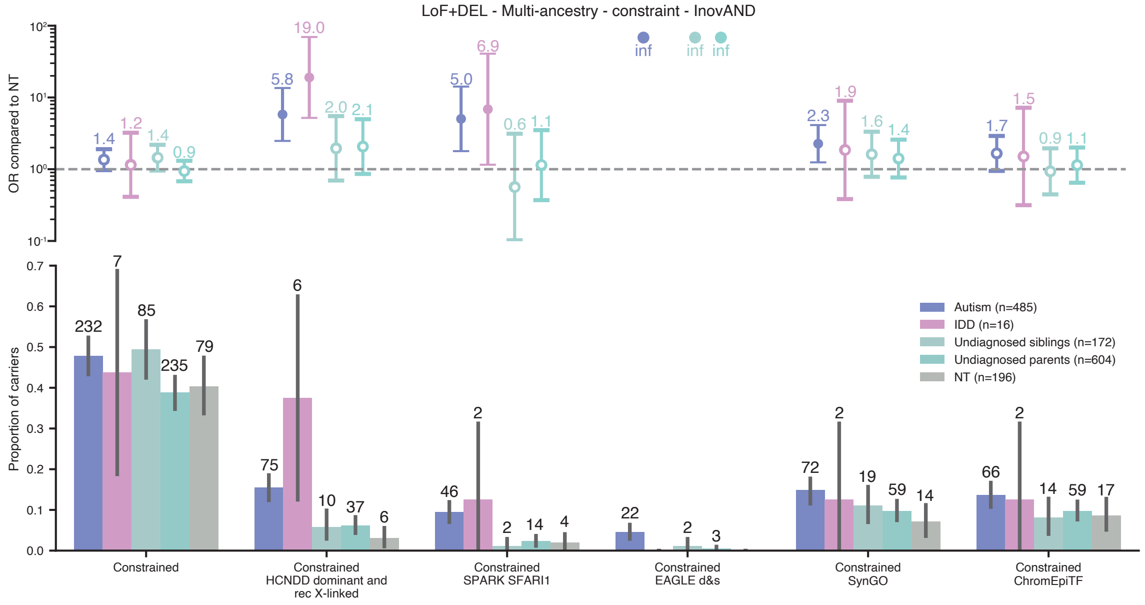
LoF+DEL - Multi-ancestry - constraint - InovAND
OR compared to NT
Proportion of carriers
Autism (n=485)
IDD (n=16)
Undiagnosed siblings (n=172)
Undiagnosed parents (n=604)
NT (n=196)
Constrained
Constrained HCNDD dominant and rec X-linked
Constrained SPARK SFARI1
Constrained EAGLE d&s
Constrained SynGO
Constrained ChromEpiTF

d

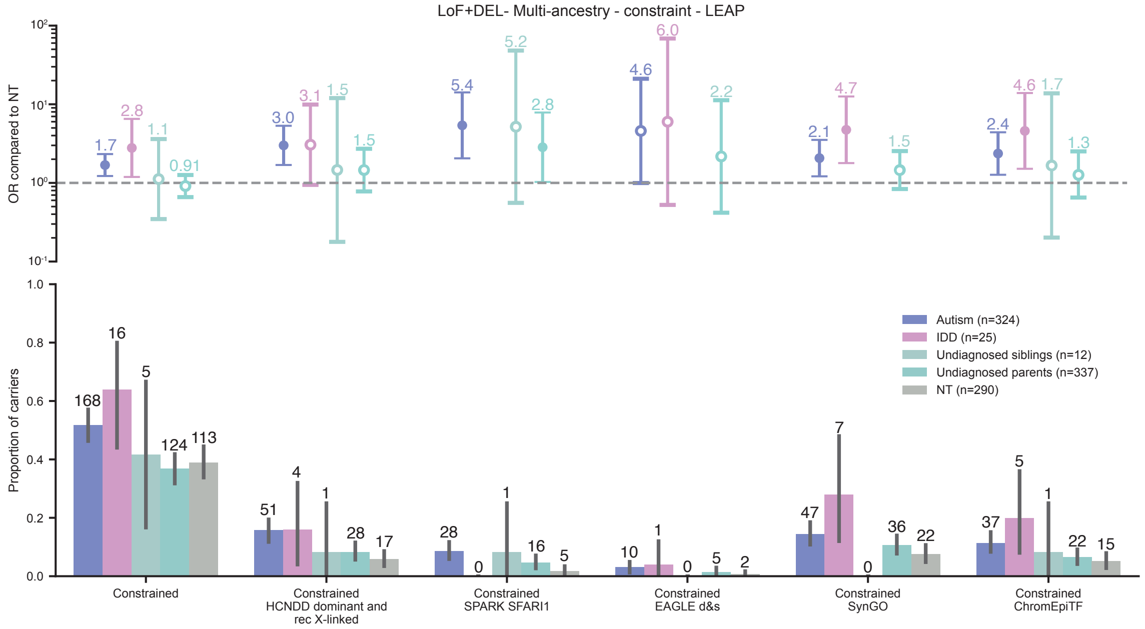
LoF+DEL- Multi-ancestry - constraint - LEAP
OR compared to NT
Proportion of carriers
Autism (n=324)
IDD (n=25)
Undiagnosed siblings (n=12)
Undiagnosed parents (n=337)
NT (n=290)
Constrained
Constrained HCNDD dominant and rec X-linked
Constrained SPARK SFARI1
Constrained EAGLE d&s
Constrained SynGO
Constrained ChromEpiTF

**e**

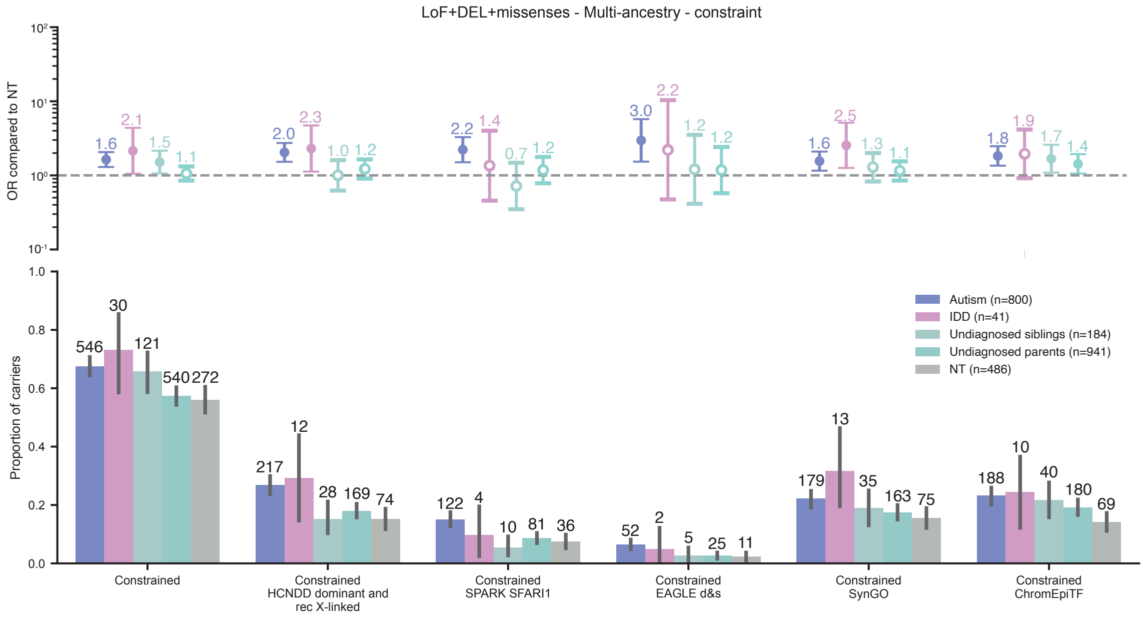

LoF+DEL+missenses - Multi-ancestry - constraint
OR compared to NT
Proportion of carriers
Autism (n=800)
IDD (n=41)
Undiagnosed siblings (n=184)
Undiagnosed parents (n=941)
NT (n=486)
Constrained
Constrained HCNDD dominant and rec X-linked
Constrained SPARK SFARI1
Constrained EAGLE d&s
Constrained SynGO
Constrained ChromEpiTF

**f**

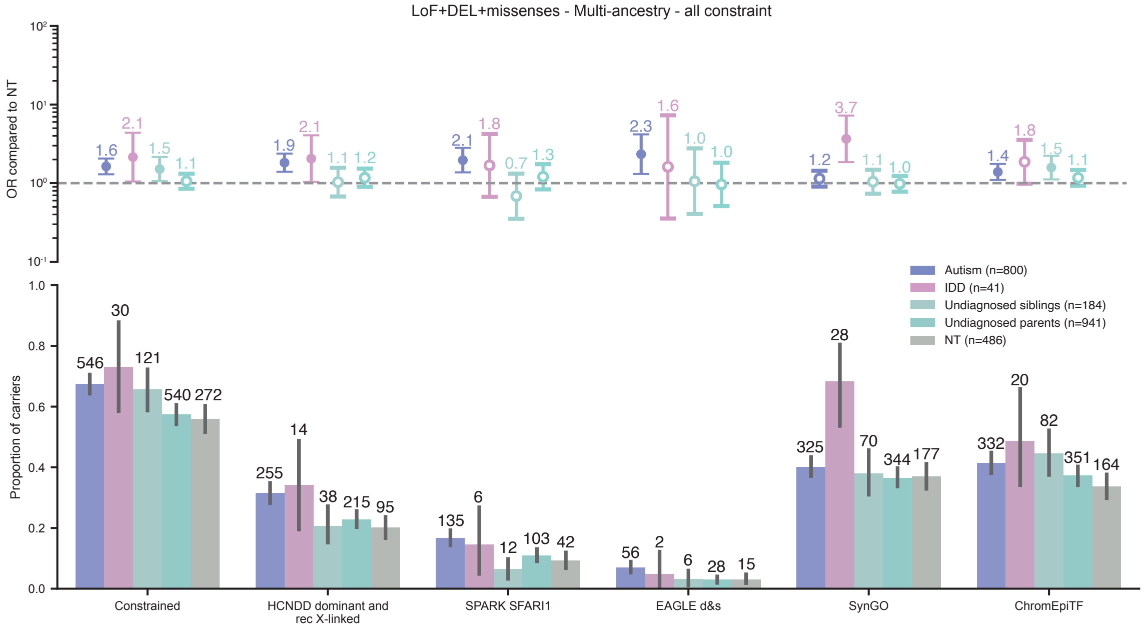

LoF+DEL+missenses - Multi-ancestry - all constraint
OR compared to NT
Proportion of carriers
Autism (n=800)
IDD (n=41)
Undiagnosed siblings (n=184)
Undiagnosed parents (n=941)
NT (n=486)
Constrained
HCNDD dominant and rec X-linked
SPARK SFARI1
EAGLE d&s
SynGO
ChromEpiTF

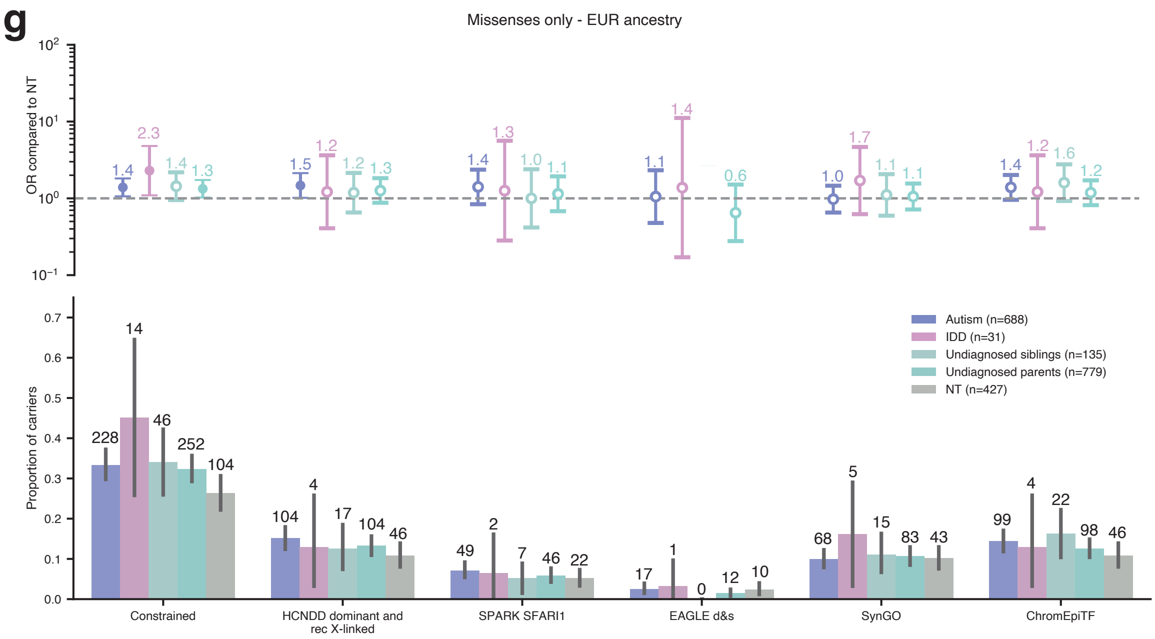

Supplementary Figure 11: **Analyses of hiURV in gene sets related to autism/NDD**. **a.** Deletions and loss of functions: multi-ancestries and without gene constrained filter. **b,** only in the population with European ancestry. **c,** multi-ancestries, constrained genes and only in InovAND participants. **d,** multi-ancestries, constrained genes and only in LEAP participants. **e,** deletions, missenses considered deleterious according to MPC score (v2) and AlphaMissense, and loss of functions: multi-ancestries, constrained genes. **f**. same as e. but with constrained and not constrained genes. **g**, missenses considered deleterious according to MPC score (v2) and AlphaMissense only: multi-ancestries and without gene constrained filter. OR (with CI95%) for each condition have been calculated from NT. The circle is filled in when the difference is significant without correction for multiple testing.

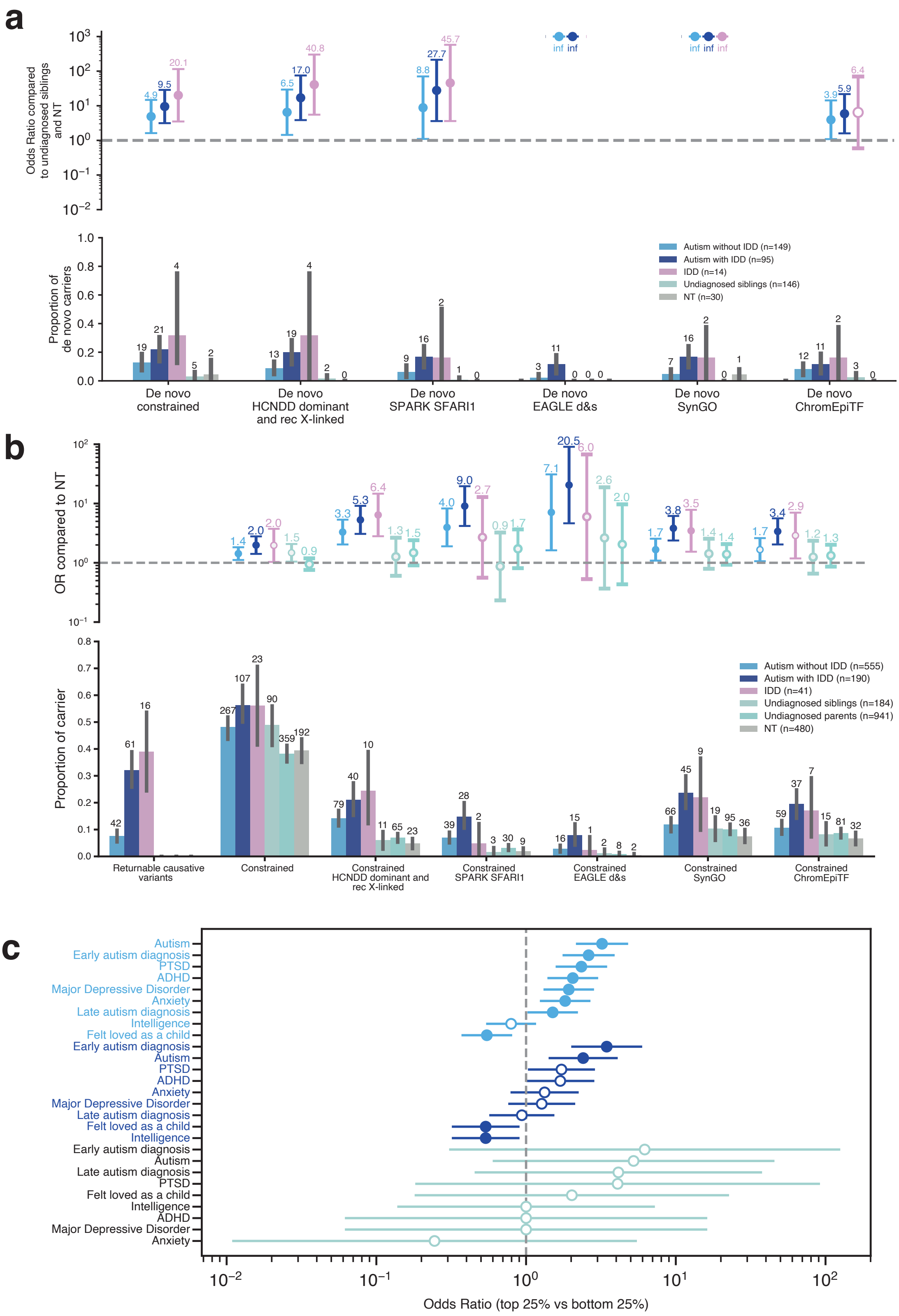

Supplementary Figure 12: **Genetic profile of the participant (with IDD phenotype) of the LEAP-InovAND cohort. a**. Genetic study of *de novo* variants (deletions, loss of function and deleterious missenses) in both cohorts (multi-ancestry). The bottom panel represents the proportion of *de novo* variant carriers according to the different gene lists used. The figures represent the number of carriers per population and per gene list. The top panel represents the OR based on the proportions of the bottom panel. For *de novo* variations, odd ratio (OR) are calculated by comparing the frequency of *de novo* variants in autistic or IDD people to the pooled frequency in undiagnosed siblings and NT. **b.** Genetic study of returnable genetic diagnosis and predicted pathogenic variants (deletions and loss of function) in constraint genes in both cohorts (multi-ancestry). For this analysis, ORs (with CI95%) are computed and corrected (FDR, one-sided independent t-tests adjusted for multiple comparisons using Benjamini–Hochberg correction, difference is significant when the circle is full) compared to that of NT**. c.** PGS for different GWAS compared to NT, representing the OR for the top 25% (Q1) versus the bottom 25% (Q4) of the PGS distribution. The difference is significant when the circle is full.

By using predicted deleterious deletions and LoF variants affecting constrained genes, we aimed to identify possible specific biological pathways or cellular signatures described in the articles by Litman *et al*. and Herring *et al.* [42,43]. To do this, we calculated the odds ratio of autistic people across the three different clusters, using the NT people from the entire cohort as a reference (**Supplementary Fig. 13**).

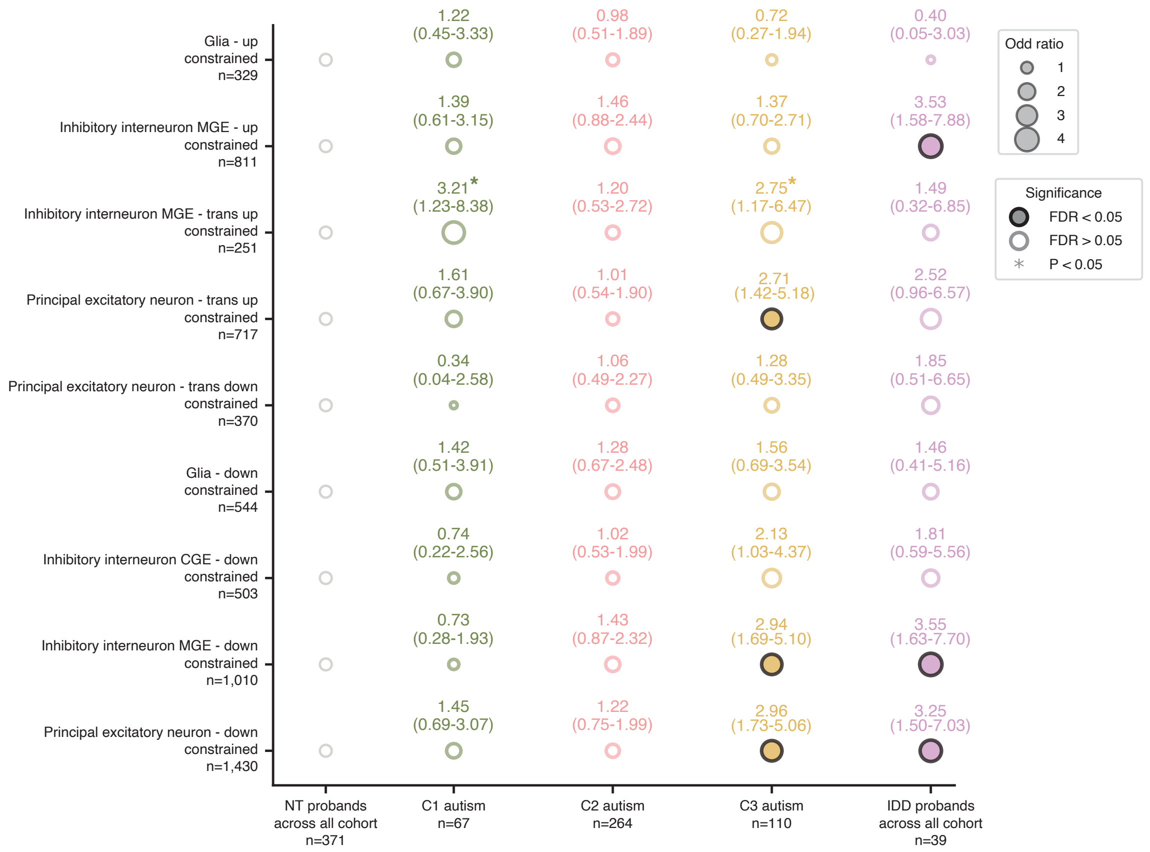

Supplementary Figure 13 Patterns of hiURV (*de novo* / inherited LoF/DEL with a MAF<0.1%) variant enrichment across clusters (x axis), major cell types of the prefrontal cortex (y axis) and gene expression trends (y axis). For each cluster, we computed the OR (bubble size) and corrected P values (FDR, one-sided independent t-tests adjusted for multiple comparisons using Benjamini–Hochberg correction) of variant burden compared to that of NT probands cross all cohort. Open circles indicate FDR>0.05 (not significant), and closed circles indicate significant enrichment (FDR≤0.05), asterisk represent significancy before FDR correction.

*2.6 Polygenic scores in LEAP-InovAND*

Various summary statistics for different traits were used, including autism (Grove *et al.* 2019), ADHD (Demontis *et al.* 2023), intelligence (Savage *et al.* 2018) and genetic generalized epilepsy (ILAE 2023). Heritability estimates and cross-trait genetic correlations derived from GWAS data were computed. Consistent with previous findings, autism exhibits robust positive genetic correlations with ADHD (rg = 0.55, $P_{Bonferroni} = 7.0 \times 10^{-28}$), anxiety (rg = 0.38, $P_{Bonferroni} = 3.3 \times 10^{-13}$) major depressive disorder (rg = 0.38, $P_{Bonferroni} = 3.7 \times 10^{-31}$), and intelligence (rg = 0.21, $P_{Bonferroni} = 8.9 \times 10^{-11}$). Conversely, autism demonstrates negative genetic correlations with subjective well-being (rg = –0.48, $P_{Bonferroni} = 7.1 \times 10^{-16}$), the self-reported sense of being "felt loved as a child" (rg = –0.39, $P_{Bonferroni} = 4.1 \times 10^{-13}$), and indicators of social connectedness, including satisfaction with family relationships (rg = –0.31, $P_{Bonferroni} = 4.4 \times 10^{-8}$) and friendship satisfaction (rg = –0.35, $P_{Bonferroni} = 2.85 \times 10^{-12}$) (**Supplementary Fig. 16**).

We processed those summary statistics using SBayesRC [44] (gctb v.2.5.2) using the LD matrix UKeur_HM3. We then extracted the UKeur_HM3 SNPs from the WGS data and imputed the missing ones using gctb's guidelines. We then computed the PGS using PLINK v.2 [45] (v.20240818) score option. All personal PGS were corrected for ancestry using the first 10 PCs before being normalized using the neurotypical people from European ancestry.

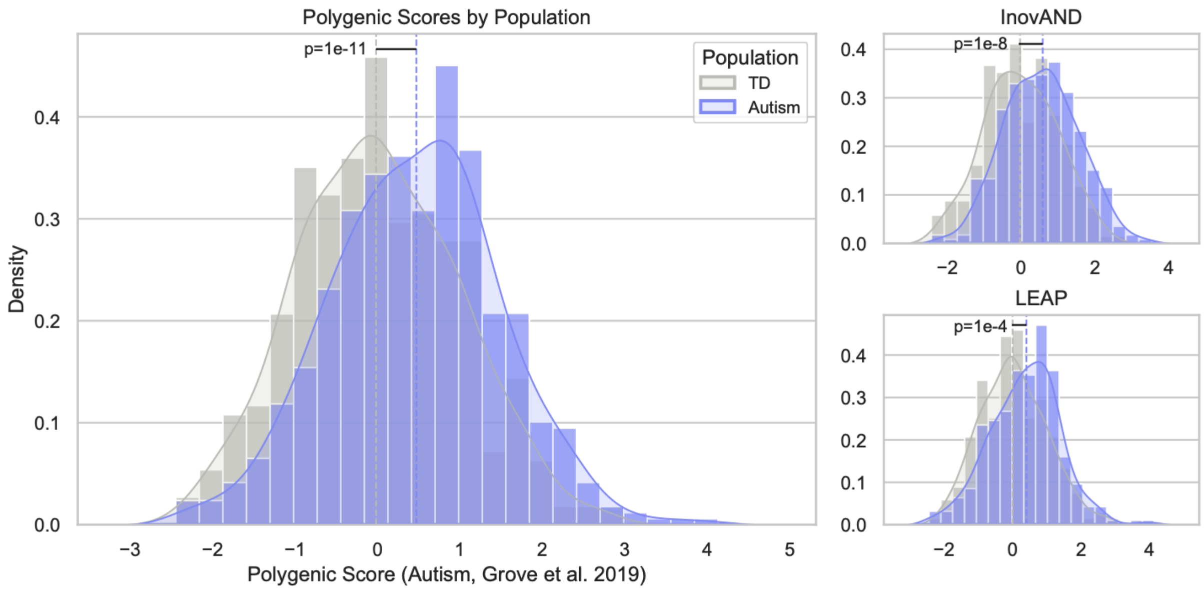

Supplementary Figure 14: Polygenic scores in the LEAP-InovAND datasets. PGS of autism (Grove et al. 2019), with LEAP and InovAND separated

##### *2.7 Statistical analysis for genetic associations*

For genetic data, associations between the presence of genetic variants and categorical variables were assessed using Fisher's exact test, with or without false discovery rate (FDR, one-sided independent t-tests adjusted for multiple comparisons using Benjamini–Hochberg correction) correction depending on the dependence structure of the data. Odds ratios (ORs) with 95% confidence intervals (CI95%) were calculated using the neurotypical (NT) group or, when appropriate, the combined NT/sibling group as the reference category.

##### *2.8 Replication in the SPARK cohort*

To confirm the robustness of our findings, we replicated the analyses in the SPARK cohort, a large-scale study of people with autism. SPARK includes a much larger number of participants and provides access to detailed clinical and genetic information. These features make it possible to validate and extend our results in a broader, well-characterized population.

Genetic analyses were conducted using datasets available for download (iWES v.3). Rare variant analyses were performed on whole-exome sequencing (WES) data and were restricted to SNV/indels. For common variants analyses, SNPs were detected using SNP-arrays or the Twist Comprehensive Exome panel, followed by separate imputation procedures using the HRC v1.1 panel and subsequent dataset merging.

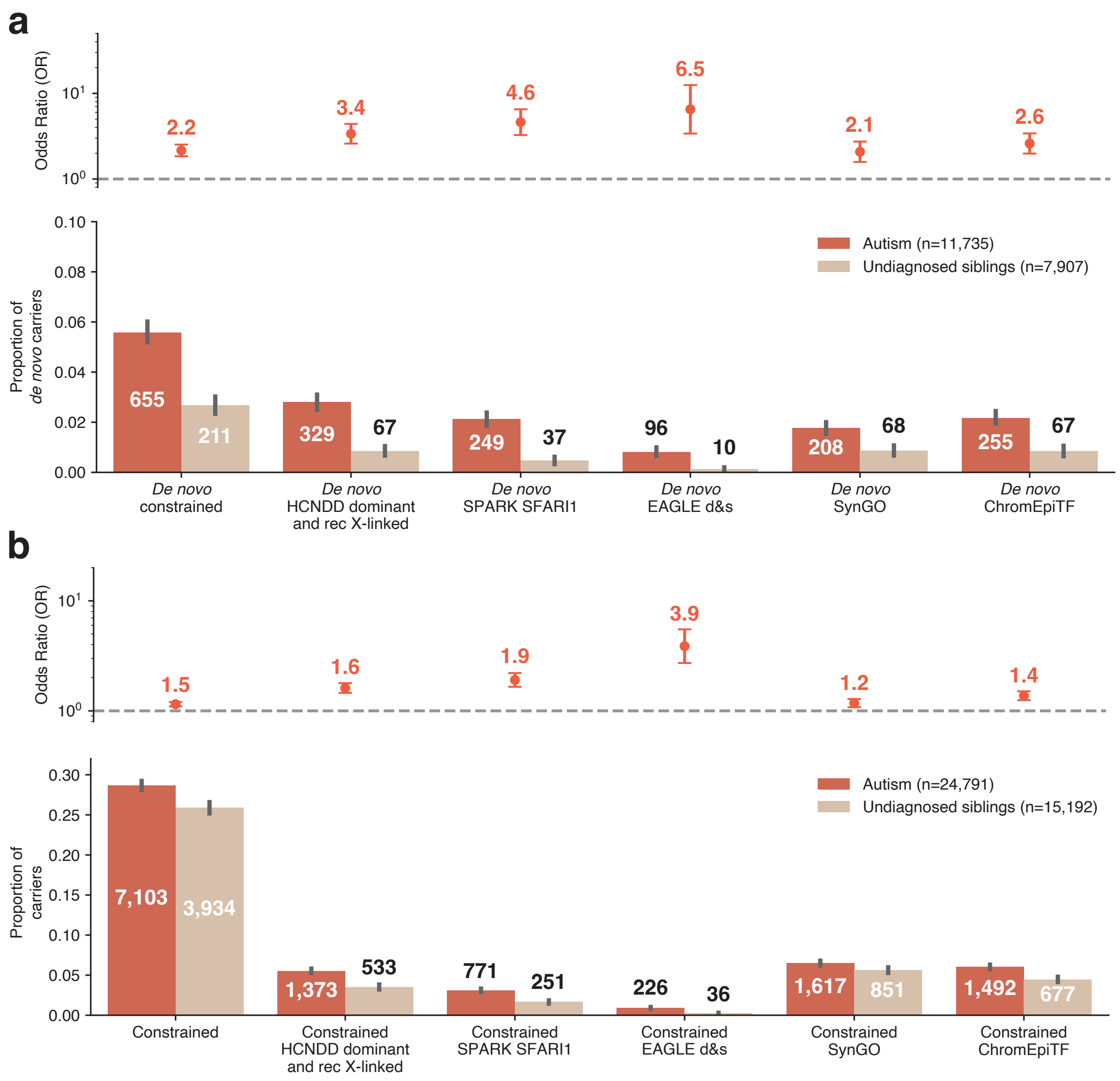

Supplementary Figure 15: Rare variant study in the SPARK replication cohort. **a**. Genetic study of *de novo* variants (loss of function and deleterious missenses). The bottom panel represents the proportion of *de novo* variant carriers according to the different gene lists used. The figures represent the number of carriers per population and per gene list. The top panel represents the OR (with CI95%) based on the proportions of the bottom panel. **b**. Genetic study of hiURV variants in constraint genes. OR (with CI95%) are calculated using undiagnosed siblings as a reference.

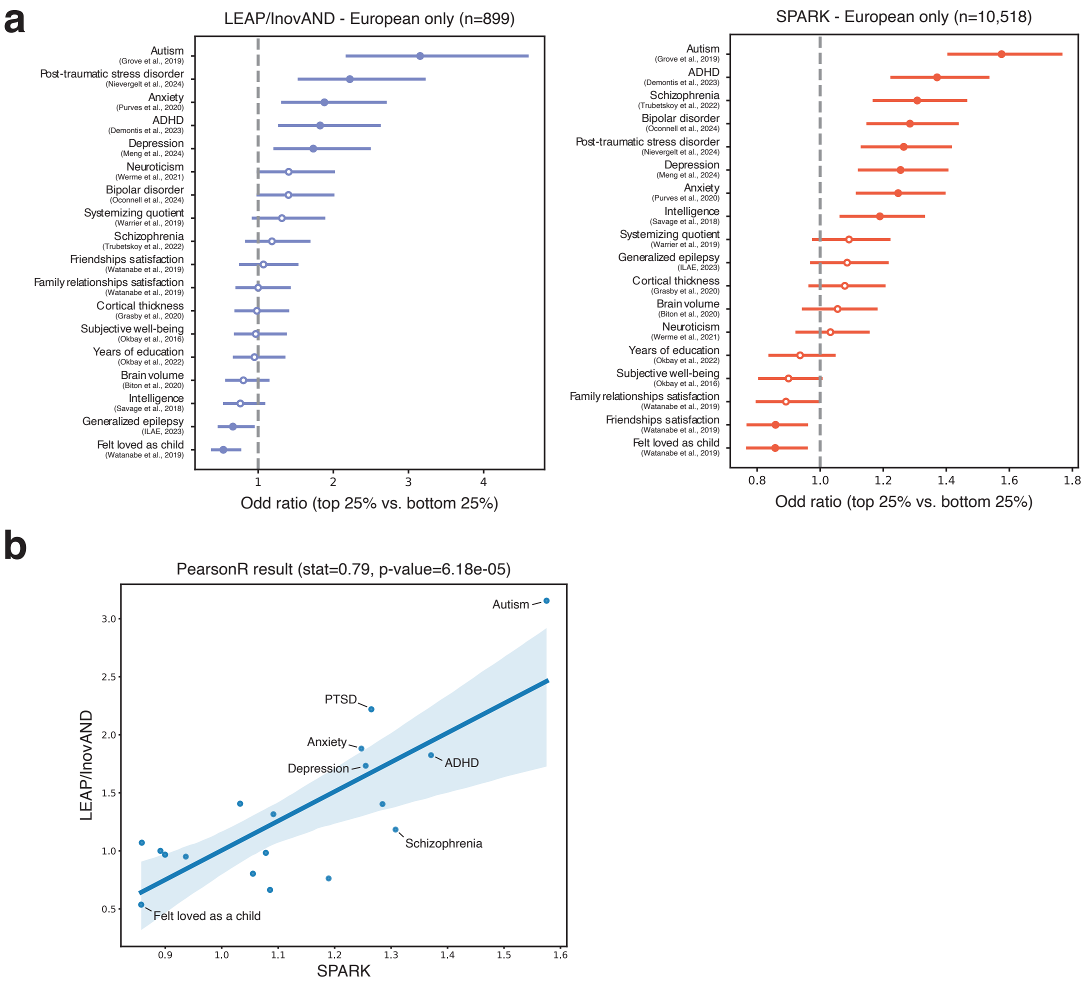

Supplementary Figure 16: Polygenic scores in the LEAP-InovAND and SPARK datasets. **a - left panel** OR quantifying the enrichment of autistic people in Q1 (top 25%) versus Q4 (bottom 25%) of the PGS distribution for each trait, compared with non-autistic people of European ancestry from the LEAP-InovAND cohort. **a - right panel** Odds ratios (OR) quantifying the enrichment of autistic people in the top quartile (Q4) versus the bottom quartile (Q1) of the polygenic score (PGS) distribution for each trait, relative to their non-autistic siblings within European participants of the SPARK cohort **b**. Scatter plot showing the Pearson correlation of OR between the SPARK and LEAP-InovAND cohorts.

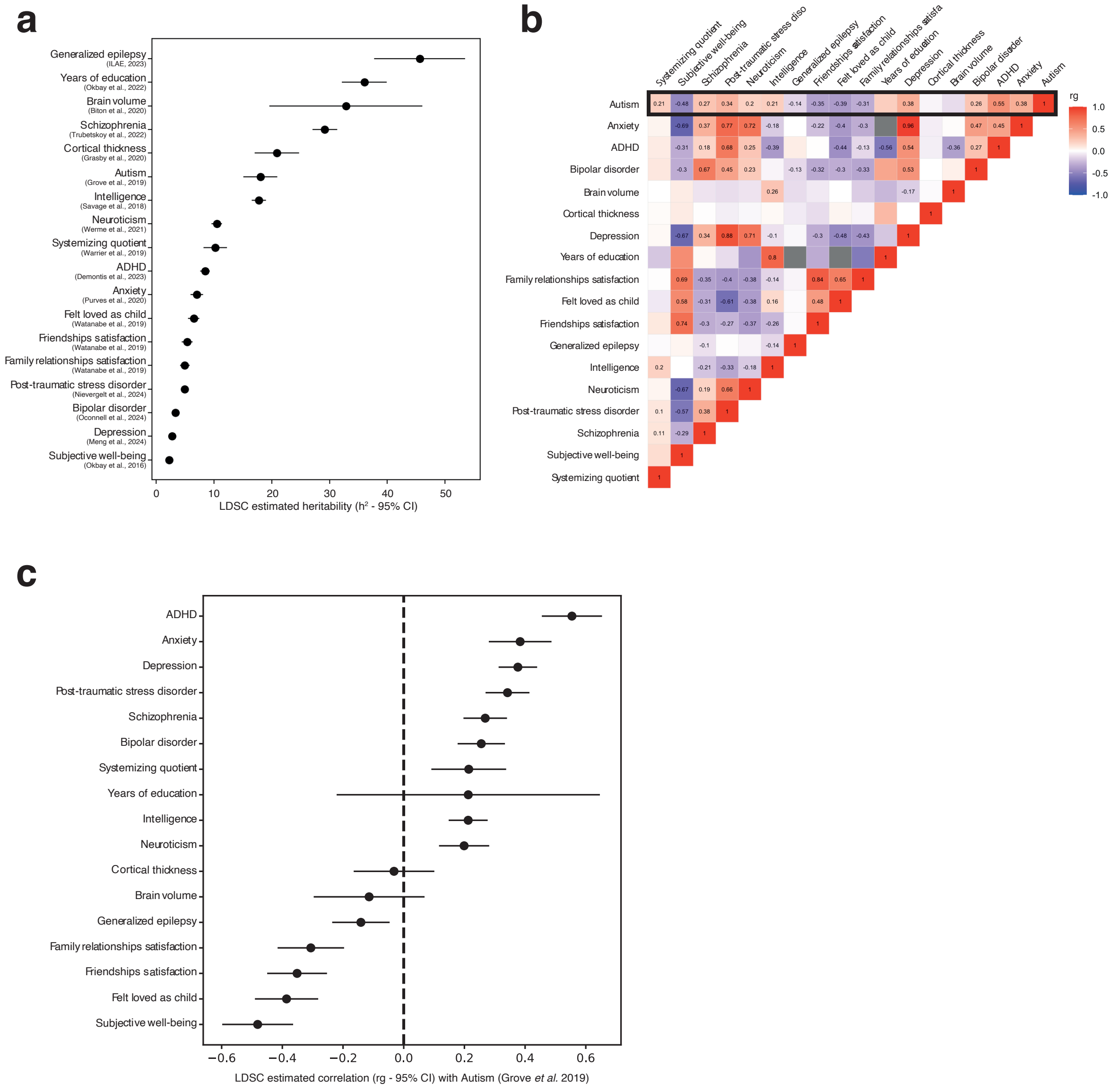

Supplementary Figure 17: Genome-wide estimates of heritability and genetic correlations. **a**. SNP-based heritability ($h^2$) estimates with 95% confidence intervals (CIs), calculated using LDSC v1.0.1 [46]. **b**. Pairwise genetic correlations (rg) between traits estimated with LDSC. Significant correlations after Bonferroni correction for multiple testing are indicated. Correlation values range from -1 (negative correlation, blue) to +1 (positive correlation, red). **c**. Genetic correlations of the analyzed traits with the autism GWAS, shown with corresponding 95% CI.

#### Section 4. MRI acquisition and pre-processing

*MRI Acquisition*

*LEAP cohort*: Magnetic resonance imaging (MRI) data were acquired across five sites participating in the LEAP consortium: Cambridge University, King's College London (KCL), Central Institute of Mental Health Mannheim, Radboud University Nijmegen Medical Centre and University Medical Center Utrecht, using a standardized MRI protocol. The structural scan was acquired using a magnetization prepared rapid gradient-echo (MPRAGE) sequence according to the ADNI-2/GO-protocol (http://adni.loni.usc.edu). The resting-state fMRI scan was acquired using a multi-echo planar imaging sequence [47]. For detailed acquisition parameters for each site in the LEAP cohort (see **Supplementary Table 5**). High-resolution structural T1-weighted volumetric images were acquired with full head coverage, at 1.2-mm thickness with 1.2×1.2-mm in-plane resolution (see **Supplementary Table 4**).

InovAND cohort: MRI data for the InovAND cohort were collected at a single site between 2010 and 2024, using three Philips scanners operating at two field strengths (1.5T & 3T, **Supplementary Table 6**). Regarding the sMRI acquisition, two types of T1-weighted sequences were used: Fast Field Echo (FFE) sequence on a 1.5 T Ingenia Philips machine: TR = 25 ms, TE = 5.6 ms, FOV = 240x240 mm, a 1-mm isotropic voxel resolution, and 170 slices. This sequence was adapted for the 3 T Ingenia machine. A 3D Turbo Field Echo (TFE) sequences: These were implemented on all machines with shorter TR and TE values (TR = [7-8] ms and TE = [3-4] ms), which allowed a wider FOV = 260x260 mm, 175 slices while keeping a 1mm isotropic resolution. They were also implemented on the 3T machine, with a finer voxel resolution of 0.5x0.5x0.75 mm (using Sense and Compressed Sensing (CS) to fasten the acquisition). An updated TFE sequence version has been recently implemented with the following parameters: TR = 6.8 ms, TE = 3 ms, FOV = 256x256 mm, 200 slices, and 0.9 mm isotropic voxel resolution (see **Supplementary Table 7**).

Supplementary Table 4: LEAP structural MRI acquisition parameters across sites. Note. TR: repetition time; TE: echo time; FA: flip angle; FOV: field of view.

| LEAP - sMRI Acquisition Parameters by Site | | | | | | | | |
|---|---|---|---|---|---|---|---|---|
| Site | Scanner Model | Field Strength | Acquisition Sequence | TR / TE / FA / FOV | Slice Thickness | Number of Slices | Voxel Size | Coverage |
| Cambridge | Siemens Magnetom Verio | 3T | Tfl3d1_ns | 2.3/2.95/9/270 | 1.2 | 176 | 1.1 x 1.1 x 1.2 | 256*256 |
| KCL | GE Discovery mr750 | 3T | SAG ADNI GO ACC SPGR | 7.31/3.02/11/270 | 1.2 | 196 | 1.1 x 1.1 x 1.2 | 256*256 |
| Mannheim | Siemens Magnetom TIM Trio | 3T | MPRAGE ADNI | 2.3/2.93/9/270 | 1.2 | 176 | 1.1 x 1.1 x 1.2 | 256*256 |
| Nijmegen | Siemens Magnetom Skyra | 3T | Tfl3d1_16ns | 2.3/2.93/9/270 | 1.2 | 176 | 1.1 x 1.1 x 1.2 | 256*256 |
| Rome | GE Signa HDxt | 3T | SAG ADNI GO ACC SPGR | 5.96/1.76/11/270 | 1.2 | 172 | 1.1 x 1.1 x 1.2 | 256*256 |
| Utrecht | Philips Achieva/Ingenia CX | 3T | ADNI GO 2 | 6.76/3.1/9/270 | 1.2 | 170 | 1.1 x 1.1 x 1.2 | 256*256 |

Supplementary Table 5: LEAP rsfMRI acquisition parameters by site and EPI BOLD multi-echo imaging sequence. Note. TR: repetition time; TE: echo time; FA: flip angle; FOV: field of view.

| LEAP - fMRI Acquisition Parameters by Site | | | | | | | |
|---|---|---|---|---|---|---|---|
| Site | Scanner Model | Field Strength | Instruction | TR / TE / TI / FA / Slices | N_vol | N_sclices | Vox_size |
| Cambridge | Siemens Magnetom Verio | 3T | Fixation | 2300/ 12/29/46/80 | 266 | 33 | 3.8 x 3.8 x 3.8 |
| KCL | GE Discovery mr750 | 3T | Fixation | 2300/ NA/31/48/90 | 215 | 33 | 3.8 x 3.8 x 3.9 |
| Mannheim | Siemens Magnetom TIM Trio | 3T | Fixation | 2300/ 12/29/46/80 | 215 | 33 | 3.8 x 3.8 x 3.10 |
| Nijmegen | Siemens Magnetom Skyra | 3T | Fixation | 2300/ 12/NA/NA/80 | 266 | 33 | 3.8 x 3.8 x 3.11 |
| Rome | GE Signa HDxt | 3T | Fixation | | NA | NA | |
| Utrecht | Philips Achieva/Ingenia CX | 3T | Fixation | 2300/ 13/31/49/80 | 200 | 33 | 3.75 x 3.75 x 3.75 |

Supplementary Table 6: InovAND structural MRI acquisition parameters across sites. Note. TR: repetition time; TE: echo time; FA: flip angle; FOV: field of view.

| InovAND - sMRI Acquisition Parameters by Site | | | | | | | | |
|---|---|---|---|---|---|---|---|---|
| Site | Scanner Model | Field Strength | Acquisition Sequence | TR / TE / FA / FOV | Slice Thickness | Number of Slices | Voxel Size | Coverage |
| | | | | | NA | NA | | NA |
| RDB 1 | Ingenia Philips machine | 1.5T | Fast Field Echo (FFE) | 2.5/5.6/NA/240 | 1.2 | 170 | 1.0 x 1.0 x 1.0 | NA |
| RDB 2 | Ingenia Philips machine | 1.5T | 3D Turbo Field Echo | 7.8/3.4/NA/260 | 1.2 | 170 | 1.0 x 1.0 x 1.1 | NA |
| RDB 3 | Ingenia Philips machine | 3T | 3D Turbo Field Echo | 7.8/3.4/NA/261 | 1.2 | 175 | 0.5 x 0.5 x 0.75 | NA |

Supplementary Table 7: InovAND rsfMRI acquisition parameters by site and EPI BOLD multi-echo imaging sequence. Note. TR: repetition time; TE: echo time; FA: flip angle; FOV: field of view.

| InovAND - fMRI Acquisition Parameters by Site | | | | | | | |
|---|---|---|---|---|---|---|---|
| Site | Scanner Model | Field Strength | Instruction | TR / TE / TI / FA / Slices | N_vol | N_sclices | Vox_size |
| RDB 1 | Philips Intera | 1.5T | Fixation | 2700/45/NA/90/4 | 255 | 32 | 3.6 x 3.6 x 4 |
| RDB 2 | Philips Ingenia | 1.5T | Fixation | 2700/45/NA/90/4 | 255 | 33 | 3.6 x 3.6 x 5 |
| RDB 3 | Philips Ingenia | 3T | Fixation | 2700/45/NA/90/5 | 255 | 33 | 3.6 x 3.6 x 4 |

*sMRI Preprocessing*

All T1-weighted structural brain MRI scans were processed using FreeSurfer (version 6.0.0). Quality control of the T1-weighted brain MR images and the segmentation of the cortical and subcortical regions was performed by two different raters (AA and ML) for InovAND dataset and by three independent raters, blind to group membership for LEAP dataset, more info in [48]. Images were segmented using the Desikan-Killiany atlas 33 into 68 cortical and 14 subcortical regions + 2 lateral ventricles. Cortical thickness (CT) and surface area (SA) measures were extracted for the 68 cortical regions (34 per hemisphere). Volumes were calculated for the 16 subcortical regions. We also extracted global metrics from FreeSurfer: mean CT, total SA, intracranial volume (ICV), total gray volume (GV), and cerebrospinal fluid (CSF) volume.

*rsfMRI Preprocessing*

rsfMRI and structural scans were obtained using 3T and 1.5T across 7 sites (See Supplementary Section 3). Fmriprep (v.23.0.0) was applied to realign and slice-time correct the fMRI volumes. The resulting fMRI volumes were co-registered to their corresponding T1 volumes using FLIRT. Finally, the fMRI time series underwent confound regression using the eXtensible Connectivity Pipeline (XCP (v.0.8.0); https://github.com/PennLINC/xcp_d) with the 36p+Despike model. In brief, bandpass filtering between 0.01 and 0.08 Hz was applied, followed by despiking with AFNI and regression of 36 parameters from the time series. Regressors included six motion attributes derived during realignment, a global signal and two physiological parameters, as well as their derivatives, squares of derivatives and quadratic terms. Mean framewise displacement was calculated, from Power *et al.*[49], for each participant and used to account for any residual effect of head motions in subsequent analysis. We removed high motion volumes (> 0.3 mm framewise displacement due to head movement) and the volume immediately preceding and following these volumes. Following motion censoring, we selected participants with at least 3 min of quality data.

To investigate autism-related FC alterations across the brain, we applied the well-validated Schaefer parcellation to divide the cortex of each participant into 100 functional regions of interest (ROI) [50] combined with 15 subcortical ROIs from the California Institute of Technology probabilistic subcortical atlas [51]. All functional MRI data were aligned to the MNI152NLin2009cAsym template space via each subject's T1-weighted anatomical image using nonlinear registration. We extracted the mean time series from each ROI and computed the Pearson correlation between every pair of time series to create the similarity matrix. Then we applied the Fisher transformation to each similarity matrix. To reduce the influence of outliers and improve comparability across subjects, we normalized each FC matrix using a robust scaling procedure. Specifically, we subtracted the median and divided by the interquartile range (IQR). This transformation ensures that the dynamic range of connectivity values is standardized across people while preserving personal differences in FC patterns. We used ComBat [52] to regress the scan-site-specific effects from the FC data.

To contextualize findings within known functional networks, we assigned cortical ROI to the Yeo 7-network parcellation. This allowed us to quantify connectivity alterations within and between canonical resting-state networks. Based on the method from Ilioska et al. [53], for each network pair, we computed (1) the raw number of ROI-to-ROI connections that significantly differed between diagnostic groups based on independent two-sample t-tests ($N_{raw}$), and (2) the proportion of altered connections relative to the number of possible edges ($N_{norm}$). While $N_{raw}$ highlights networks with large absolute contributions to group differences, $N_{norm}$ adjusts for differences in network size and density, revealing disproportionately affected systems.

*Neuroimaging Statistical Analyses*

*Structural MRI*

For each neuroimaging feature of interest, including subcortical volumes, cortical thickness, and cortical surface area measures derived from the Desikan–Killiany atlas, we fitted a linear regression model with age, sex, age squared, MRI acquisition site, and estimated total intracranial volume (eTIV) as covariates. Specifically, for each feature $i$, the following model was estimated:

$$Y = \beta_0 + \beta_1 Sex + \beta_2 Age + \beta_3 Age^2 + \beta_4 Site + \beta_5 TIV + \varepsilon,$$

where $\varepsilon$ represents the residual error term. Residuals ($\varepsilon$) were extracted for everyone. To ensure comparability across participants, residuals were standardized by computing z-scores relative to the mean and standard deviation of the NT group. Specifically, for each feature, z-scores were calculated as:

$$z = \frac{\varepsilon - \mu_{TD}}{\sigma_{TD}},$$

where $\mu_{TD}$ and $\sigma_{TD}$ are the mean and standard deviation of the residuals in the NT population, respectively. At each ROI, we also used a Welch's t-test to assess between group differences at a false discovery rate (FDR) corrected p value ($p_{adj}$) $<$ 0.05

Supplementary Table 8: Cortical Thickness- Autism vs NT. ROI: Region of Interest (cortical regions from the Desikan-Killiany atlas) t: t-statistic from group comparison (autism vs NT); p: Uncorrected p-value; q: Bonferroni-corrected p-value; d: Cohen's d (effect size).

| | Left Hemisphere | | | | Right Hemisphere | | | |
|---|---|---|---|---|---|---|---|---|
| **ROI** | t | p | q | d | t | p | q | d |
| Bankssts | 3.246 | 0.001224 | 0.083204 | 0.199 | 2.777 | 0.005626 | 0.382583 | 0.172 |
| Caudalanteriorcingulate | 1.411 | 0.158538 | 1 | 0.086 | −0.238 | 0.811865 | 1 | −0.015 |
| Caudalmiddlefrontal | 2.177 | 0.029814 | 1 | 0.135 | 1.534 | 0.125337 | 1 | 0.094 |
| Cuneus | 2.526 | 0.011744 | 0.798589 | 0.159 | 2.837 | 0.004674 | 0.317812 | 0.173 |
| Entorhinal | 0.295 | 0.768352 | 1 | 0.018 | −0.381 | 0.703072 | 1 | −0.023 |
| Frontalpole | 3.051 | 0.002356 | 0.160224 | 0.183 | 1.018 | 0.308998 | 1 | 0.063 |
| Fusiform | 2.156 | 0.031433 | 1 | 0.138 | 3.234 | 0.001283 | 0.087262 | 0.210 |
| Inferiorparietal | 2.687 | 0.00737 | 0.501189 | 0.165 | 3.542 | 0.000422 | 0.028706 | 0.220 |
| Inferiortemporal | 3.386 | 0.00075 | 0.051007 | 0.215 | 1.901 | 0.057659 | 1 | 0.119 |
| Insula | 1.919 | 0.055338 | 1 | 0.122 | 0.603 | 0.546642 | 1 | 0.037 |
| Isthmuscingulate | 2.351 | 0.019001 | 1 | 0.150 | 0.775 | 0.438376 | 1 | 0.049 |
| Lateraloccipital | 2.783 | 0.005534 | 0.376296 | 0.176 | 3.416 | 0.000672 | 0.04568 | 0.216 |
| Lateralorbitofrontal | 1.780 | 0.075518 | 1 | 0.111 | 1.484 | 0.138341 | 1 | 0.091 |
| Lingual | 2.778 | 0.005619 | 0.382072 | 0.174 | 2.301 | 0.021694 | 1 | 0.146 |
| Medialorbitofrontal | 1.921 | 0.055161 | 1 | 0.120 | 1.788 | 0.074194 | 1 | 0.109 |
| Middletemporal | 2.838 | 0.004662 | 0.316994 | 0.175 | 2.227 | 0.02624 | 1 | 0.138 |
| Paracentral | 1.027 | 0.304791 | 1 | 0.062 | 1.371 | 0.170701 | 1 | 0.085 |
| Parahippocampal | −0.521 | 0.602392 | 1 | −0.033 | −0.905 | 0.365705 | 1 | −0.058 |
| Parsopercularis | 1.908 | 0.05683 | 1 | 0.118 | 2.387 | 0.017249 | 1 | 0.150 |
| Parsorbitalis | 2.197 | 0.028375 | 1 | 0.138 | 1.855 | 0.063988 | 1 | 0.117 |
| Parstriangularis | 1.271 | 0.204246 | 1 | 0.078 | 1.380 | 0.167925 | 1 | 0.087 |
| Pericalcarine | 1.948 | 0.05179 | 1 | 0.122 | 1.605 | 0.108973 | 1 | 0.099 |
| Postcentral | 1.426 | 0.154374 | 1 | 0.087 | 2.061 | 0.039632 | 1 | 0.125 |
| Posteriorcingulate | 2.793 | 0.005368 | 0.365051 | 0.176 | 1.805 | 0.071539 | 1 | 0.114 |
| Precentral | 1.809 | 0.070876 | 1 | 0.114 | −0.079 | 0.937349 | 1 | −0.005 |
| Precuneus | 2.637 | 0.008533 | 0.580258 | 0.165 | 3.186 | 0.001505 | 0.102373 | 0.197 |
| Rostralanteriorcingulate | 2.356 | 0.018775 | 1 | 0.150 | 1.535 | 0.125131 | 1 | 0.096 |
| Rostralmiddlefrontal | 3.673 | 0.00025 | 0.017496 | 0.228 | 3.146 | 0.001723 | 0.117185 | 0.197 |

| | | | | | | | | |
|---|---|---|---|---|---|---|---|---|
| | | 7 | | | | | | |
| Superiorfrontal | 2.450 | 0.014518 | 0.987222 | 0.150 | 2.079 | 0.037988 | 1 | 0.128 |
| Superiorparietal | 2.059 | 0.039845 | 1 | 0.125 | 2.502 | 0.012567 | 0.854578 | 0.151 |
| Superiortemporal | 5.357 | 1.122e-07 | 7.628e-06 | 0.328 | 3.609 | 0.000327 | 0.022255 | 0.222 |
| Supramarginal | 2.589 | 0.0098 | 0.666388 | 0.155 | 1.860 | 0.063325 | 1 | 0.115 |
| Temporalpole | 0.014 | 0.989035 | 1 | 0.001 | −0.536 | 0.592071 | 1 | −0.033 |
| Transversetemporal | 2.794 | 0.005338 | 0.363005 | 0.173 | 1.720 | 0.085805 | 1 | 0.107 |

Bonferroni correction: Highlighted cells indicate significant results ($q < 0.05$).

Supplementary Table 9: Surface Area- Autism vs NT. ROI: Region of Interest (cortical regions from the Desikan-Killiany atlas) t: t-statistic from group comparison (autism vs NT); p: Uncorrected p-value; q: Bonferroni-corrected p-value; d: Cohen's d (effect size).

| | Left Hemisphere | | | | Right Hemisphere | | | |
|---|---|---|---|---|---|---|---|---|
| **ROI** | t | p | q | d | t | p | q | d |
| Bankssts | 0.781 | 0.434804 | 1 | 0.049 | 1.161 | 0.245975 | 1 | 0.072 |
| Caudalanteriorcingulate | 0.331 | 0.74067 | 1 | 0.021 | −1.059 | 0.289856 | 1 | −0.069 |
| Caudalmiddlefrontal | 0.466 | 0.641618 | 1 | 0.030 | 0.907 | 0.364855 | 1 | 0.058 |
| Cuneus | 0.264 | 0.791539 | 1 | 0.016 | 0.269 | 0.788276 | 1 | 0.017 |
| Entorhinal | 0.372 | 0.709683 | 1 | 0.023 | 0.231 | 0.817327 | 1 | 0.014 |
| Frontalpole | −0.399 | 0.690073 | 1 | −0.026 | 0.563 | 0.573947 | 1 | 0.036 |
| Fusiform | 1.712 | 0.087443 | 1 | 0.110 | 1.392 | 0.164259 | 1 | 0.092 |
| Inferiorparietal | 0.376 | 0.707393 | 1 | 0.024 | 0.963 | 0.335797 | 1 | 0.061 |
| Inferiortemporal | 0.661 | 0.50906 | 1 | 0.042 | 1.177 | 0.239779 | 1 | 0.076 |
| Insula | −0.108 | 0.914245 | 1 | −0.007 | 0.389 | 0.697212 | 1 | 0.025 |
| Isthmuscingulate | 1.128 | 0.259618 | 1 | 0.069 | 0.710 | 0.477954 | 1 | 0.045 |
| Lateraloccipital | 1.192 | 0.233783 | 1 | 0.075 | 1.842 | 0.065886 | 1 | 0.118 |
| Lateralorbitofrontal | −0.614 | 0.539174 | 1 | −0.039 | 0.648 | 0.516987 | 1 | 0.041 |
| Lingual | 0.841 | 0.400447 | 1 | 0.054 | 1.878 | 0.060854 | 1 | 0.119 |
| Medialorbitofrontal | 0.763 | 0.445479 | 1 | 0.051 | 0.917 | 0.359572 | 1 | 0.058 |
| Middletemporal | 1.308 | 0.191195 | 1 | 0.081 | 1.490 | 0.136658 | 1 | 0.093 |
| Paracentral | 0.516 | 0.605759 | 1 | 0.032 | 0.655 | 0.512665 | 1 | 0.040 |
| Parahippocampal | 2.881 | 0.004084 | 0.277732 | 0.181 | 1.918 | 0.055512 | 1 | 0.125 |
| Parsopercularis | −0.049 | 0.960694 | 1 | −0.003 | −0.801 | 0.423491 | 1 | −0.050 |
| Parsorbitalis | −1.029 | 0.303929 | 1 | −0.063 | −0.253 | 0.800283 | 1 | −0.016 |
| Parstriangularis | −1.157 | 0.247465 | 1 | −0.073 | −2.335 | 0.019823 | 1 | −0.146 |
| Pericalcarine | 0.372 | 0.709779 | 1 | 0.023 | 1.429 | 0.153429 | 1 | 0.087 |
| Postcentral | 1.652 | 0.099003 | 1 | 0.104 | 1.545 | 0.12277 | 1 | 0.094 |
| Posteriorcingulate | −0.154 | 0.877475 | 1 | −0.010 | 1.067 | 0.286443 | 1 | 0.066 |
| Precentral | −0.174 | 0.861696 | 1 | −0.011 | 0.632 | 0.527345 | 1 | 0.039 |
| Precuneus | 1.616 | 0.106529 | 1 | 0.100 | 1.813 | 0.070301 | 1 | 0.113 |
| Rostralanteriorcingulate | 0.997 | 0.319264 | 1 | 0.063 | 0.610 | 0.541887 | 1 | 0.040 |
| Rostralmiddlefrontal | 0.068 | 0.945469 | 1 | 0.004 | 0.442 | 0.658338 | 1 | 0.028 |
| Superiorfrontal | 0.565 | 0.572392 | 1 | 0.036 | 0.526 | 0.598984 | 1 | 0.033 |
| Superiorparietal | 1.760 | 0.078757 | 1 | 0.110 | 0.509 | 0.61089 | 1 | 0.031 |
| Superiortemporal | 1.157 | 0.247648 | 1 | 0.073 | 1.283 | 0.199969 | 1 | 0.079 |
| Supramarginal | 1.587 | 0.112941 | 1 | 0.102 | 1.363 | 0.173254 | 1 | 0.084 |
| Temporalpole | 0.250 | 0.802376 | 1 | 0.015 | 2.136 | 0.033008 | 1 | 0.138 |
| Transversetemporal | −0.192 | 0.848119 | 1 | −0.012 | 0.152 | 0.879586 | 1 | 0.010 |

Bonferroni correction: Highlighted cells indicate significant results (q < 0.05).

Supplementary Table 10: Subcortical Volumes- Autism vs NT. ROI: Region of Interest (cortical regions from the Desikan-Killiany atlas) t: t-statistic from group comparison (autism vs NT); p: Uncorrected p-value; q: Bonferroni-corrected p-value; d: Cohen's d (effect size).

| | Left Hemisphere | | | | Right Hemisphere | | | |
|---|---|---|---|---|---|---|---|---|
| ROI | t | p | q | d | t | p | q | d |
| Amygdala | 2.120 | 0.034297 | 0.960306 | 0.128 | 2.238 | 0.025533 | 0.714921 | 0.142 |
| Caudate | 1.090 | 0.275987 | 1 | 0.067 | 1.348 | 0.178082 | 1 | 0.083 |
| Cerebellum-Cortex | 0.747 | 0.455428 | 1 | 0.047 | 1.620 | 0.105655 | 1 | 0.103 |
| Cerebellum-White-Matter | 0.498 | 0.618824 | 1 | 0.031 | −0.017 | 0.986837 | 1 | −0.001 |
| Hippocampus | −1.114 | 0.265676 | 1 | −0.072 | −0.138 | 0.890628 | 1 | −0.009 |
| Lateral-Ventricle | 1.855 | 0.064026 | 1 | 0.117 | 3.262 | 0.001154 | 0.032306 | 0.196 |
| Pallidum | −0.940 | 0.347374 | 1 | −0.058 | −0.403 | 0.687007 | 1 | −0.025 |
| Putamen | 0.263 | 0.792757 | 1 | 0.016 | 0.100 | 0.920102 | 1 | 0.006 |
| Thalamus-Proper | −0.391 | 0.695547 | 1 | −0.024 | −0.077 | 0.93902 | 1 | −0.005 |
| Ventraldc | 1.418 | 0.156704 | 1 | 0.092 | 0.939 | 0.348203 | 1 | 0.061 |

Bonferroni correction: Highlighted cells indicate significant results (q < 0.05).

#### *Resting state fMRI*

To assess connectivity differences between clusters, we performed two-sided Welch's t-tests (unequal variance) for each functional connection (*i.e.,* each edge in the connectivity matrix). To ensure robustness, only connections with a false discovery rate (FDR)-corrected p-value< 0.05 were retained for further analysis. Each significant connection was annotated with its corresponding source and target regions based on atlas-defined labels. For each contrast, we quantified the number of significantly hyperconnected (t>0) and hypoconnected (t< 0) edges within and between network pairs.

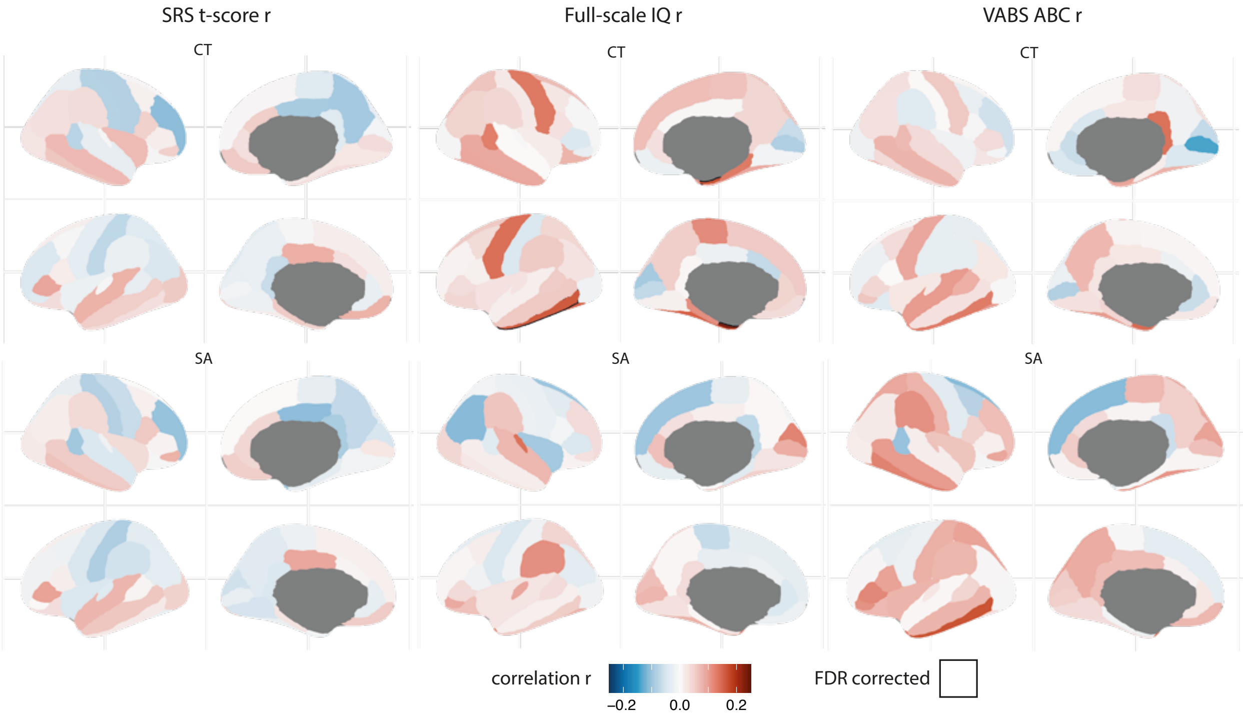

Supplementary Figure 18: **Associations between cortical thickness/surface area and clinical features in people with autism**. Cortical thickness/surface area was correlated with four clinical measures using Pearson correlation across the autism cohort. Brain maps display vertex-wise correlation coefficients (r) for: social responsiveness scale total score (SRS-2 t-score), repetitive behavior scale-revised total score (RBS-R), full-scale IQ (FSIQ), and (VABS)-II. CT: Cortical Thickness; SA: Surface Area.

#### *Removal of site-specific artifacts*

**Supplementary Fig. 19** shows the edges that show significant effects of scanning site before and after ComBat harmonization, indicating that ComBat successfully removed effects of scanning site. To further test the effects of our harmonization procedure, we trained a one vs. one, multilabel, support vector machine (SVM) linear classifier to classify data by scanning site. We trained the classifier on 70% of the functional connectivity data and tested it on the remaining 30% of the data. **Supplementary Fig. 19b** shows the area under the receiver operating characteristic curve (ROC AUC), across sites before and after ComBat, again demonstrating that ComBat successfully removed the effects of scanning site. ROC AUC scores for one vs. one SVM linear classification of scanning sites before and after ComBat.

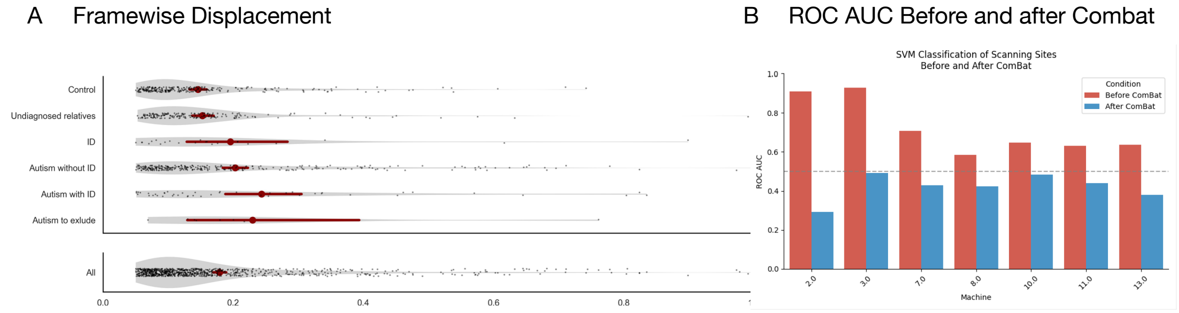

Supplementary Figure 19: Framewise Displacement and Combat ROC **A**. Framewise Displacement, calculated from Power et al. across all phenotypes **B**. Area under the receiver operating characteristic curve (AUC ROC) scores across scanning sites, before and after ComBat.

#### Section 5. EEG acquisition and Preprocessing

Five sites acquired EEG data at baseline, following international standards using three different systems.

EEG acquisition: LEAP: a complete description of EU-AIMS LEAP EEG protocol is described elsewhere [54]. Briefly, 4 minutes of resting state EEG were recorded per participant (2 min with eyes open and 2 min with eyes closed) with 30s blocks alternatively. All sites used 10–20 layout caps, with 60–70 electrodes. InovAND: all subjects were recorded in resting-state with eyes open and closed for periods of 3 to 5 minutes using a 129-GSN hydrocel Geodesic montage. Sampling was at 1,000 Hz, and the reference electrode was number 55 in the Cz position.

EEG preprocessing: Signal pre-processing and analysis were performed entirely in python using MNE-python [55]. EEG signals were resampled to 1000Hz and then high-pass filtered at 1 Hz to remove low-frequency drifts, and a notched filter was applied to suppress line noise (50 Hz and its harmonics). Then, we performed a robust referencing, utilizing an iterative method to optimize the EEG reference. During this step, noisy channels, identified prior to referencing, were excluded from the initial reference computations. These channels were later interpolated. IClabel was then performed, and bad ICs (independent components) were discarded.

Feature extraction: Power spectral density (PSD) was estimated using Welch's method with a Hanning window and 50% overlap. Absolute and relative power were calculated for five frequency bands: delta (1–4 Hz), theta (4–8 Hz), alpha (8–13 Hz), beta (13–30 Hz), and gamma (30–49 Hz). Power values were aggregated by brain region (Frontal, Central, Posterior) and laterality (Left, Right, Mid) based on channel groupings specific to each acquisition system. Alpha peak frequency identification: To maximize sensitivity to posterior alpha rhythms, spectral estimates were restricted to occipital electrodes (e.g., O1, O2, Oz, Poz & E72, E77, E76, E71, E62, E67). Power spectra were extracted from 1 to 50 Hz, with PSD values averaged across both epochs and the selected channels to obtain a robust subject-level spectrum. Alpha peak frequency was identified by finding the maximum amplitude within the 6–15 Hz frequency range for each participant. The peak was identified separately for eyes open and eyes closed conditions. Manual validation was performed for each participant to ensure accurate peak detection.

Statistical correction: To account for demographic and site effects, the following correction pipeline was applied. For each feature (power band × brain region), linear regression was performed to remove the effects of age, age-squared, and sex. This step was applied separately to each feature to account for feature-specific relationships with demographic variables. Site effect correction: ComBat [58] was applied to the regression residuals to remove site and cohort effects while preserving biological variability. ComBat uses an empirical Bayes framework to estimate and remove batch effects, with age, sex, age-squared, and cohort included as covariates in the ComBat model.

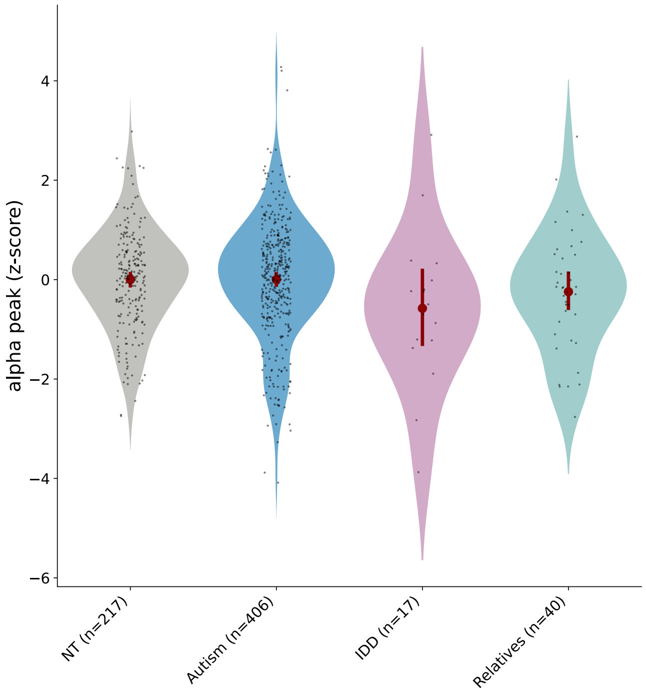

Supplementary Figure 20: EEG alpha peak [6-13] Hz across phenotypes.

### Section 6. Clustering analyses

Supplementary Table 11: Review of studies applying clustering methods in autism research. ABIDE: Autism Brain Imaging Data Exchange, ADHD: attention-deficit/hyperactivity disorder, AGRE: Autism Genetic Resource Exchange, AHBA: Allen Human Brain Atlas, CABIC: China Autism Brain Imaging Consortium, CARD: Cambridge Autism Research Database, CT: Cortical Thickness, EHR: Electronic Health Record, fMRI: functional MRI, GFMM: General Finite Mixture Model, HBN: Healthy Brain Network, HCL: hierarchical clustering, KMC: K-means clustering, LEAP: Longitudinal European Autism Project, LoF: Loss-of-Function variants (nonsense, frameshift and splice-site), MRI : Magnetic Resonance Imaging (of the brain), MSEL: Mullen Scales of Early Learning, NDA: National Institute of Mental Health Data Archive, OCD: obsessive–compulsive disorder, POND: Province of Ontario Neurodevelopmental Disorders Network, RMET: Reading the Mind in the Eyes Test, RRB: restricted and repetitive behaviors, SA: Surface Area, SPARK: Simons Foundation Powering Autism Research for Knowledge, NT: Neurotypical, VIQ: Verbal-Intellectual Quotient.

| **Year** | **Data input** | **Clustering method** | **cohort** | **Sample size** | **Age years** | **No. Clusters** | **Reference** |
|---|---|---|---|---|---|---|---|
| 2009 | ADI-R score | HCL, KMC | AGRE phenotype database | 1954 autism | 1-44 (mean=8) | 4 | H. Wu et al. [56] |
| 2012 | Evolution of social, communication, and repetitive behavior | maximum likelihood methods | local | 6975 autism | 2-14 | 6 | Fountain et al. [57] |
| 2016 | RMET | Agglomerative HCL | CARD | 694 autism, 249 NT | Mean=37 | 5 autism, 4 NT | Lombardo et al. [58] |
| 2018 | cognitive tasks + fMRI | Functional Random Forest | local | 47 autism and 58 NT | 9-13 | 3 autism, 4 NT | Feczko et al. [59] |
| 2020 | fMRI | KMC | POND | 407 (175 autism, 93 ADHD, 55 OCD, 84 NT) | 5-19 | 2 (clusters with neuroanatomical homogeneity did not correspond to diagnostic labels) | Choi et al. [60] |
| 2020 | CT (MRI) | Spectral clustering with a cosine similarity affinity matrix | LEAP | 316 autism 206 NT | 6-31 | 5 | Zahibi et al. [61] |
| 2020 | Exome Sequencing (segregating LoF) | Gene-set LoF burden | NDAR | 1704 pair of siblings (autism and NT) and 50 multi affected siblings | unknown | dyslipidemia-associated autism | Luo et al. [62] |
| 2021 | MRI (volume, CT, SA) and clinical (SCQ) | regression clustering | POND | 416: 192 autism, 109 ADHD , 50 OCD, 65 NT | 5-19 | 2 | Kushki et al. [63] |
| 2022 | Morphology (congenital anomalies, physical anomalies) | Arbitrary ranges: essential (score 0–3), equivocal (score 4–5) or complex (score ≥6) | ADOS | 325 autism | unknown | 3 | Chan et al. [64] |
| 2023 | fMRI | HCL | POND<br>HBN | 164 ADHD; 217 autism; 60 OCD; 110 NT<br>374 ADHD; 66 autism; 11 OCD; 100 NT | 5-19 | 2 and 6 | Vandewouw et al. [65] |
| 2023 | MRI (volume, CT, SA) | KMC | POND | 565 (262 autism, 171 ADHD, 132 NT) | 5-19 | 2 | Sadat et al. [66] |
| 2023 | VABS | KMC, *k*-nearest-neighbors | NDA | 1,098 autism | 0-6<br>13 | 3 | Mandelli et al. [67] |
| 2023 | fMRI + VIQ, social affect and RRB | cosine similarity matrix, HCL | ABIDE | 299 autism, 907 NT | | 4 | Buch et al. [68] |

| | | | | | | | |
|---|---|---|---|---|---|---|---|
| 2024 | MSEL and VABS | stability-based relative clustering validation analysis | NDA | 615 autism | 2-5.7 | 2 | Mandelli et al. [69] |
| 2025 | SA (MRI), brain transcriptome (AHBA) | KME | LEAP | 359 autism | 6-30 | 3 | Leyhausen et al. [70] |
| 2025 | SCQ, RBS-R, CBCL | GFMM | SPARK | 5,392 autism | 2-18 | 4 | Litman et al. [42] |
| 2025 | CT, SA, volume (MRI) | spectral clustering | POND HBN | 747 (312 autism, 220 ADHD, 70 OCD, 145 NT) 582 (60 autism, 445 ADHD, 19 OCD, 58 NT) | 5-19 | 2 | Sadat et al. [71] |
| 2025 | T1w/T2w-ratio (MRI) | Spectral clustering | POND | 310 (136 autism, 100 ADHD, 74 NT) | 5-19 | 3-4 | Norbom et al. [72] |
| 2025 | MRI | Spectral clustering | ABIDE, CABIC | 274 autism | <13 | 2 | Fan et al. [73] |

*Feature selection and optimal number of clusters.*

We initially selected a broad set of clinical measures, including three IQ subscales (full-scale IQ, verbal IQ, and non-verbal IQ), the Social Responsiveness Scale (SRS-2 t-score), the VABS (composite score and domain scores: communication, daily living skills, and socialization), the Repetitive Behavior Scale-Revised (RBS-R), and the Short Sensory Profile (SSP). Principal component analysis (PCA) was then performed using the FactoMineR package in R, after excluding people with missing data, no relatives (n = 374) and standardizing all features. The first two components captured a substantial proportion of the variance (Dim1: 59.6%; Dim2: 19%), with IQ and SRS scores emerging as the primary contributors to the first principal components, as shown by the PCA loading plot (see **Supplementary Fig. 21**). Specifically, IQ measures were the strongest contributors to the second principal component (full-scale IQ: 21.5%; performance IQ: 21.4%; verbal IQ: 15.9%), while SRS contributed substantially to both dimensions (8.99% to PC1 and 11.22% to PC2). In contrast, while VABS scores contributed strongly to the first principal component (12.6–14.8%), they contributed minimally to the second component (0.06–2.4%). Moreover, restricting the analysis to IQ and SRS allowed us to maximize the sample size (n = 1,026 people), whereas inclusion of VABS scores would have brought the sample size down to just over 500 people (See **Supplementary Table 12**). For these reasons, both statistical and practical, we focused subsequent clustering analyses on IQ and SRS dimensions only.

To determine the optimal number of clusters, we computed a comprehensive set of fit indices across k = 2 to 10. This multi-criterion approach is standard practice for cluster validation and addresses the challenge that no single method reliably identifies the optimal number of clusters across all datasets. We first applied an unsupervised approach focused exclusively on evaluating the intrinsic structure of the data using the NbClust R package[74], which implements 26 well-established internal validation indices simultaneously. The NbClust analysis identified 3 clusters as the optimal partitioning of the dataset (k = 2–10 tested), supported by 10/26 internal validation indices (*e.g.*, silhouette width, D index). Secondary peaks suggested k = 8 (5/26 indices), but the majority rule favored a 3-cluster solution.

To provide comprehensive validation beyond the NbClust consensus, we additionally computed multiple independent fit indices including information-theoretic criteria (Akaike Information Criterion (AIC), Bayesian Information Criterion (BIC), Integrated Classification Likelihood (ICL)), partition-based indices (Silhouette coefficient, Variance Ratio Score (VRS), Davies-Bouldin index, Pseudo F statistic), and reference distribution methods (Gap statistic). These analyses converged on k = 3 as optimal: the Variance Ratio Score, Gap statistic, and Pseudo F statistic all identified k = 3 as providing superior cluster separation (See **Supplementary Table 13**, with optimal values highlighted in bold).

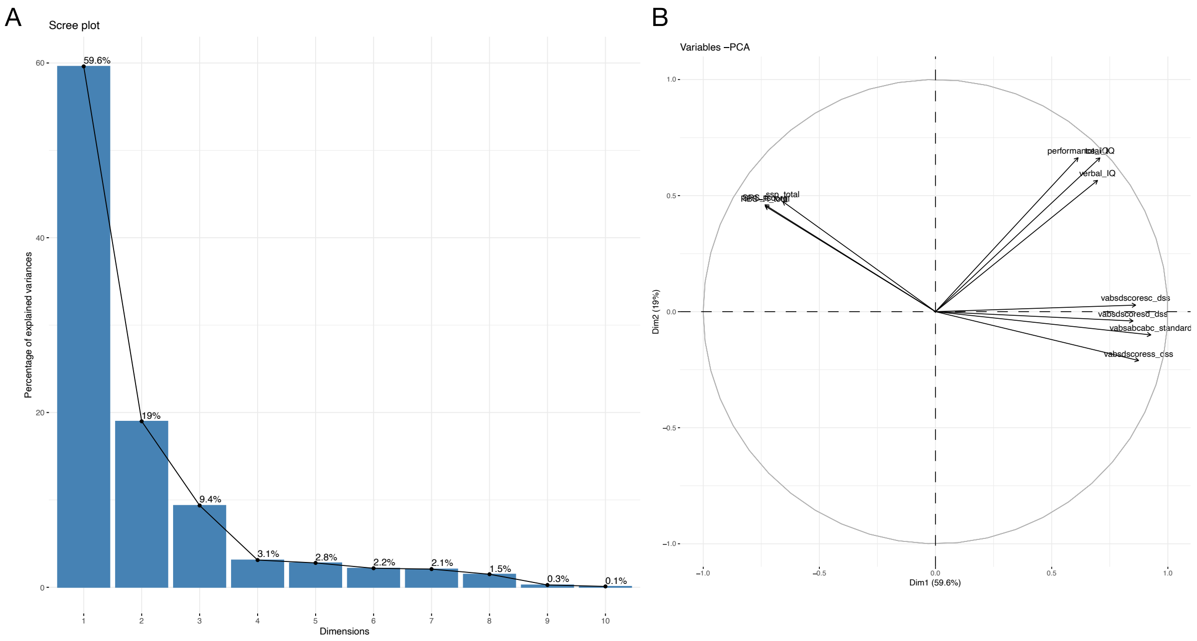

Supplementary Figure 21: Feature selection for clustering using principal component analysis (PCA). A. Scree plot showing the percentage of variance explained by each principal component. The first two components account for 59.6% and 19.0% of the variance, respectively. B. PCA biplot illustrating the projection of people and the direction and contribution of the 10 selected variables in the reduced dimensional space.

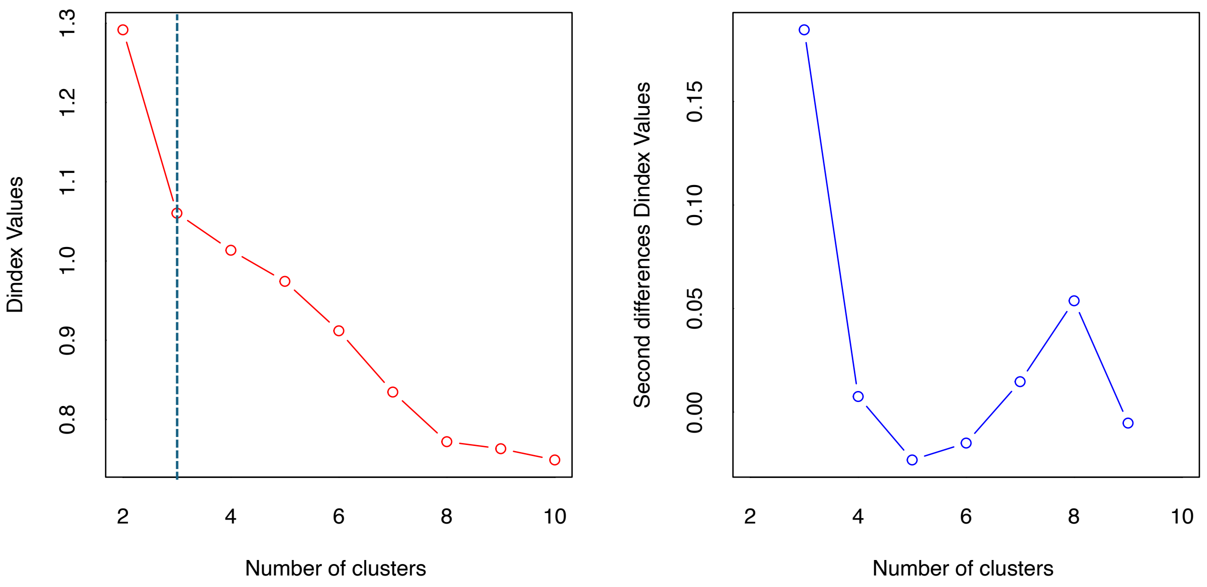

Supplementary Figure 22: Determining the optimal number of clusters using D Index and second differences. The analysis evaluated cluster solutions ranging from $k$ = 2 to 10 using multiple internal validation indices. The left panel shows D index values across different cluster numbers, and the right panel presents the second differences of the D index. A three-cluster solution emerged as optimal, supported by 10 out of 26 indices (e.g., silhouette width, D index)

Supplementary Table 12: Sample size completeness across variable combinations. The "IQ + SRS" combination provides the largest sample (N = 1,026; 38.7%), demonstrating parsimony advantages. Adding variables reduces sample size, justifying the selection of IQ and SRS-2 as the optimal 2-variable combination for clustering analysis.

**Sample Size Completeness Across Variable Combinations**

Number of participants with complete data for different combinations of clinical measures. Higher sample sizes provide greater statistical power for cluster validation.

| Variable Combination | N Variables | N Participants | % of Total Sample |
|---|---|---|---|
| IQ + SRS | 2 | **1,026** | 38.7 |
| IQ + SRS + RBS-R | 3 | 838 | 31.6 |
| IQ + SRS + SSP | 3 | 667 | 25.2 |
| IQ + SRS + ADI Social | 3 | 610 | 23.0 |
| IQ + VABS Composite | 2 | 565 | 21.3 |
| IQ + SRS + ADOS | 3 | 549 | 20.7 |
| IQ + SRS + VABS Composite | 3 | 511 | 19.3 |
| IQ + SRS + VABS + RBS-R | 4 | 481 | 18.1 |
| IQ + SRS + VABS + SSP | 4 | 418 | 15.8 |
| IQ + SRS + VABS + SSP + RBS-R | 5 | 409 | 15.4 |

Supplementary Table 13: Comprehensive fit indices computed for determining the optimal number of clusters (k = 2 to 10). Higher values are better for Silhouette, VRS (Variance Ratio Score), Gap statistic, and Pseudo F statistic, as these measure cluster separation and quality. Lower values are better for AIC (Akaike Information Criterion), BIC (Bayesian Information Criterion), ICL (Integrated Classification Likelihood), Davies-Bouldin index, and WSS (Within-cluster Sum of Squares), as these measure model complexity and within-cluster variance. Best values for each index across all k values are highlighted in bold. The optimal number of clusters (k = 3) was determined by consensus across multiple indices, with VRS, Gap statistic, and Pseudo F all identifying k = 3 as optimal. Abbreviations: AIC: Akaike Information Criterion; BIC: Bayesian Information Criterion; ICL: Integrated Classification Likelihood; VRS: Variance Ratio Score; WSS: Within-cluster Sum of Squares; Gap SE: Standard error of the Gap statistic (from B = 50 bootstrap iterations).

**Fit Indices for Determining Optimal Number of Clusters**

Higher values are better for Silhouette, VRS, Gap, and Pseudo F. Lower values are better for AIC, BIC, ICL, Davies-Bouldin, and WSS. Best values for each index are highlighted in **bold**.

| k | AIC | BIC | ICL | Silhouette | VRS | Davies-Bouldin | WSS | Gap | Gap SE | Pseudo F |
|---|---|---|---|---|---|---|---|---|---|---|
| 2 | 5237.68 | -5293.95 | -5527.21 | 0.380 | 739.30 | 0.900 | 1190.50 | 0.548 | 0.012 | 739.30 |
| 3 | 5217.82 | -5305.68 | -5891.80 | **0.405** | **977.56** | **0.828** | 704.18 | **0.569** | 0.010 | **977.56** |
| 4 | 5186.53 | -5306.00 | -6082.51 | 0.346 | 842.78 | 0.874 | 590.11 | 0.492 | 0.011 | 842.78 |
| 5 | 5187.17 | -5338.24 | -6308.30 | 0.316 | 795.42 | 0.930 | 498.03 | 0.475 | 0.010 | 795.42 |
| 6 | 5179.11 | -5361.78 | -6481.45 | 0.307 | 802.31 | 0.965 | 415.58 | 0.471 | 0.009 | 802.31 |
| 7 | 5185.44 | -5399.71 | -6664.02 | 0.333 | 845.58 | 0.860 | 342.87 | 0.489 | 0.008 | 845.58 |
| 8 | 5180.77 | -5426.64 | -6690.79 | 0.333 | 842.70 | 0.842 | 301.71 | 0.477 | 0.008 | 842.70 |
| 9 | **5172.89** | -5450.36 | -6678.28 | 0.335 | 833.16 | 0.860 | 271.38 | 0.477 | 0.009 | 833.16 |
| 10 | 5173.72 | **-5482.79** | **-6844.31** | 0.309 | 835.56 | 0.842 | **244.00** | 0.469 | 0.009 | 835.56 |

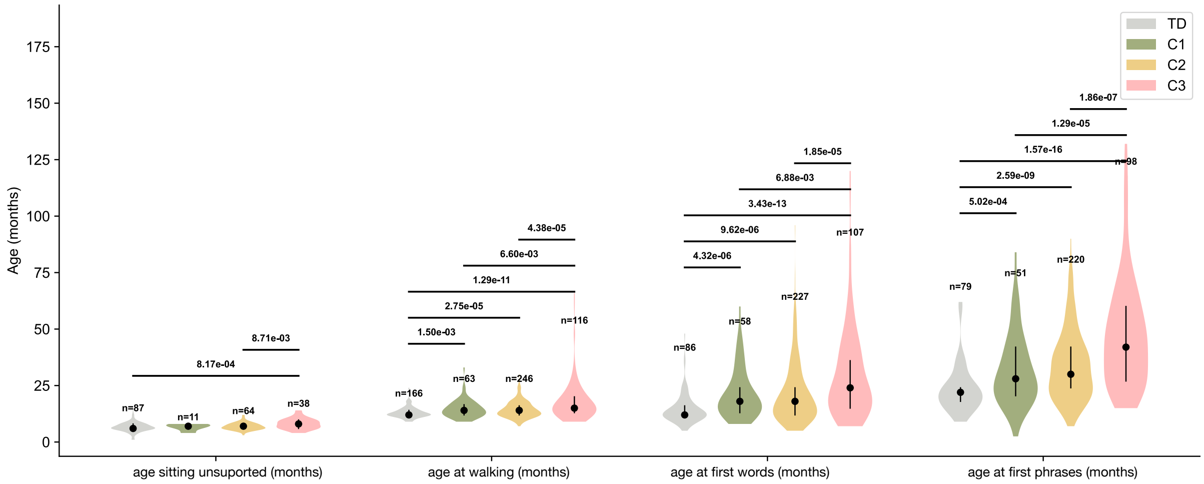

Supplementary Figure 23: Developmental milestone across the clinically derived clusters compared NT individuals. Lines connect groups with significant differences (Mann-Whitney U test, $p < 0.05$).

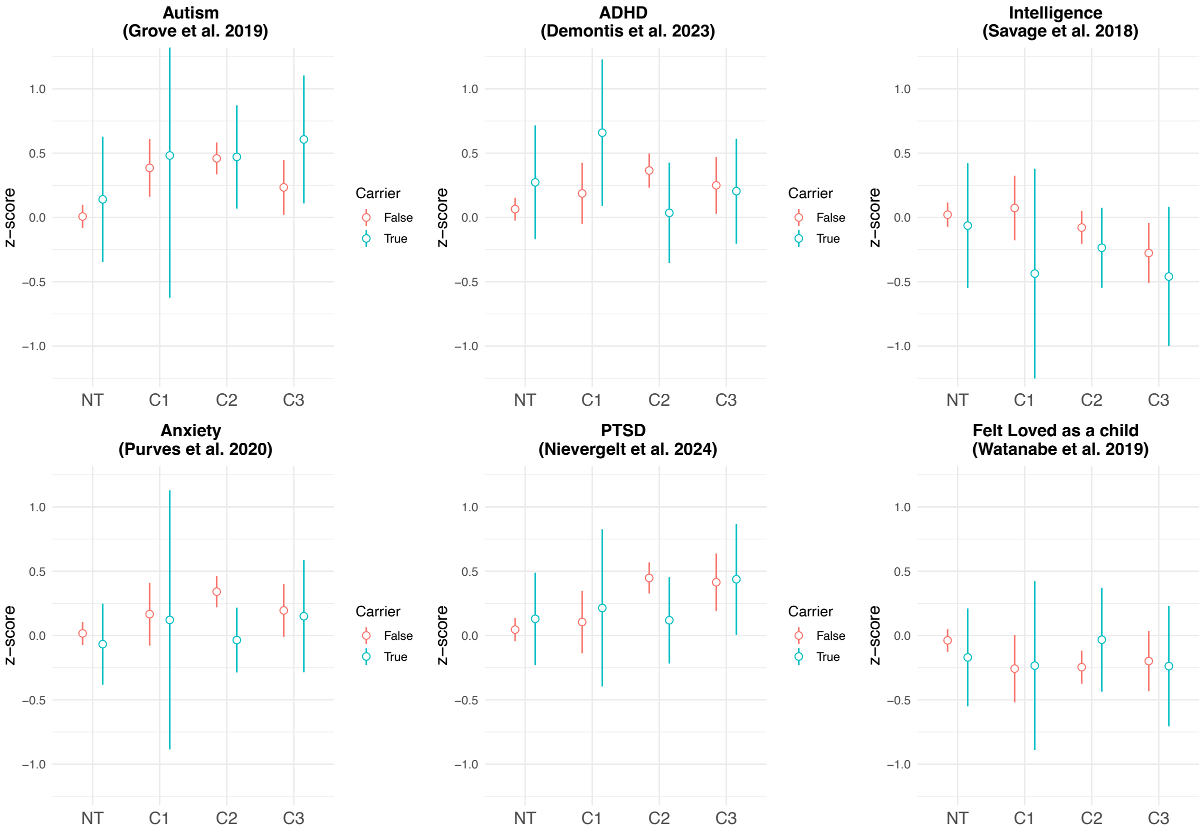

Supplementary Figure 24: Polygenic scores and rare variant in HCNDD carrier status across clinically derived clusters. Point plots display the distribution of six polygenic scores (PGS) across the three identified clusters with autism only (C1, C2, C3) and the NT. Colors indicate carrier status for rare variants in high-confidence neurodevelopmental disorder (HCNDD) dominant genes (carrier: True; non-carrier: False).

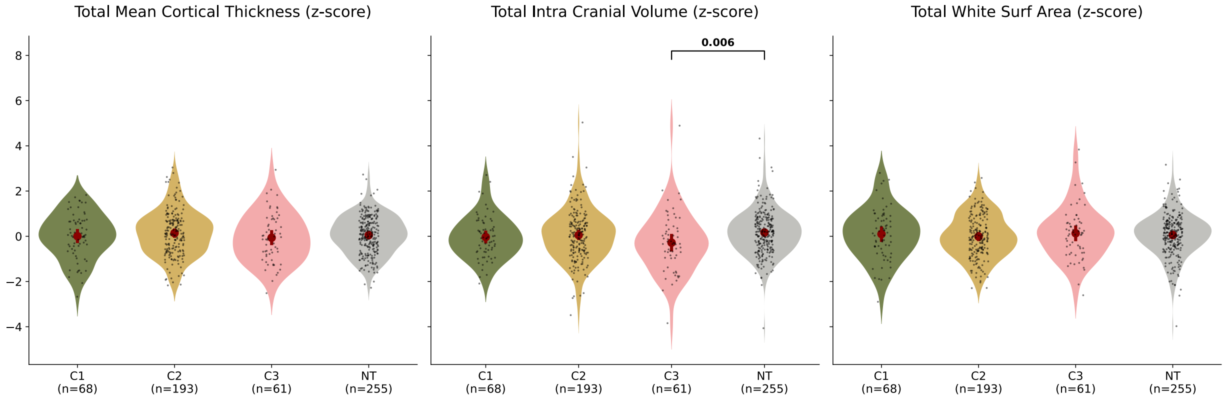

Supplementary Figure 25: Structural MRI measures (total mean cortical thickness, total intracranial volume, and total white surface area), z-scored, comparing autism clusters (C1, C2, C3) with NT people. Violin plots show the distribution of z-scored values for each measure, with individual data points, means, and 95% confidence intervals. Statistical comparisons between each cluster and NT were performed using t-tests or Mann-Whitney U tests (depending on normality assumptions) with FDR correction for multiple comparisons. Significant differences ($P_{FDR}$ < 0.05) are indicated on the figure.

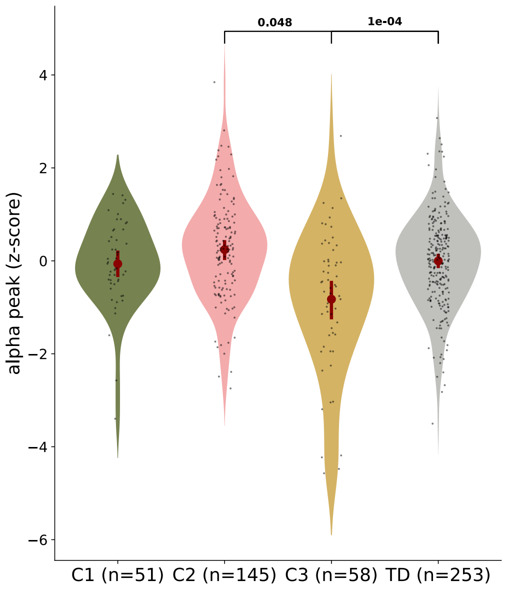

Supplementary Figure 26: EEG alpha peak frequency (6-13 Hz) in the occipital region, z-scored and corrected for age and sex, comparing autism clusters (C1, C2, C3) with neurotypical (NT). Violin plots show the distribution of z-scored values for each measure, with individual data points, means, and 95% confidence intervals. Statistical comparisons between each cluster and TD were performed using t-tests or Mann-Whitney U tests (depending on normality assumptions) with FDR correction for multiple comparisons. Significant differences ($P_{FDR}$ < 0.05) are indicated on the figure.

#### Section 7. Database and data sharing

The data management strategy for this research project encompasses the collection and storage of harmonized datasets, secure hosting and sharing with research partners, and post-publication dissemination to the scientific community (external data sharing). Two distinct processes were implemented for the two datasets, tailored to the specific requirements and constraints associated with each study (**Supplementary Fig. 27**).

##### ***EU-AIMS / AIMS-2-TRIALS LEAP***

***Data hosting:*** This multi-centric and highly multimodal project required a specialized platform capable of securely handling large datasets in a GDPR-compliant manner, and across several European research institutions.

To this end, the OWEY data lake, physically hosted at Institut Pasteur, was deployed. OWEY automates critical operations preceding data sharing: participant sharing consent verification, pseudonymization, automatic classification, and fine-grained access management. These processes are integrated into a seamless system that ensures regulatory compliance, interoperability, and operational efficiency. OWEY allows key features for storing and sharing data:

- **Automated participant sharing consent verification**: Access to data is contingent on the presence of valid consent, collected using the REDCap electronic data capture tools hosted at the Institut Pasteur [75,76]. If a participant withdrawn his consent to share data inside the research consortium, the corresponding data is hidden within OWEY, ensuring dynamic regulatory compliance.
- **Double pseudonymization**: Participant identifiers, already pseudonymized when uploaded, are systematically replaced with persistent anonymous codes (second pseudonymization), enabling traceability across data modality and data uploads, while minimizing the risk of re-identification.
- **Automated file organization and traceability**: All files are structured according to a multi-axis scheme (study, site, specialty, analysis level, etc.),

allowing for rapid retrieval and preventing duplication or data loss in a dynamic, asynchronous data flow.

- **Horizontal scalability**: The system supports increasing volumes of data, users, and concurrent connections through a distributed and flexible architecture.

***InovAND***

***Data hosting:*** InovAND is the center of excellence in Île-de-France region of France dedicated to autism and NDD. The dataset includes participants from 4 hospital centers. Unlike LEAP, data from InovAND is currently hosted on local servers at Institut Pasteur and partner institutions (*e.g.*, AP-HP, Fondation Fondamental).

***External data sharing***

The external sharing of all modalities from timepoint 1 of the LEAP study was completed in September 2025 and includes: clinical data, cognitive data, eye-tracking data, neuroimaging data (EEG, resting and functional MRI, anatomy), and genetic data [77]. All modalities from the InovAND dataset will similarly be shared in early 2026.

These datasets are securely stored on ELIXIR-LU servers, part of the European infrastructure for life science information, based at the Luxembourg Centre for Systems Biomedicine (LCSB) and supported by the Luxembourg National Data Service (LNDS) (https://elixir-luxembourg.org/). ELIXIR-LU provides a GDPR-compliant secure hosting infrastructure, and data is made accessible upon request via the data catalog website: https://datacatalog.elixir-luxembourg.org/ [78].

Access to the datasets would be granted based on each project proposal after review of the Data Access Committee, which includes members of the Consortium, scientific Data Modality leads, Ethics expert(s), and Autism community representatives. Prior to implementing external data sharing the AIMS-2-TRIALS consortium worked with autism community representatives (A-Reps – see below) to develop our data sharing plans to understand and address issues of importance to the community surrounding these endeavors (*e.g.*, unethical or unintended use, data security, biased data interpretation). This group also inputted on the development of the data sharing policies and consortium ethical principles required for sharing data, and design of the review committee and process.

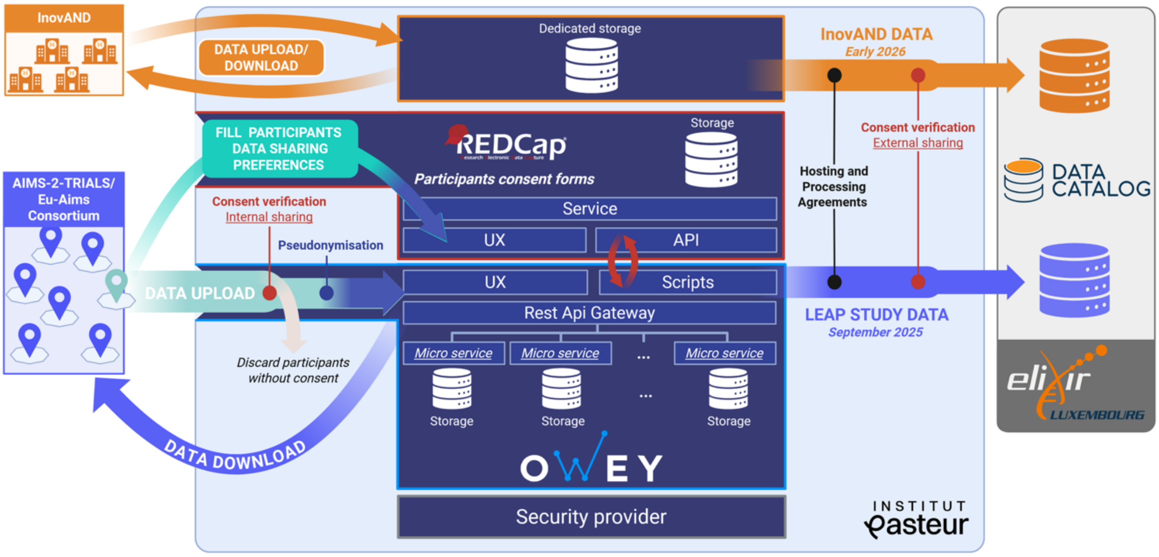

Supplementary Figure 27: Schematic representation of the data flows and data storage infrastructure for InovAND (top, orange) and LEAP data (bottom, blue). InovAND: data is stored on secure servers at Institut Pasteur and accessible for researchers within the institution. LEAP: participant's consent is collected by each center concomitantly with their data. Data-sharing choices expressed by participants upon consent collection are reported and maintained in a dedicated REDCap project hosted at Institut Pasteur. During the data upload from one center, data is automatically filtered based on participants consent for internal sharing (i.e. within the EU-AIMS consortium), verified using the REDCap API. The participants' identifiers are also pseudonymized, i.e. get attributed to a unique and anonymous identifier in OWEY. These steps allow each center to securely access and download data from all other centers and across modalities during the entire study run, in conformity with participants' data sharing choices expressed in the consent. The external sharing of the LEAP dataset required each participating research center to sign a data hosting and processing agreement with ELIXIR-LU, a European life sciences infrastructure offering hosting services. Prior to the data transfer from OWEY to ELIXIR-LU, data was filtered a second time based on the participants consent to share their data externally (i.e. outside the Eu-aims consortium). The dataset is available since September 2025 on the ELIXIR-LU data catalog: https://datacatalog.elixir-luxembourg.org/e/study/ELU-4-4D20BB-1. A similar process will be followed to share the InovAND dataset hosted at Institut Pasteur, with a targeted availability in early 2026.

#### Section 8. Frequently asked questions

**1. What are genes and how can they influence a person's development?**

Genes are made of DNA and contain information for building and maintaining our bodies, including our brains. Everybody has genetic variants and most of them have very small effects on the person's development. The average number of genetic differences between two unrelated individuals is estimated to be around 3-4 million. Compared to the size of the human genome, this is on average 1 variant each 1,000 nucleotides. A nucleotide is the building block of DNA that codes for different proteins in the human body, and the variants might impact the structure of the proteins they code for and/or modify how they work. These variants can be neutral, and they will have no consequence on the phenotype (what we observed). Some will have very small effects, and some will have a large effect. For example, a genetic variant that totally disrupts the function of a protein, also named loss of function (LoF) variant, that is important for the development of the brain might influence the person's development. If a variant has a functional effect on traits like learning, social interaction, and behaviour, it does not mean that the effect is always negative on the development of the person.

**2. If autism is largely genetic, why doesn't it always run clearly in families?**

This question highlights the complexity of genetics. There are two main reasons:

- *De novo* variants: Sometimes, a genetic variant happens in an individual and is apparently not inherited from the parents. The genetic studies are usually done on DNA isolated from a blood/saliva sample from the participants. Sometimes, a variant is observed in the child genome, but not in their parent. These *de novo* variants can either originate in the germ cells of the parents predominantly from the paternal side or they can appear at the first divisions of the egg then almost all the body of this individual will carry this *de novo* variant. If the variant appears at a later stage of the development, only some organs will carry this variant. We say that the person is mosaic for this variant (some cells have the mutation some don’t).

- A combination of genetic and environmental factors: in most of the cases, autism isn't the result of a single genetic variant but a combination of many common genetic variants that individually have a tiny effect. A person might inherit a particular set of these variations from their parents that, when combined, contribute to autistic traits. This is why the family pattern isn't always obvious. Another possibility is that two individuals with similar genetic profiles might be in different environments resulting in different autistic traits or support needs.

**3. What does a brain scan (like an MRI) measure? How is it different from an EEG?**

MRI (magnetic resonance imaging) is like a detailed, 3D photograph of the brain's structure. It shows the size, shape, and thickness of different brain areas.

EEG (electroencephalography) is more like a recording of the brain's activity. It measures the tiny electrical signals that brain cells use to communicate with each other in real-time. Think of MRI as a map of the brain's geography and EEG as a listen to its live conversation.

**4. What do we mean when we talk about differences in the autistic brain?**

Certain brain features may be different in autistic people compared to non-autistic people. This aligns with the neurodiversity perspective, which suggests that autism is a natural variation in the brain, and is not a defect, simply differences. These differences can come with both challenges and strengths. The goal of this research is to better understand how the brain works to provide better support options for autistic people who need them.

**5. How can you tell if a genetic change or a brain difference is related to autism, and not just a natural variation?**

All variants are considered natural. This is the occurrence of genetic variations that have produced differences between people. Scientists study groups of autistic people and compare them to groups of non-autistic people at a large scale. If a specific genetic change or brain feature appears significantly more often in the autistic group, it suggests a correlation. However, a significant correlation between a genetic variant and autism is not a formal proof that this variant contributes to autism. This difference between correlation and causality is addressed in **FAQ #7**.

Importantly, it is worth noting that there is a lot of variation within autism and characteristics and traits will be different amongst autistic people. This is why subgrouping is important, whilst also considering everyone as unique.

**6. What is the connection between genetic variants, the brain, and actual behavior? Is it direct?**

The connection is complex and is like a chain of events within the body:

a. The products of the genes (RNA and proteins) are used to produce cells. These cells will then divide, differentiate, migrate and connect during development. Some genes associated with autism will play a key role in these processes and a genetic variant affecting the function of these genes might influence the probability of having an autistic development.

b. Cells are in an environment with other cells, and the genetic variant can also influence the differentiation of a specific type of cell. This was very nicely illustrated by Conrad Hal Waddington as the epigenetic landscape (**Supplementary Fig. 28**). This is a powerful metaphor that helps explain how genes play a role in development. It illustrates marbles rolling down a hill that has grooves and valleys, and when the marbles reach the lowest point, this represents the different cell 'fates' and tissue types.

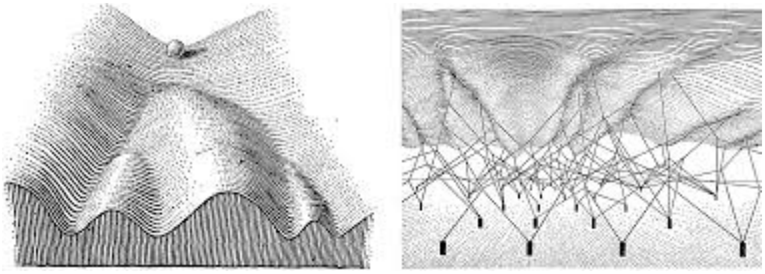

Supplementary Figure *28* Waddington's epigenetic landscape illustrating how a cell's developmental trajectory is guided by gene regulation and epigenetic factors, leading to distinct cell fates*.*

c. Through brain development, all kinds of cells including the glial cells (non-neuronal cells in the central nervous system) and the neurons, will make connections that are necessary to transfer and store information in different regions of the brain. One of the most well-known junctions between nerve cells is called the synapse. Many genes involved in the formation and the functions of these synapses are associated with autism. The genetic variants within these genes might reduce or increase the strength of a specific synapse or influence the number of these synapses. The genetic variants can also cause an imbalance between the inhibitory and the excitatory synapses. Such imbalance in excitation and inhibition is a well-known cause of epilepsy. Many genes associated with epilepsy are also associated with autism. Why some people carry such genetic variants will have either epilepsy or autism or both or neither remains largely unknown.

c. The way these circuits' function influences how a person perceives the world, thinks, and acts, their behaviour. It's not a simple "one gene = one behaviour" relationship. The brain is incredibly complex, and experiences also shape its connections throughout life.

**7. What is the difference between correlation and causality in genetic studies?**

Correlation and causality are two distinct concepts often discussed in scientific research, including genetic studies. Understanding the difference between them is crucial for interpreting research findings accurately. Correlation refers to a statistical relationship between two variables, indicating how they change together. It does not imply that one variable causes the other. In contrast, causality refers to a direct cause-and-effect relationship between two variables, where one variable directly influences the other. While correlation is a relationship between variables, causality requests stronger evidence often including experimental data and biological mechanisms.

In this study, we used a novel independent dataset of autistic and neurotypical (non-autistic) people not studied before, and we confirmed that autistic individuals have on average more genetic variants that modify the function of genes previously known to be associated with neurodevelopment conditions (*e.g,*, at genes that play key roles during brain development, such as chromatin remodeling and synaptic functions). We also show that common variants found frequently in the general population and previously associated with autism are also more frequently observed in the autistic people compared to non-autistic individuals in this new LEAP-InovAND dataset. Remarkably, these genetic results are observed separately in the LEAP and the InovAND dataset. Altogether, these results indicate that the statistical correlation between these genetic variants and autism is very robust.

**8. How do environmental factors fit into the picture if autism is genetic?**

There is the interplay between genetic variants and environment that will influence behavior. At the individual level, depending on the impact of the genetic variants and on the environmental factors, those factors will influence everyone differently. Sometimes genetics will play a large role, sometimes the environment will play a large role. In autism, twin studies indicate that 80% of the effect is genetics. This does not preclude that a new environment might change the quality of life of autistic people. The role of the environment in development shows the potential for the development of medications and person-centered support for those with co-occurring medical conditions. There is a need to tailor support for people who want and need it.

Because heritability of autism is above 80%, this means that genetic variants play a large part of the difference between an autistic and a non-autistic individual.

**9. What is the most important takeaway from this research for the public to understand?**

The most important takeaway is that autism is diverse, not one single condition. It is heterogeneous with diverse biological roots. By combining genetics and brain science, we are starting to map out this diversity. In this study, with this sample size, we could find three different subgroups of autism. One type in cluster 1 includes mostly neurotypical people and some autistic people, people in cluster 2 have medium to high autistic traits, and average to high measured IQ. Finally, the last cluster, cluster 3, includes autistic people with medium to high autistic traits with very low measured IQ. These subgroups differ in terms of their genetics and brain structure and function. Analysis of data collected on features such as sensory information and functionality/autonomy, would likely have identified more subgroups and this needs further investigation in future research. This understanding of the variation within autism is a crucial step away from stereotypes and towards a future where support and understanding can be tailored to everyone's unique needs and strengths.

**10. Why subgrouping participants by ancestries?**

One confounding factor in genetic association studies is the ancestry of the participants. If the population of autistic people has a different ancestry compared to the NT then the difference in genetic variant frequency might be due to the genetic differences between populations rather than a difference between autistic and neurotypical people. In this study, we run the analyses either only in participants from European ancestries or in all participants. We chose to present the pan ancestry results in the main text and the European only ancestry in the supplementary results.

**11. What are polygenic scores, how do you compute them and what do they mean?**

Polygenic scores (PGS) provide a quantitative measure of a person's genetic predisposition to a trait. Higher scores typically indicate a higher probability to have the trait, while lower scores indicate a lower probability. PGS are important to understand the genetic architecture of a trait, it is <u>not</u> a prediction that someone with high PGS for the trait has the trait.

To compute the PGS for a specific trait, you first need a genome wide association study (GWAS) for this trait. The GWAS will compare the genetic profile of people with and without a specific trait. It will detect variations known as common single nucleotide polymorphism (SNPs), which are more frequent in the people with the trait compared to the people without the trait. The traits can be categorical (*e.g.*, autistic *vs.* non-autistic people) or quantitative (*e.g.*, measured IQ scores, numbers of years of education). The GWAS will provide for each SNPs an odd ratio or a *beta* value SNPs that indicate the level of the effect on the trait.

In the vast majority of the GWAS, each SNP is associated with a very low effect size (odd ratio, OR <1.1). However, because they are many, collectively they explain a significant percentage of the heritability of a trait.

In this study we used several GWAS to compute the PGS of different traits for each participant of LEAP-InovAND. Then, we divided the participants in four groups (quantiles) with low PGS (Q1), medium low PGS (Q2), medium high PGS (Q3) and high PGS (Q4). Finally, we compared the proportion of autistic and neurotypical people in the quantiles Q4 (high PGS) vs Q1 (low PGS).

Importantly, if someone has a high PGS for a trait, it does not mean that this person has the trait. What it means is that this person accumulates genetic variants that were previously identified as more frequent in people with this trait. See **FAQ #7** about correlation and causality.

Here are three examples:

a. We used the PGS for anxiety and major depression and found that autistic people had higher PGS for these traits compared to NT individuals. It means that on average autistic individuals accumulate genetic variants that were previously detected more frequently in people with anxiety or major depression compared to the general population. These genetic results do not imply that people with autism will always have more anxiety and major depression compared to non-autistic people or that people with anxiety or depression will be more likely to be autistic. It means that some of the genetic variants associated with anxiety and major depression might be shared with autism and that there is a higher likelihood of co-occurrence.

b. We used the PGS for intelligence and found that autistic individuals with low IQ scores have also on average lower intelligence PGS compared to the neurotypical participants. Nevertheless, someone with low or high PGS for intelligence will not always be in the group of low or high IQ respectively. We also acknowledge that intelligence is a difficult trait to measure especially in people with autism.

c. We used the PGS for the question “felt loved as a child”. The initial GWAS was performed in the UK Biobank where 170,000 participants were asked to answer this question “Did you feel loved as a child?” The participants had to choose between several answers (very often true, often true, sometimes true, rarely true, never true). For each answer, a score has been assigned and the GWAS has been performed. Genetic variations were identified as being more frequent in the people who answered positively (often true as a maximum score) compared to those who answered negatively (never true as a minimum score). If someone has a low PGS for this trait, it most likely reflects that genetic variations can influence the way we think we are accepted positively as a child. It does NOT imply that this person felt not loved as a child or that this person was not loved as a child. It seems that these variants are less frequent in autistic individuals compared to non-autistic individuals from the LEAP-InovAND dataset and in autistic individuals compared to their nonautistic siblings from the SPARK cohort.

Understanding the genetic architecture of autism is an important piece of information to explain, for example, why the level of anxiety is high in people with autism. It does not mean that this genetic susceptibility for anxiety is a fate. Recognizing the diversity of people with autism and providing a more inclusive environment should reduce the level of anxiety of people with autism. Having low levels of PGS for “felt loved as a child” could represent a risk factor for bad quality of life. But again, this risk could be reduced if there was a better recognition of the diversity of the child and a better inclusion early in life.

**12. A low PGS of ‘felt loved as a child’ means that autistic people are less loved as a child than non-autistic people?**

No, this analysis indicates that genetic variants associated with “felt being loved as a child” are less frequent in autistic people compared to non-autistic siblings.

In this analysis, we used the results from a previous genome wide association study (GWAS) for which participants from UK Biobank answered a question about whether they felt loved in childhood (see below the technical note on the GWAS was performed).

Using this GWAS, we found three results:

*Result #1:* We first calculated the genetic correlation (noted rg) between the autism GWAS and the “felt loved as a child” GWAS. This analysis quantifies how much of the genetic signal is shared between the two traits. We found a correlation rg of -0.39 which means that some SNPs from both GWAS are shared but their effects move in opposite directions. In other words, some of the alleles that increase the likelihood of “felt loved as a child” decrease the likelihood to be autistic.

*Result #2:* We then computed the PGS for “felt loved as a child” for all participants from the LEAP-InovAND dataset. We found that autistic individuals have lower PGS for “felt loved as a child” compared to non-autistic people.

*Result #3:* Finally, we computed the same PGS in an independent cohort of the SPARK of 24,791 autistic people and 15,192 non-autistic siblings. Again, we found that autistic people had lower PGS for “felt loved as a child” compared to their autistic individuals.

These results indicate that autistic people have on average lower PGS for “felt loved as a child” compared to non-autistic individuals or their non-autistic siblings. These results should be integrated with the results obtained from other PGS. We know that the genetic variants associated with autism are also associated with different traits related to psychiatric conditions and to well-being. For example, autistic individuals also have higher PGS for anxiety and depression. These two traits are also negatively correlated to “felt loved as a child”. Future studies would address if some variants involved in the “felt loved as a child” phenotype have a specific contribution to autism

or if this genetic correlation between autism and “felt loved as a child” is mostly driven by the genetic variants associated with anxiety/depression.
Most importantly, this anticorrelation does NOT mean that parents love children with autism any less.

**Technical note on how the GWAS for “felt loved as a child” was performed:** The paper that made the **“felt loved as a child”** GWAS for the first time was published by Watanabe et al. Nature Genetics 2019 (DOI: 10.1038/s41588-019-0481-0). In 2017, 157,130 participants answered a question about whether they felt loved in childhood (https://biobank.ctsu.ox.ac.uk/ukb/field.cgi?id=20489). They answered Very often true: 8,487; Often: 39,801; Sometimes true: 25,613; Rarely true: 7304; Never true: 2,300; Prefer not to answer: 625. The GWAS has been performed using a linear model testing 9564533 SNPs on 126,348/157,130 (80%) people (https://atlas.ctglab.nl/traitDB/3748). The heritability captured by SNPs is 0,073 ± 0,0048.

**13) What specific support could these findings help to develop for autistic people?**

These findings could help develop practical and personalised support for autistic people rather than just stating scientific “facts”. By combining clinical data, genetics and brain imaging, we can better understand the unique developmental trajectory autistic people experience. This could, in the future, lead to:

a. Tailored strategy for daily life support, understanding neurological and genetic differences can influence support that matches people strengths and challenges, such as communication tools, sensory adjustments or learning approaches.
b. Early and proactive support: biomarkers from genetic and brain imaging could help identify needs earlier, allowing timely educational or social support.
c. Informed guidance for caregivers and professionals: families, educators and clinicians could use these findings to choose strategies that are most likely to benefit each person and to anticipate/monitor co-occurring conditions (such as epilepsy), if

certain genetic variants or brain features indicate higher probability of responding to a certain intervention or support.

d. Enhanced well-being: understanding people's profiles can help reduce stress, sensory overload and other difficulties, improving quality of life by tailoring supports that work best for certain groups of people.

In a nutshell, these findings could in the future support a custom-built, strength-based approach that helps autistic people develop in ways that suit their personal needs

**14) Would the research team consider engaging with Critical Autism Studies in addition to combining genetic and brain imaging findings?**

Yes. We are eager to engage with Critical Autism Studies and to co-produce work with autistic scholars and community members. Beyond integrating genetics and brain imaging, we will embed participatory research approaches that center lived experience and real-world outcomes. For partnership and governance, we already created an advisory panel of autistic researchers and advocates. This includes autistic co-investigators/ co-authors who provide input across design, analysis, and dissemination.

**15) Would the research team acknowledge that the deficit approach that has framed this work is a limitation as it does not reflect community models and understanding (*e.g.*, monotropism, Double Empathy, Neurodiversity Paradigm)?**

Yes, this is an essential limitation of our study and one that affects much of the autism literature. Although these datasets are not limited to a binary autistic/non-autistic subgrouping and include several dimensional questionnaires, the projects were launched more than a decade ago and reflect the measurement priorities and diagnostic practices of that time. As a result, some domains now considered important are underrepresented (such as sensory characteristics). We therefore interpret findings with caution and emphasize analyses that leverage the available continuous measures. Future work is needed to develop autism traits, IQ and daily functioning measures in collaboration with autistic people that reflect autistic people's experiences and a neurodiversity affirming approach to autism.

**16) Why is it important to do research looking at subgroups within autism?**

Autism is highly heterogeneous. In the seventies, Lorna Wing already popularized the term “autistic spectrum” to capture this heterogeneity and to steer research and practice toward dimensional, person-centered approaches. Autistic people differ in communication, language, cognition, sensory profiles, co-occurring conditions, support needs, and strengths.

Subgrouping helps make sense of this diversity.

- It improves scientific signals. Grouping people with more similar characteristics reduces averaging effects and increases power and reproducibility. In our study, certain genetic architectures and brain-anatomy patterns were enriched in specific subgroups.
- It enables better support. Subgroups can guide tailored accommodations, services, and educational or clinical strategies, moving away from one-size-fits-all approaches. A next step for this project could be to collect systematic data on supports received by LEAP-InovAND participants and evaluate their effectiveness. These projects were designed to be observational rather than clinical trials for certain interventions, but it could give some initial data that could be built on with future research.
- It informs clinical trials. Subgrouping by relevant features (*e.g.*, language level, co-occurring ADHD/anxiety, presence of specific genetic variants) helps match interventions to those most likely to benefit and clarifies why results vary.
- It clarifies mechanisms. Different genetic, neural, and environmental pathways may underlie different profiles; subgroup analyses can reveal these pathways. Our data implicate genes involved in chromatin remodeling and synaptic function across clusters, with stronger associations with low IQ.

- It supports equity and planning. Understanding who has which needs, and when across the lifespan, helps allocate resources and reduce disparities in access and outcomes.

Key principle to keep in mind: Subgroups are tools, not labels. Boundaries are not defined, people may fit multiple subgroups, and profiles can change over time. In the future, such stratification analyses will include participatory research methods. They will define and interpret subgroups with autistic people and the families' prioritizing outcomes that matter to them (quality of life, autonomy, participation). There is the need to validate subgroups across datasets and link them to real-world outcomes rather than only diagnostic categories.

**17) What do you mean by the term 'neurotypical' in this paper?**

In this paper, we have used the term 'neurotypical' based on the survey on autism-related language preference published by Keating *et al.*, 2023 [79]. By "neurotypical," we refer to participants without a clinical diagnosis of autism or intellectual and developmental disabilities (IDD) at the time of enrollment. Notably, some individuals assigned to the neurotypical group had elevated Social Responsiveness Scale (SRS-2) scores, indicating higher levels of autistic traits despite no diagnosis. Similarly, some participants with IDD showed elevated SRS scores and, in some cases, might meet criteria for an autism diagnosis. We report these overlaps because they reflect the dimensional nature of social-communication traits and the porous boundaries between diagnostic categories.
